# Ambient PM_2.5_ Concentration and the Cardiovascular Response to Exercise Training: A Systematic Review and Meta-Analysis Across Global Pollution Gradients

**DOI:** 10.64898/2026.08.20.26360886

**Authors:** James A. Donaldson, Samuel Cai, Anna L Hansell, Joshua Vande Hey, Rikesh Panchal, Jamie Edwards, Ahmed Abdelrazik, Thomas Yates, Andre Ng, Jamie O’Driscoll

**Affiliations:** Department of Cardiovascular Sciences, University of Leicester, Leicester, United Kingdom; Centre for Environmental Health and Sustainability, Division of Public Health and Epidemiology, School of Medical Sciences, University of Leicester, Leicester, United Kingdom; NIHR Leicester Biomedical Research Centre, Leicester, United Kingdom; Leicester British Heart Foundation Centre of Research Excellence, University of Leicester BHF Cardiovascular Research Centre, Glenfield Hospital, Leicester, United Kingdom; NIHR Health Protection Research Unit in Chemical Threats and Hazards at the University of Leicester, United Kingdom; Diabetes Research Centre, College of Life Sciences, University of Leicester, Leicester, United Kingdom; Leicester Diabetes Centre, Leicester General Hospital, University Hospitals of Leicester NHS Trust, Leicester, United Kingdom; School of Health, Sport and Bioscience, University of East London, London, United Kingdom

## Abstract

**Background:** Exercise training is a cornerstone intervention for cardiovascular disease, yet large cohort studies have reported attenuation of physical activity benefits at elevated air pollution concentrations, creating uncertainty around exercise prescription in polluted settings where cardiovascular disease burden is greatest.

**Objectives:** To determine whether ambient PM_2.5_ concentration modifies the cardiovascular benefits of structured exercise training, using a global sample of trials spanning a >100-fold pollution gradient.

**Methods:** We conducted a systematic review and multilevel meta-analysis of exercise training interventions reporting pre–post changes in systolic blood pressure (SBP), diastolic blood pressure (DBP), peak oxygen uptake (VO₂max), or resting heart rate (HR) in adults. Annual ambient PM_2.5_ concentrations (3.5–283 µg/m³) were assigned to each study location from CAMS ERA5 reanalysis data. Three-level random-effects models with cluster-robust variance estimation accounted for arms nested within studies. PM_2.5_ meta-regression was conducted unadjusted and adjusted for world region, exercise mode, trial duration, and health condition, with subgroup analyses by exercise mode and hypertension status.

**Results:** Across 465 studies (27,629 participants), exercise training produced clinically meaningful benefits for all outcomes (SBP −6.1 mmHg, DBP −3.3 mmHg, VO₂max +3.2 ml/kg/min, HR −2.8 bpm; all p < 0.001), with benefits consistently larger in higher-pollution settings. Hypertensive participants showed the greatest improvements, particularly from aerobic exercise (SBP standardised mean difference 0.396 in the Low vs 1.020 in the High PM_2.5_ stratum). Although aerobic and resistance training participants experience similar chronic ambient PM_2.5_ exposure, only aerobic exercise showed a stratum gradient (interaction p = 0.074).

**Discussion:** Exercise training delivers clinically meaningful cardiovascular benefits at every pollution level tested. The larger benefits observed in higher-pollution settings reflect the greater cardiovascular risk burden of those populations, and hypertensive patients stand to gain the most, particularly from aerobic exercise.

## Introduction

Hypertension affects approximately 1.3 billion adults worldwide and is the leading modifiable risk factor for cardiovascular disease, stroke, and premature mortality (World Health Organization 2024). Structured exercise training is recommended by international clinical guidelines as a first-line non-pharmacological intervention for hypertension, with meta-analytic evidence demonstrating reductions in systolic blood pressure of 5–8 mmHg across diverse populations (Cornelissen and Smart 2013; Pescatello et al. 2019). Recent updates to the European Society of Cardiology (ESC) guidelines (McEvoy et al. 2024) and the American Heart Association (AHA) clinical practice guidelines (Pescatello et al. 2024) continue to emphasise exercise as a cornerstone of blood pressure management. Exercise also improves cardiorespiratory fitness, a powerful independent predictor of cardiovascular mortality (Ross et al. 2016).

Ambient air pollution, particularly fine particulate matter (PM_2.5_), is recognised as a major cardiovascular risk factor, contributing to an estimated 4.2 million premature deaths annually (Cohen et al. 2017). Long-term PM_2.5_ exposure is associated with increased incidence of hypertension (Cai et al. 2016), endothelial dysfunction (Rajagopalan et al. 2018), and accelerated atherosclerosis (Münzel et al. 2021). During exercise, minute ventilation increases up to eightfold, substantially amplifying the inhaled dose of airborne pollutants (Giles and Koehle 2014). This raises a clinically important question: does chronic exposure to ambient PM_2.5_ attenuate the cardiovascular benefits of exercise training?

The existing evidence is mixed and spans different study designs. Acute exposure studies have demonstrated transient attenuation of cardiovascular benefits when exercising in polluted environments (Sinharay et al. 2018). Observational cohort studies examining mortality outcomes have yielded nuanced findings: Guo et al. (2020) reported that habitual exercise prevented hypertension even in polluted areas, though protective effects diminished at PM_2.5_ exceeding 54 µg/m³. Most recently, Ku et al. (2025) pooled individual-level data from seven cohorts (n = 1.5 million adults) and found that the mortality reduction from leisure-time physical activity was attenuated at PM_2.5_ > 25 µg/m³, although exercise remained beneficial at all pollution levels.

However, these studies examined habitual physical activity and long-term mortality, a fundamentally different exposure contrast from structured exercise training and its chronic physiological adaptations (blood pressure, cardiorespiratory fitness). No study has systematically examined whether ambient pollution levels modify the cardiovascular adaptations to structured exercise training across a global pollution gradient.

This evidence gap has direct clinical consequences. In the absence of clear guidance, clinicians in high-pollution settings, disproportionately located in low- and middle-income countries, face uncertainty about whether to prescribe outdoor exercise without environmental caveats. Public health messaging that emphasises pollution risks during physical activity, while well-intentioned, may inadvertently discourage exercise in populations that would benefit most from it.

We therefore conducted a systematic review and multilevel meta-analysis to determine whether ambient PM_2.5_ concentration modifies the effects of structured exercise training on blood pressure, cardiorespiratory fitness, and resting heart rate. We analysed data from 465 studies spanning four continents and a >100-fold range of PM_2.5_ exposures, employing regional stratification and five pre-specified sensitivity analyses to distinguish genuine pollution-modification effects from ecological confounding.

## Methods

### Protocol Registration

This systematic review was prospectively registered with PROSPERO (registration number: CRD420251068843).

### Search Strategy and Study Selection

We searched MEDLINE, Embase, and PubMed for studies investigating exercise training interventions and cardiovascular outcomes in adults. The search combined terms for exercise interventions (exercise program, exercise therapy, physical activity program), blood pressure outcomes (blood pressure, hypertension, mean arterial pressure), and study settings. The search was conducted on 3 November 2025; full search strategies for all databases were published with the registered protocol on PROSPERO (CRD420251068843). Study selection was performed by three reviewers using Rayyan software through four systematic screening stages: (1) study design, adult population, and exercise intervention type; (2) measurement of blood pressure (or another eligible cardiovascular outcome: VO₂max or resting HR) before and after intervention; (3) clearly reported study location with available air pollution data; and (4) data extraction. The PRISMA flow diagram (Figure 1) details study numbers at each stage.

**Figure 1.**
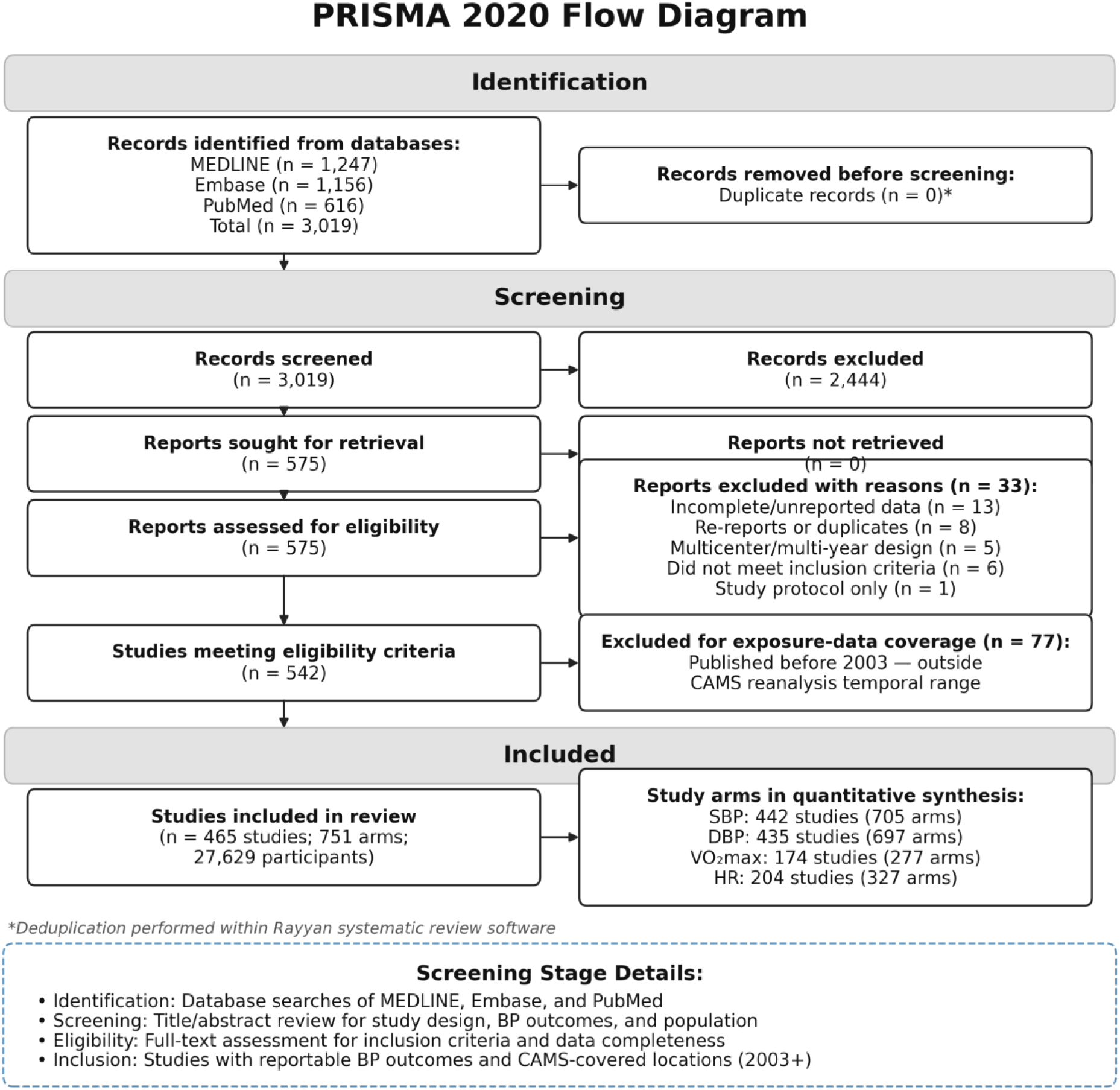
PRISMA flow diagram of study selection.

### Inclusion and Exclusion Criteria

We included peer-reviewed studies reporting pre–post changes in SBP, DBP, resting HR, or VO₂max following structured exercise interventions of ≥4 weeks in adults (≥18 years) with or without diagnosed hypertension. Studies involving participants on antihypertensive or other medications were included provided the medication regimen remained stable throughout the trial; studies comparing exercise against pharmacological interventions were not included. We excluded cross-sectional studies, case reports, reviews, conference abstracts, single acute exercise sessions, studies in children or pregnant women, and studies without objective blood pressure measurements or identifiable study location. Cross-over trials were included using only the first-period data where available, or full cross-over data where period-specific results were not separately reported. One study (Jakicic et al. 2015) was excluded post hoc because the intervention was delivered entirely by telephone with no fixed study site, precluding pollution exposure assignment.

**Figure 2.**
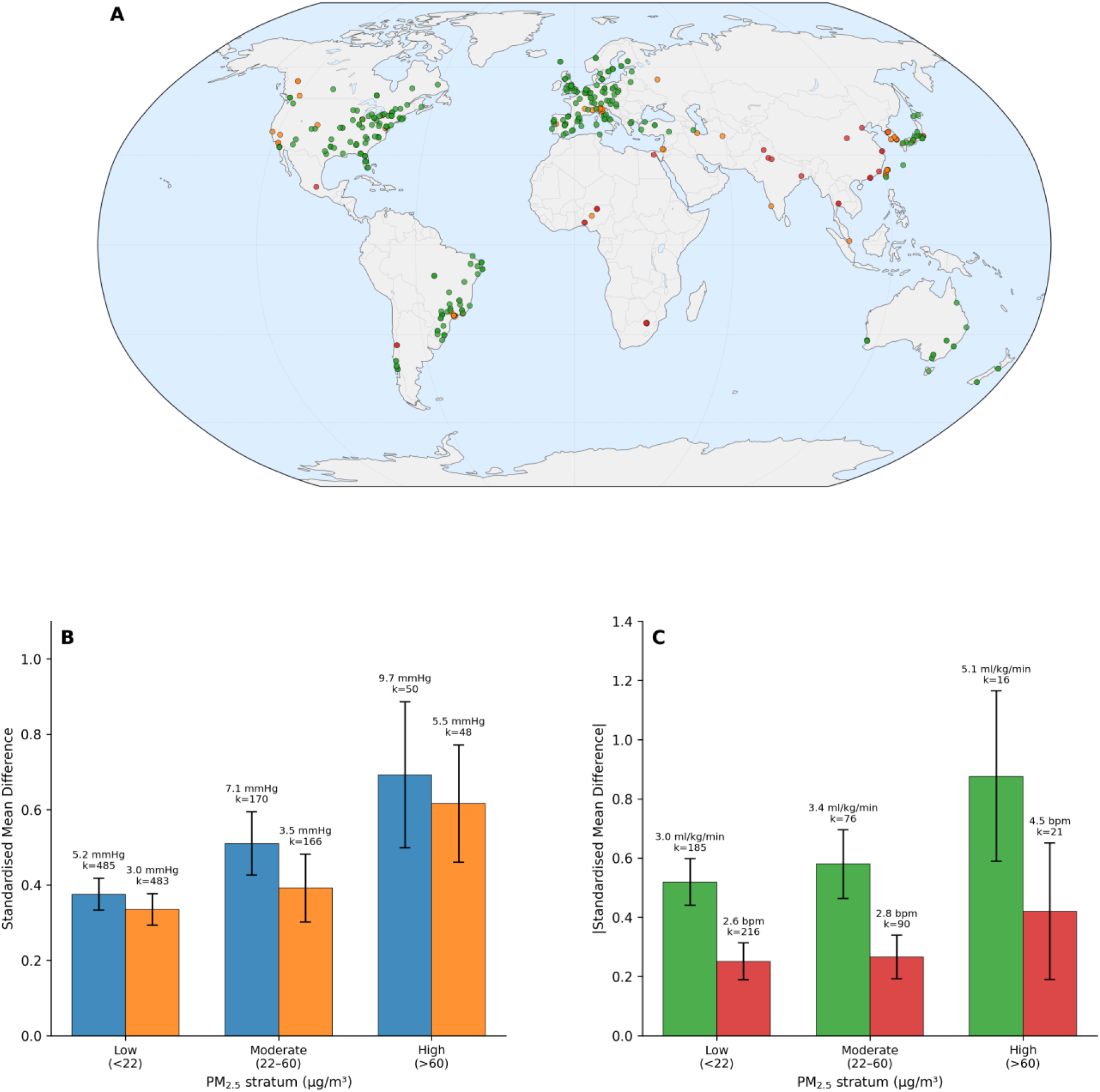
A) Geographic distribution of included studies (n=465) by PM_2.5_ stratum. B) Blood pressure effects by stratum. C) VO₂max and HR effects by stratum.

### Data Extraction and Air Pollution Exposure Assessment

Data were extracted using standardised forms capturing study characteristics (author, year, location, design), participant characteristics (sample size, age, health condition), intervention details (exercise type, intensity, duration), and cardiovascular outcomes (pre–post means and standard deviations). Annual mean ambient PM_2.5_ concentrations were assigned to each study location using the Copernicus Atmosphere Monitoring Service (CAMS) ERA5 global reanalysis dataset (≈80 km resolution). Study cities were geocoded and matched to the nearest grid cell.

CAMS monthly mean PM_2.5_ data are available from 2003 to 2021; studies published before 2003 (n = 77) were excluded as they fall outside the CAMS temporal coverage. The final PM_2.5_ range was 3.5–283 µg/m³. Hypertension status was classified according to baseline blood pressure values reported by each study rather than by regional guideline thresholds, with participants categorised as hypertensive (SBP ≥140 or DBP ≥90 mmHg, or study-reported diagnosis), pre-hypertensive (SBP 120–139 or DBP 80–89 mmHg), or normotensive. Exercise modalities were classified as aerobic, resistance, combined, or mind-body. Isometric exercise training (IET), including isometric handgrip and wall squat protocols, was classified under the resistance training category.

### Effect Size Calculation

Standardised mean change scores (SMCR; Becker 1988) were calculated as Hedges’ g for each arm–outcome combination. Pre–post correlations were assumed as 0.70 for SBP and DBP, 0.65 for VO₂max, and 0.55 for HR (Rosenthal 1991; Morris and DeShon 2002). Positive values indicate beneficial reductions in SBP, DBP, and HR; negative values indicate beneficial improvements in VO₂max.

### Statistical Analysis

We fitted three-level random-effects models using restricted maximum likelihood (REML) estimation in metafor for R (Viechtbauer 2010), with σ²₃ (between-study) and σ²₂ (within-study, between-arm) variance components. Cluster-robust standard errors (CR2 estimator; clubSandwich package) were applied throughout (Pustejovsky 2022). Effect sizes with |yᵢ| > 3 were excluded as outliers, with a sensitivity analysis retaining all observations. Full model equations and estimation details are provided in Appendix S1.

PM_2.5_ meta-regression used log-transformed concentrations and proceeded in three stages: (1) unadjusted linear model with cluster-robust variance; (2) natural cubic spline (df = 3) with likelihood ratio test for nonlinearity; (3) fully adjusted model including world region, exercise mode, trial duration category, and health condition. The primary inferential comparison was between the unadjusted and fully adjusted models to quantify confounding attenuation.

Four world regions with ≥100 SBP arms (North America, Europe, East Asia, Latin America) were analysed separately with pooled models and both unadjusted and adjusted PM_2.5_ meta-regression. Five pre-specified sensitivity analyses tested robustness: (A) restriction to common covariate cells with ≥5 arms; (B) marginal standardisation with stabilised weights (capped at 5×) to equalise covariate distributions across PM_2.5_ levels; (C) PM_2.5_ × exercise mode interaction with Wald tests; (D) stratification by trial duration; and (E) progressive exclusion of high-PM_2.5_ European studies at thresholds of ≤25, ≤20, and ≤18 µg/m³. Publication bias was assessed using Egger’s regression, Begg’s rank correlation, and the trim-and-fill method.

Pre-specified subgroup analyses examined whether PM_2.5_ associations differed by exercise mode (aerobic, resistance, combined) and health condition (hypertensive, pre-hypertensive). Within each exercise mode, separate PM_2.5_ meta-regressions were fitted with log-transformed concentrations. A formal mode × PM_2.5_ interaction was tested using a likelihood ratio test comparing models with and without cross-product terms (ML estimation). For hypertensive and pre-hypertensive subgroups, PM_2.5_ was categorised into tertiles derived from the full eligible sample, and pooled effect sizes were computed within each tertile, both overall and stratified by exercise mode. Q-between tests assessed heterogeneity across tertiles. To characterise the composition of the high-pollution stratum, we additionally compared study characteristics (world region, health condition, exercise mode, baseline SBP, and BMI) across PM_2.5_ tertiles using χ² tests for categorical variables and one-way ANOVA for continuous variables; these comparisons were descriptive and post hoc.

### Risk of Bias Assessment

Risk of bias was assessed for all 465 included studies using the Cochrane RoB 2 tool for randomised trials and the ROBINS-I tool for non-randomised intervention studies. Study design was classified by examining each paper’s methods: studies that explicitly described random allocation to study groups were classified as RCTs; all others (single-group pre–post, sequential cohorts, quasi-experimental designs, etc.) were classified as non-RCTs. For RCTs, the five RoB 2 domains (randomisation process; deviations from intended interventions; missing outcome data; outcome measurement; selection of the reported result) were each judged Low, Some concerns, or High. For non-RCTs, the seven ROBINS-I domains (confounding; participant selection; intervention classification; deviations from intended interventions; missing data; outcome measurement; selection of the reported result) were each judged Low, Moderate, Serious, Critical, or No information. Missing or unclear information was conservatively rated Some concerns (RoB 2) or No information (ROBINS-I). Assessments were performed by two reviewers (JAD, AA). Both reviewers adjudicated discordant cases against the source PDFs, and consensus judgements were retained for the analysis per-study assessments and rationales are provided in Supplementary Table S16, with summary distributions in Figure S7 and per-study traffic-light judgements in Figure S8.

## Results

### Study Selection and Pooled Effects

Study and participant characteristics are summarised in Table 1. The search identified 465 included studies contributing 705 SBP arms (442 studies), 697 DBP arms (435 studies), 277 VO₂max arms (174 studies), and 327 HR arms (204 studies); the PRISMA flow diagram is presented in Figure 1.

**Table 1.**
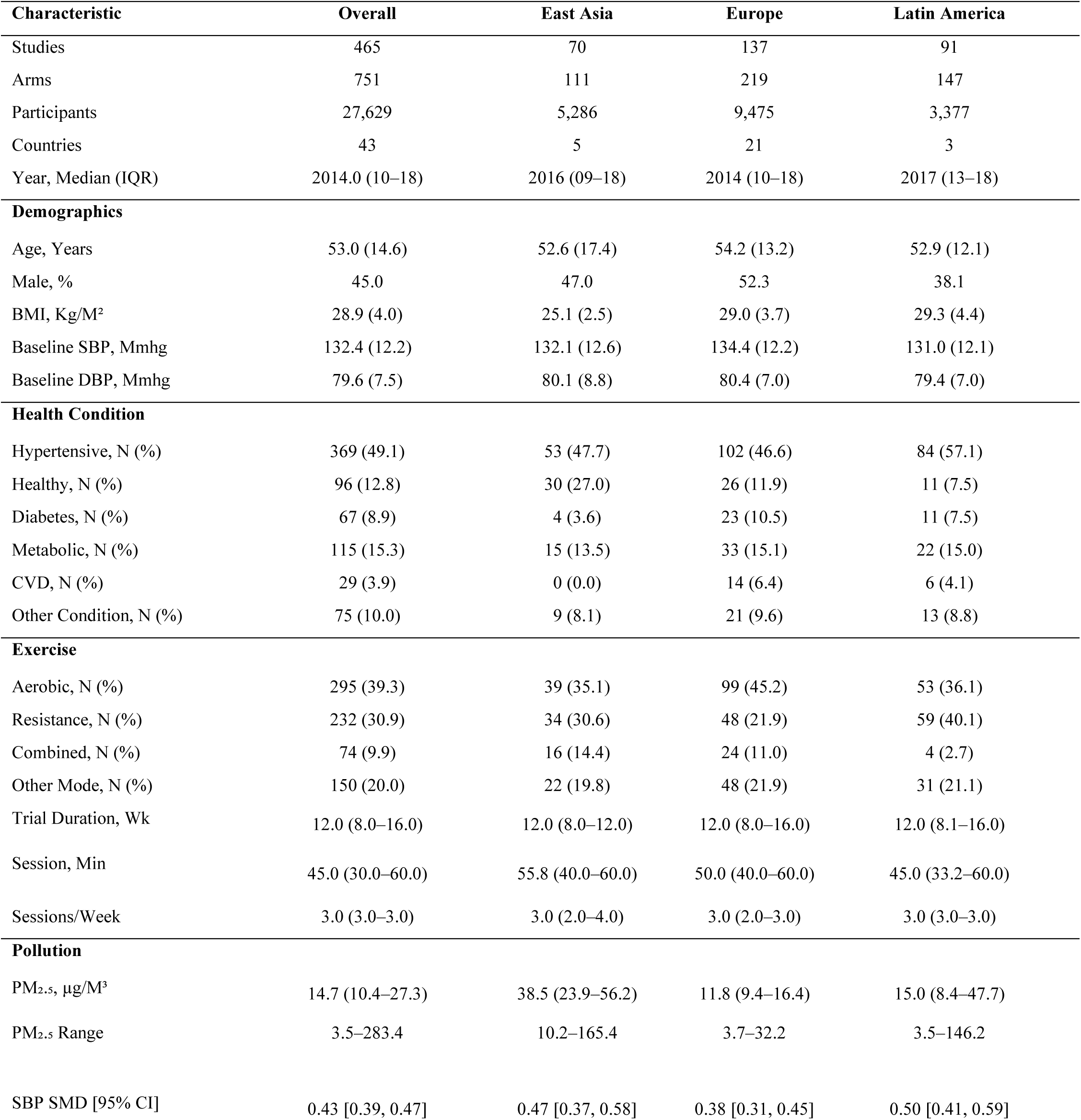
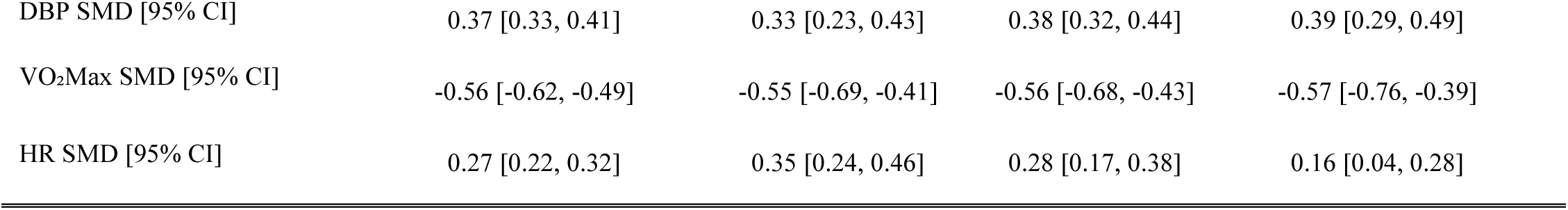
Study and participant characteristics overall, by world region, and by PM_2.5_ stratum. Continuous variables are shown as mean (SD) or median (IQR) as appropriate; categorical variables as n (%). ‘Other’ regional column groups Africa, Australasia, Middle East, South Asia, and South/Southeast Asia. PM_2.5_ strata: Low <22, Moderate 22–60, High >60 µg/m³. p-values from χ² (categorical) or Kruskal–Wallis / one-way ANOVA (continuous).

| Characteristic | Overall | East Asia | Europe | Latin America |
| --- | --- | --- | --- | --- |
| Studies | 465 | 70 | 137 | 91 |
| Arms | 751 | 111 | 219 | 147 |
| Participants | 27,629 | 5,286 | 9,475 | 3,377 |
| Countries | 43 | 5 | 21 | 3 |
| Year, Median (IQR) | 2014.0 (10–18) | 2016 (09–18) | 2014 (10–18) | 2017 (13–18) |
| **Demographics** |  |  |  |  |
| Age, Years | 53.0 (14.6) | 52.6 (17.4) | 54.2 (13.2) | 52.9 (12.1) |
| Male, % | 45.0 | 47.0 | 52.3 | 38.1 |
| BMI, Kg/M<sup>2</sup> | 28.9 (4.0) | 25.1 (2.5) | 29.0 (3.7) | 29.3 (4.4) |
| Baseline SBP, Mmhg | 132.4 (12.2) | 132.1 (12.6) | 134.4 (12.2) | 131.0 (12.1) |
| Baseline DBP, Mmhg | 79.6 (7.5) | 80.1 (8.8) | 80.4 (7.0) | 79.4 (7.0) |
| **Health Condition** |  |  |  |  |
| Hypertensive, N (%) | 369 (49.1) | 53 (47.7) | 102 (46.6) | 84 (57.1) |
| Healthy, N (%) | 96 (12.8) | 30 (27.0) | 26 (11.9) | 11 (7.5) |
| Diabetes, N (%) | 67 (8.9) | 4 (3.6) | 23 (10.5) | 11 (7.5) |
| Metabolic, N (%) | 115 (15.3) | 15 (13.5) | 33 (15.1) | 22 (15.0) |
| CVD, N (%) | 29 (3.9) | 0 (0.0) | 14 (6.4) | 6 (4.1) |
| Other Condition, N (%) | 75 (10.0) | 9 (8.1) | 21 (9.6) | 13 (8.8) |
| **Exercise** |  |  |  |  |
| Aerobic, N (%) | 295 (39.3) | 39 (35.1) | 99 (45.2) | 53 (36.1) |
| Resistance, N (%) | 232 (30.9) | 34 (30.6) | 48 (21.9) | 59 (40.1) |
| Combined, N (%) | 74 (9.9) | 16 (14.4) | 24 (11.0) | 4 (2.7) |
| Other Mode, N (%) | 150 (20.0) | 22 (19.8) | 48 (21.9) | 31 (21.1) |
| Trial Duration, Wk | 12.0 (8.0–16.0) | 12.0 (8.0–12.0) | 12.0 (8.0–16.0) | 12.0 (8.1–16.0) |
| Session, Min | 45.0 (30.0–60.0) | 55.8 (40.0–60.0) | 50.0 (40.0–60.0) | 45.0 (33.2–60.0) |
| Sessions/Week | 3.0 (3.0–3.0) | 3.0 (2.0–4.0) | 3.0 (2.0–3.0) | 3.0 (3.0–3.0) |
| **Pollution** |  |  |  |  |
| PM<sub>2.5</sub>, µg/M<sup>3</sup> | 14.7 (10.4–27.3) | 38.5 (23.9–56.2) | 11.8 (9.4–16.4) | 15.0 (8.4–47.7) |
| PM<sub>2.5</sub> Range | 3.5–283.4 | 10.2–165.4 | 3.7–32.2 | 3.5–146.2 |
| SBP SMD [95% CI] | 0.43 [0.39, 0.47] | 0.47 [0.37, 0.58] | 0.38 [0.31, 0.45] | 0.50 [0.41, 0.59] |
| DBP SMD [95% CI] | 0.37 [0.33, 0.41] | 0.33 [0.23, 0.43] | 0.38 [0.32, 0.44] | 0.39 [0.29, 0.49] |
| VO<sub>2</sub>Max SMD [95% CI] | -0.56 [-0.62, -0.49] | -0.55 [-0.69, -0.41] | -0.56 [-0.68, -0.43] | -0.57 [-0.76, -0.39] |
| HR SMD [95% CI] | 0.27 [0.22, 0.32] | 0.35 [0.24, 0.46] | 0.28 [0.17, 0.38] | 0.16 [0.04, 0.28] |

Exercise training reduced SBP by 6.1 mmHg (SMD = 0.433 [0.394, 0.472]; p < 0.001; pooled baseline SD = 14.0 mmHg), DBP by 3.3 mmHg (SMD = 0.372 [0.334, 0.410]; p < 0.001; pooled baseline SD = 8.9 mmHg), VO₂max by 3.2 ml/kg/min (SMD = −0.558 [−0.623, −0.492]; p < 0.001; pooled baseline SD = 5.8 ml/kg/min; see Table 2 footnote), and HR by 2.8 bpm (SMD = 0.267 [0.218, 0.315]; p < 0.001). Between-study variance accounted for 35.2–46.1% and within-study variance for 25.3–36.7% of total variance across outcomes. Prediction intervals were wide for all outcomes (SBP: [−0.357, 1.223]; DBP: [−0.411, 1.155]; Table 2).

**Table 2.** Pooled effects of exercise training (three-level random-effects meta-analysis). Pooled SD = root-mean-square baseline SD used for absolute unit back-transformation. Note: The VO₂max pooled SD (5.8 ml/kg/min) is lower than typical population values (∼8 ml/kg/min) because included studies used heterogeneous measurement protocols and selected participants with narrower fitness ranges; absolute VO₂max values should therefore be interpreted as conservative estimates.

| Outcome | k | Studies | SMD | 95% CI | 95% PI | Between % | Within % | Pooled SD |
| --- | --- | --- | --- | --- | --- | --- | --- | --- |
| SBP | 705 | 442 | 0.433 | [0.394, 0.472] | [-0.357, 1.223] | 46.1 | 30.9 | 14.0 |
| DBP | 697 | 435 | 0.372 | [0.334, 0.410] | [-0.411, 1.155] | 40.2 | 36.7 | 8.9 |
| VO<sub>2</sub>max | 277 | 174 | -0.558 | [-0.623, -0.492] | [-1.39, 0.274] | 40.1 | 34.3 | 5.8 |
| HR | 327 | 204 | 0.267 | [0.218, 0.315] | [-0.353, 0.886] | 35.2 | 25.3 | 10.6 |

### Pooled Effects by PM_2.5_ Stratum

Stratified by ambient PM_2.5_ (<22 / 22–60 / >60 µg/m³), SBP pooled SMDs were 0.375 [0.333, 0.418], 0.510 [0.426, 0.594], and 0.692 [0.499, 0.886] for Low, Moderate, and High strata respectively (Qₘ = 19.12, p < 0.001). DBP pooled SMDs were 0.335, 0.392, and 0.616 (Qₘ = 14.57, p < 0.001). VO₂max pooled SMDs were −0.519, −0.580, and −0.876 (Qₘ = 6.39, p = 0.041). HR showed no significant stratum difference (Qₘ = 3.70, p = 0.157). Full stratum results are presented in Table 3.

**Table 3.** Pooled effects by PM_2.5_ stratum (<22 / 22–60 / >60 µg/m³). Absolute values computed using global RMS pooled baseline SDs.

| Outcome | Stratum | k | Studies | SMD | 95% CI | p | Abs. |
| --- | --- | --- | --- | --- | --- | --- | --- |
| SBP | Low (<22) | 485 | 300 | 0.375 | (0.333, 0.418) | <0.001 | 5.3 |
| SBP | Moderate (22-60) | 170 | 108 | 0.510 | (0.426, 0.594) | <0.001 | 7.2 |
| SBP | High (>60) | 50 | 34 | 0.692 | (0.499, 0.886) | <0.001 | 9.7 |
| DBP | Low (<22) | 483 | 296 | 0.335 | (0.293, 0.377) | <0.001 | 3.0 |
| DBP | Moderate (22-60) | 166 | 105 | 0.392 | (0.302, 0.482) | <0.001 | 3.5 |
| DBP | High (>60) | 48 | 34 | 0.616 | (0.460, 0.772) | <0.001 | 5.5 |
| VO<sub>2</sub>max | Low (<22) | 185 | 119 | -0.519 | (-0.598, -0.440) | <0.001 | 3.0 |
| **VO2max** | Moderate (22-60) | 76 | 43 | -0.580 | (-0.696, -0.463) | <0.001 | 3.3 |
| **VO2max** | High (>60) | 16 | 12 | -0.876 | (-1.164, -0.589) | <0.001 | 5.0 |
| **HR** | Low (<22) | 216 | 134 | 0.250 | (0.188, 0.313) | <0.001 | 2.7 |
| **HR** | Moderate (22-60) | 90 | 58 | 0.266 | (0.192, 0.339) | <0.001 | 2.8 |
| **HR** | High (>60) | 21 | 12 | 0.420 | (0.190, 0.651) | 0.005 | 4.5 |

### Systematic Review of Regional Distribution and Study Characteristics

The 465 included studies came from 301 distinct study locations across 43 countries (Table S17). Low-pollution studies were predominantly from North America and Europe; high-pollution studies predominantly from East Asia and Latin America. This geographic–pollution co-distribution means that comparing across PM_2.5_ strata also compares across regions that differ systematically in how structured exercise is delivered and in their patient populations.

Regional variation in pooled SBP outcomes was substantial (all absolute values computed using the global RMS pooled baseline SD of 14.0 mmHg). Latin American studies showed the largest reductions (SMD = 0.503, 7.1 mmHg; k = 136, 86 studies). East Asian studies: SMD = 0.473, 6.6 mmHg (k = 107, 69 studies). European studies: SMD = 0.383, 5.4 mmHg (k = 200, 125 studies). North American studies showed the smallest (SMD = 0.353, 4.9 mmHg; k = 192, 117 studies). These regional differences are comparable in magnitude to the PM_2.5_ stratum differences reported above (Table S3).

Study characteristics differed systematically across regions (Table 4). Latin American trials had the highest proportion of hypertensive patients (55.1%), while Europe had the highest weighted mean baseline SBP (134.4 mmHg) and highest proportion of aerobic protocols (54.8%). East Asian trials had the highest mean PM_2.5_ (51 µg/m³), a level in the upper range of the Moderate stratum (22–60 µg/m³). Latin American trials had the shortest mean duration (13.1 weeks). North American trials were the most heterogeneous in exercise mode. Across the four viable regions, the proportion of hypertensive participants correlated strongly with the pooled SBP effect size (r = 0.836, n = 4 regions; Figure 3B), connecting regional composition to outcome variation. This correlation, while based on only four datapoints and therefore not amenable to formal inference, is consistent with the known dose–response relationship between baseline blood pressure and exercise-induced reduction.

**Figure 3.**
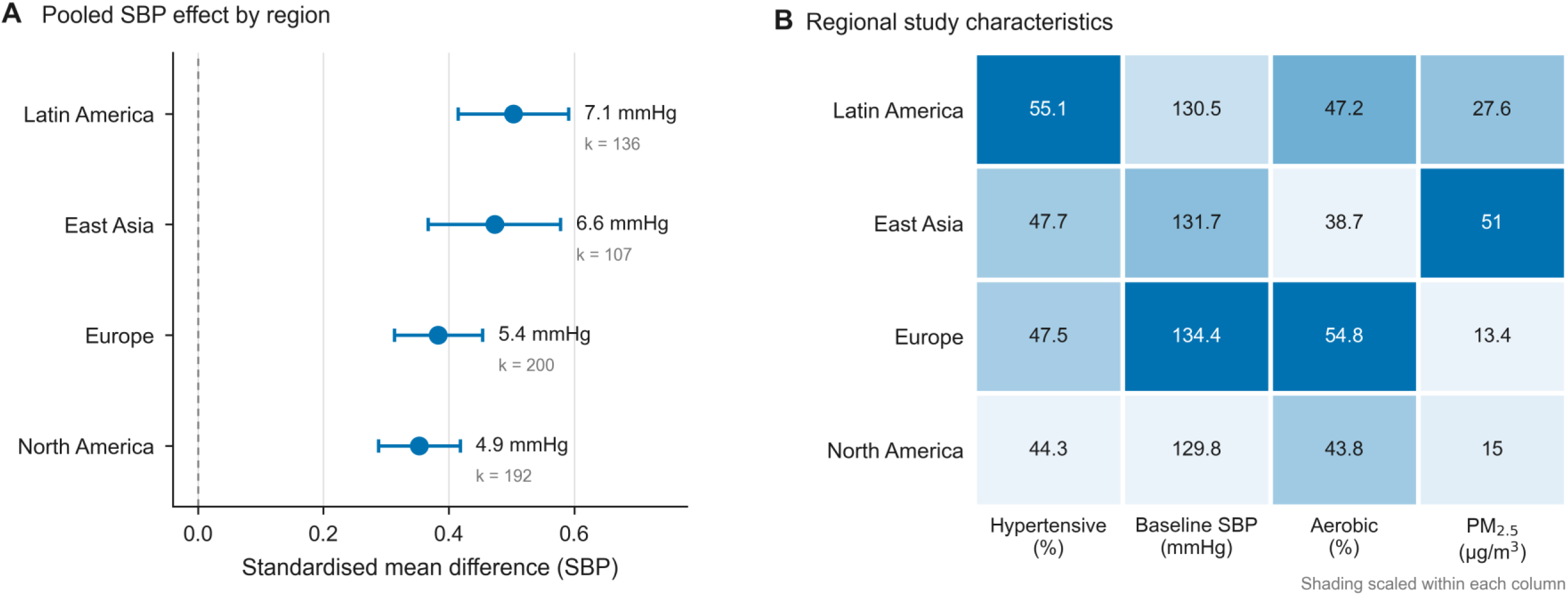
Regional pooled SBP effects (A) and characteristics (B) heatmap.

**Table 4.** Regional study characteristics.

| **Region** | **Studies** | **Arms** | **% HTN** | **Mean SBP<sub>0</sub>** | **% Aerobic** | **Dur (wk)** | **PM<sub>2.5</sub>** | **PM<sub>2.5</sub> Range** |
| --- | --- | --- | --- | --- | --- | --- | --- | --- |
| **East Asia** | 69 | 107 | 47.7 | 131.7 | 38.7 | 13.5 | 51.0 | 10.2-165.4 |
| **Europe** | 125 | 200 | 47.5 | 134.4 | 54.8 | 14.2 | 13.4 | 3.7-32.2 |
| **Latin America** | 86 | 136 | 55.1 | 130.5 | 47.2 | 13.1 | 27.6 | 5.8-146.2 |
| **North America** | 117 | 192 | 44.3 | 129.8 | 43.8 | 15.3 | 15.0 | 3.6-34.7 |

### PM_2.5_ Meta-regression

In global meta-regression, the unadjusted log(PM_2.5_) coefficient was positive and significant for SBP (β = +0.0948, p < 0.001; k = 705), DBP (β = +0.0702, p = 0.007), and HR (β = +0.0588, p = 0.044), and negative for VO₂max (β = −0.0971, p = 0.031). After adjustment for region, exercise mode, trial duration, and health condition (k = 574 for SBP), the SBP coefficient attenuated by 70% to non-significance (β = +0.0282, p = 0.450). DBP attenuated to non-significance (β = +0.0596, p = 0.107). VO₂max attenuated substantially (β = −0.0157, p = 0.827). HR remained non-significant (β = +0.0465, p = 0.283) (Table 5). Restricted cubic spline models detected no significant nonlinearity for any outcome (SBP LRT p = 0.353; DBP p = 0.423; VO₂max p = 0.685; HR p = 0.627) (Table S2).

**Table 5.** Global PM_2.5_ meta-regression. Adjusted for region, exercise mode, duration, and health condition.

| **Outcome** | **k** | **Studies** | **β unadj** | **p unadj** | **β adj** | **p adj** | **Atten %** |
| --- | --- | --- | --- | --- | --- | --- | --- |
| **SBP** | 705 | 442 | 0.0948 | <0.001 | 0.0282 | 0.450 | 70 |
| **DBP** | 697 | 435 | 0.0702 | 0.0066 | 0.0596 | 0.107 | 15 |
| **VO2max** | 277 | 174 | -0.0971 | 0.0307 | -0.0157 | 0.827 | 84 |
| **HR** | 327 | 204 | 0.0588 | 0.0437 | 0.0465 | 0.283 | 21 |

Within-region adjusted PM_2.5_ coefficients were non-significant for SBP in all four regions (Table 6). The European DBP coefficient reached significance after adjustment (adjusted β = +0.2151, p = 0.028; k = 156), the only within-region coefficient to achieve significance in the adjusted models.

**Table 6.** Within-region PM_2.5_ coefficients. k denotes the adjusted-model sample size; unadjusted coefficients were fitted on all arms with PM_2.5_ data in each region (Table S11).

| **Outcome** | **Region** | **k** | **β unadj** | **p unadj** | **β adj** | **p adj** |
| --- | --- | --- | --- | --- | --- | --- |
| **SBP** | East Asia | 93 | 0.1879 | 0.032 | 0.0339 | 0.735 |
| **SBP** | Europe | 157 | 0.0658 | 0.388 | 0.1353 | 0.126 |
| **SBP** | Latin America | 107 | 0.0123 | 0.805 | -0.0142 | 0.780 |
| **SBP** | North America | 162 | 0.0377 | 0.594 | 0.0902 | 0.214 |
| **DBP** | East Asia | 91 | 0.1355 | 0.064 | 0.1118 | 0.205 |
| **DBP** | Europe | 156 | 0.1498 | 0.063 | 0.2151 | 0.028 |
| **DBP** | Latin America | 101 | 0.0378 | 0.528 | 0.0132 | 0.848 |
| **DBP** | North America | 164 | 0.0083 | 0.905 | 0.0410 | 0.542 |
| **VO2max** | East Asia | 27 | -0.0093 | 0.919 | 0.0311 | 0.824 |
| **VO2max** | Europe | 74 | -0.1940 | 0.242 | -0.2138 | 0.300 |
| **VO2max** | Latin America | 41 | -0.0395 | 0.780 | 0.1279 | 0.468 |
| **VO2max** | North America | 65 | -0.0738 | 0.287 | -0.1016 | 0.417 |
| **HR** | East Asia | 42 | 0.1416 | 0.106 | 0.1962 | 0.114 |
| **HR** | Europe | 65 | -0.0327 | 0.703 | -0.0936 | 0.548 |
| **HR** | Latin America | 47 | 0.0491 | 0.526 | 0.0185 | 0.711 |
| **HR** | North America | 73 | 0.0595 | 0.352 | 0.0331 | 0.767 |

**Table 7.** Clinical translation by PM_2.5_ stratum. Absolute changes computed using global RMS pooled baseline SDs (SBP 14.0 mmHg, DBP 8.9 mmHg, VO₂max 5.8 ml/kg/min). Stroke RRR derived from Law et al. (2009); broader cardiovascular risk reductions per 10 mmHg SBP (major cardiovascular events ∼20%, coronary heart disease ∼17%, stroke ∼27%, heart failure ∼28%) from Ettehad et al. (2016); VO₂max all-cause mortality RRR from Kodama et al. (2009).

| **Outcome** | **Stratum** | **k** | **SMD** | **Abs.**<br>**change** | **Units** | **RRR %** | **RRR type** |
| --- | --- | --- | --- | --- | --- | --- | --- |
| **SBP** | Low | 485 | 0.375 | 5.2 | mmHg | 16.2 | Stroke |
| **SBP** | Moderate | 170 | 0.510 | 7.2 | mmHg | 21.3 | Stroke |
| **SBP** | High | 50 | 0.692 | 9.7 | mmHg | 27.8 | Stroke |
| **DBP** | Low | 483 | 0.335 | 3.0 | mmHg | 7.8 | Stroke |
| **DBP** | Moderate | 166 | 0.392 | 3.5 | mmHg | 9.1 | Stroke |
| **DBP** | High | 48 | 0.616 | 5.5 | mmHg | 13.9 | Stroke |
| **VO2max** | Low | 185 | -0.519 | 3.0 | ml/kg/min | 10.2 | All-cause mortality |
| **VO2max** | Moderate | 76 | -0.580 | 3.4 | ml/kg/min | 11.3 | All-cause mortality |
| **VO2max** | High | 16 | -0.876 | 5.1 | ml/kg/min | 16.6 | All-cause mortality |

North American DBP showed a non-significant adjusted PM_2.5_ coefficient (β = +0.0410, p = 0.542; k = 164, 107 studies). East Asian SBP showed the largest unadjusted regional coefficient (β = +0.1879, p = 0.032; k = 107) but this attenuated to non-significance after adjustment (β = +0.0339, p = 0.736; k = 93). (Table S11).

The duration × PM_2.5_ interaction was non-significant for all outcomes: SBP (LRT χ² = 0.091, p = 0.955), DBP (LRT χ² = 2.273, p = 0.321), VO₂max (LRT χ² = 3.513, p = 0.173), and HR (LRT χ² = 1.829, p = 0.401). (Table S7). Pre-specified sensitivity analyses; 1) restriction to common covariate cells (Table S4), 2) marginal standardisation across PM_2.5_ levels (Table S5), 3) PM_2.5_ × exercise mode Wald tests (Table S6) and 4) a finer four-strata cut of PM_2.5_ (Table S9) yielded directionally consistent results.

**Figure 4.**
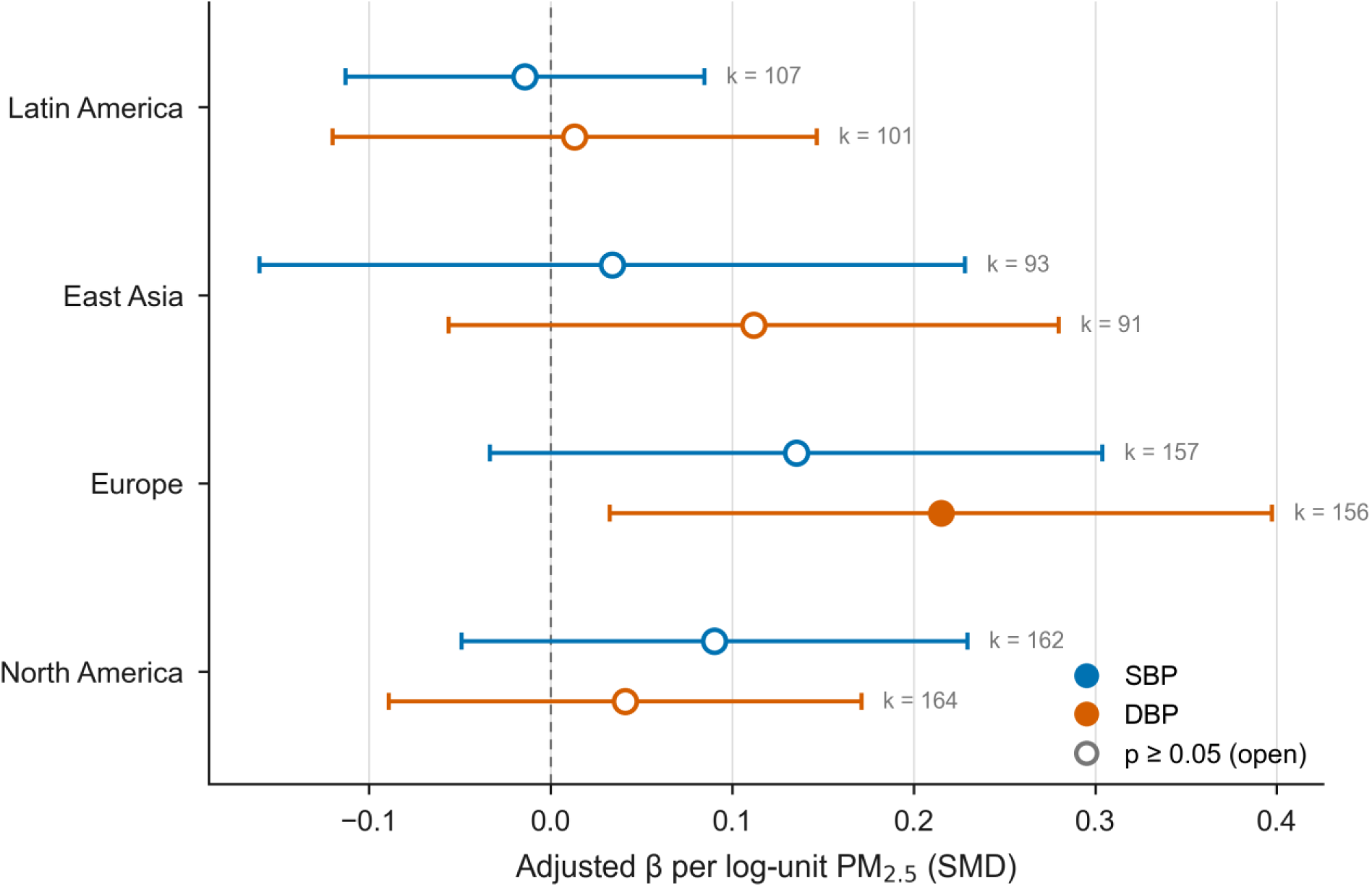
Regional adjusted PM_2.5_ coefficients.

### Subgroup Analyses

Pre-specified mode-stratified PM_2.5_ meta-regression showed a significant positive association for aerobic exercise arms (k = 273; SBP β = +0.1527, p = 0.003; DBP β = +0.0930, p = 0.017) but no association for resistance training arms (k = 219; SBP β = +0.0154, p = 0.666; DBP β = +0.0376, p = 0.341). The mode × PM_2.5_ interaction approached significance for SBP (LRT χ² = 3.18, p = 0.074). (Table S13).

Among hypertensive participants, aerobic exercise SBP pooled SMDs increased monotonically across PM_2.5_ strata (<22 / 22–60 / >60 µg/m³): 0.396 [k = 77], 0.751 [k = 30], and 1.020 [k = 11] (Qₘ p < 0.001; Figure 5), representing a 2.6-fold gradient. Resistance training showed no systematic gradient: 0.519 [k = 59], 0.618 [k = 22], 0.735 [k = 5] (Qₘ p = 0.551). DBP findings were consistent: aerobic 0.404, 0.610, 0.832; resistance 0.464, 0.521, 0.648.

**Figure 5.**
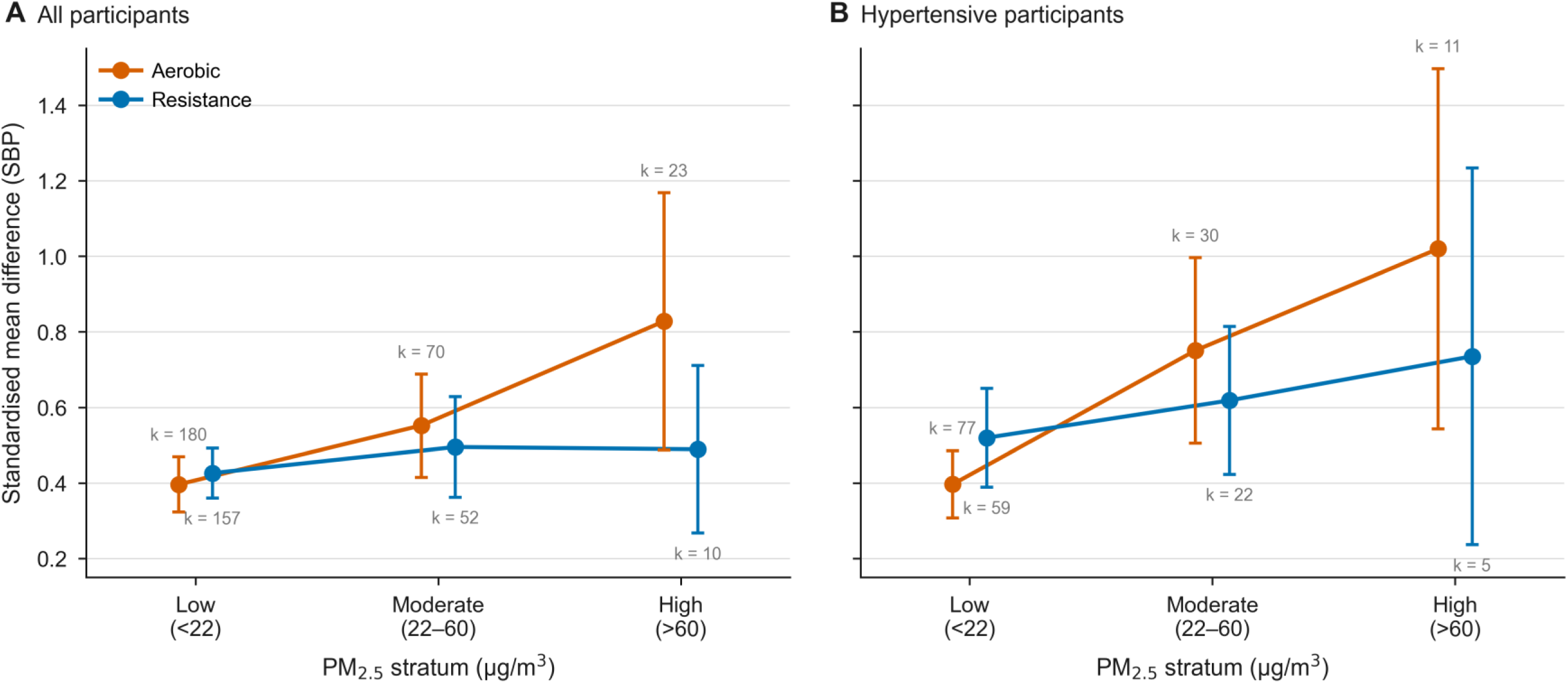
Aerobic vs resistance SBP SMD and 95% CI by PM_2.5_ stratum - all participants (A) and hypertensive subgroup (B).

For VO₂max, aerobic exercise pooled SMDs in hypertensive participants were −0.599 [-0.758, −0.440] (Low; k = 32), −0.760 [-0.967, −0.553] (Moderate; k = 18), and −1.105 [-1.599, −0.610] (High; k = 5). Resistance training VO₂max data were available for the Low stratum only (SMD = −0.413 [-0.622, −0.204]; k = 10). For resting HR, aerobic exercise pooled SMDs were 0.321 (Low; k = 26), 0.367 (Moderate; k = 15), and 0.549 (High; k = 6); resistance training SMDs were 0.289 (Low; k = 24), 0.020 (Moderate; k = 9).

### Composition of the High-Pollution Stratum

To characterise the studies driving the larger pooled effect sizes at higher pollution levels, we examined the compositional differences across PM_2.5_ tertiles (T1 <11.6, T2 11.6–19.8, T3 >19.8 µg/m³; k = 229, 237, and 239 SBP arms respectively). Regional composition differed markedly (χ² test, p < 0.001): T3 studies were predominantly from East Asia (35.1%), Latin America (23.8%), and North America (14.6%), whereas T1 studies were predominantly European (40.6%) and North American (25.3%). Only 0.4% of T1 arms originated from East Asia compared with 35.1% in T3.

Health condition composition also differed across tertiles (χ², p = 0.028). T3 studies had a slightly lower proportion of hypertensive participants (46.9%) compared with T2 (55.3%) and T1 (44.5%). Exercise mode composition differed significantly (χ², p = 0.010): combined exercise was more prevalent in T3 (13.8% vs 8.7% in T1), while mind-body exercise appeared almost exclusively in T3 (4.2% vs 0.4%). Mean baseline SBP was similar across tertiles (T1: 132.2, T2: 131.6, T3: 133.3 mmHg; one-way ANOVA, p = 0.274). Mean BMI was lower in T3 (27.6 kg/m²) than T1 (29.5 kg/m²). Full moderator-stratified estimates by PM_2.5_ tertile are reported in Table S10.

Within T3, the largest national contributions came from Brazil (k = 52; mean PM_2.5_ = 46.3; mean yi = 0.540), Japan (k = 27; 34.5 µg/m³; yi = 0.340), and the United States (k = 23; 24.6 µg/m³; yi = 0.484). The largest mean effect sizes were observed in Nigeria (k = 9; 69.8 µg/m³; yi = 1.207; 100% hypertensive), China (k = 10; 105.2 µg/m³; yi = 1.025; 50% hypertensive), and Iran (k = 6; 27.3 µg/m³; yi = 0.956; 100% hypertensive).

Sequential covariate adjustment in multilevel meta-regression quantified the contribution of each factor to the T3 excess. The unadjusted T3 vs T1 contrast was β = +0.132 (p = 0.006). Adjusting for health condition attenuated this by 0.6% (β = +0.131, p = 0.006). Adding region reduced the contrast by 35.9% (β = +0.085, p = 0.140). Further adjustment for exercise mode attenuated the contrast by 63.4% (β = +0.048, p = 0.437), and the addition of trial duration and baseline SBP yielded a final attenuation of 79.7% (β = +0.027, p = 0.647). Thus, the compositional differences in region and exercise mode accounted for the majority of the apparent T3 excess in SBP effect size (Table S8).

Table S8. Sequential covariate adjustment for the T3 vs T1 SBP effect size contrast.

### Cardiorespiratory Fitness

VO₂max improvements increased across PM_2.5_ strata (Qₘ = 6.39, p = 0.041; Table 3; Figure 6), mirroring the blood pressure findings; the High stratum estimate is based on only 16 arms.

**Figure 6.**
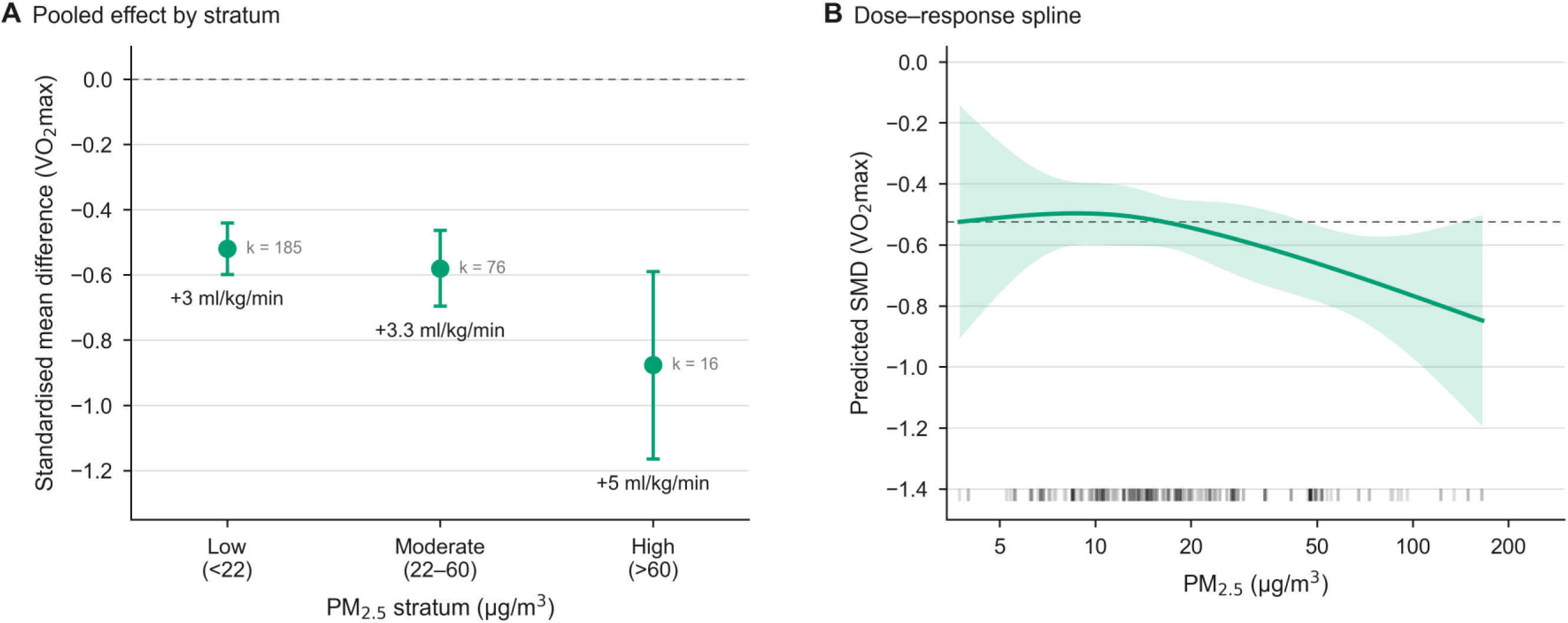
SMD of VO₂max by PM_2.5_ stratum (A) and dose-response spline (B). Note: Negative values indicate positive pre-post difference i.e. increase in VO_2Max_ following the intervention.

### Sensitivity Analysis: Isometric Exercise Training (IET) Classification

Given the large magnitude of blood pressure reductions reported for isometric exercise training (IET) relative to other exercise modalities (Edwards et al. 2022; Edwards et al. 2023), we conducted a sensitivity analysis examining the impact of IET classification on resistance training subgroup results. In the primary analysis, IET studies (isometric handgrip and wall squat protocols; k = 19 arms, 15 studies) were classified as resistance training. The IET-only subgroup showed a substantially larger pooled SBP effect (SMD = 0.671; 95% CI: 0.414, 0.928) compared to conventional dynamic resistance training alone (SMD = 0.428; 95% CI: 0.369, 0.486). For DBP, IET showed a pooled SMD of 0.459 (95% CI: 0.263, 0.655) versus 0.377 (95% CI: 0.317, 0.437) for conventional resistance training. Including IET in the resistance category modestly increased the pooled resistance SBP estimate from 0.428 to 0.446.

Critically, PM_2.5_ meta-regression coefficients were materially unchanged by IET classification (SBP β = +0.0154, p = 0.666 with IET; β = +0.019, p = 0.61 without IET), indicating that the primary pollution– exercise findings are robust to this classification decision.

### Publication Bias

Egger’s regression indicated funnel asymmetry for all four outcomes: SBP (z = 12, p < 0.001), DBP (z = 9.72, p < 0.001), VO₂max (z = −6.5, p < 0.001), and HR (z = 4.43, p < 0.001). Begg’s rank correlation was significant for SBP (τ = 0.242, p < 0.001), DBP (τ = 0.207, p < 0.001), and VO₂max (τ = −0.216, p < 0.001), with HR approaching significance (τ = 0.102, p = 0.030). Trim-and-fill imputed zero missing studies for all four outcomes; adjusted pooled estimates were unchanged (SBP: 0.432; DBP: 0.367; VO₂max: −0.555; HR: 0.265). (Table S15).

### Risk of Bias

Of the 465 included studies, 316 (68%) were classified as randomised trials and 149 (32%) as non-randomised intervention studies. Among RCTs, the distribution of overall RoB 2 judgements was Low 32 (10.1%), Some concerns 240 (75.9%), and High 44 (13.9%). The predominance of Some concerns reflected incomplete reporting of allocation concealment (D1: 56.3% Some concerns) and the inherent unblinding of participants and personnel in exercise interventions (D2: 73.4% Some concerns). Outcome measurement (D4) was the strongest RCT domain, with 65.2% rated Low risk, reflecting widespread use of validated automated blood-pressure devices and standardised cardiopulmonary testing. Among non-RCT designs, residual confounding intrinsic to non-randomised allocation rendered every study at least Moderate risk on ROBINS-I (Moderate 13.4%; Serious 86.6%), with the confounding domain (D1) uniformly Serious (85.9%) or Moderate (13.4%). Summary domain-level distributions and per-study traffic-light judgements are presented in Figures S7 and S8.

## Discussion

### Main Findings

This meta-analysis of 465 studies spanning a >100-fold ambient PM_2.5_ gradient demonstrates that exercise training produces large, clinically meaningful cardiovascular benefits regardless of pollution level or world region. The pooled SBP reduction of 6.1 mmHg and DBP reduction of 3.3 mmHg exceed established minimal clinically important differences (MCIDs) for blood pressure outcomes, confirming clinical relevance. These benefits were observed consistently across all four major world regions and every PM_2.5_ stratum studied. Cardiorespiratory fitness showed the same pattern; resting heart rate improved significantly overall, although its stratum gradient did not reach statistical significance. Across all outcomes, benefits were if anything larger in higher-pollution settings, which likely reflects the higher cardiovascular risk burden of populations in those settings. Across exercise modes, aerobic training delivered the largest overall gains, while resistance training produced consistent blood pressure reductions at every PM_2.5_ level.

Regional variation in outcomes tracks directly with population characteristics. East Asian and Latin American trials, which contributed the majority of high-pollution studies, enrolled higher proportions of hypertensive participants with higher baseline blood pressures and greater capacity for improvement.

Across the four major world regions, hypertension prevalence correlated strongly with pooled SBP effect size. This is consistent with the well-established relationship between baseline blood pressure severity and the magnitude of exercise-induced reduction (Cornelissen & Smart 2013).

Hypertension is the key effect modifier, and the mode-stratified hypertensive findings provide the clearest evidence about what drives the outcome gradient. Among hypertensive participants, aerobic exercise showed a 2.6-fold SBP effect-size gradient from the Low to the High PM_2.5_ stratum, whereas resistance training showed no gradient (Figure 5). Both groups experienced similar chronic ambient PM_2.5_ exposure throughout their trial periods. The gradient is specific to aerobic exercise in hypertensive populations, which is precisely where the concentration of high-burden patients in high-pollution settings is greatest in our data. Covariate adjustment attenuated the global SBP coefficient by 70% to non-significance, consistent with this interpretation. It should be acknowledged that these mode × stratum comparisons are based on between-trial differences and that residual confounding, particularly in baseline blood pressure severity within hypertensive subgroups, cannot be excluded.

Several non-mutually exclusive mechanisms may explain the larger observed improvements in higher-pollution settings. Populations from these regions often had greater baseline cardiovascular risk, including higher hypertension prevalence, lower baseline fitness, and potentially greater physical deconditioning, all of which increase the capacity for absolute physiological improvement following structured exercise training. Differences in healthcare access, medication optimisation, supervision intensity, and behavioural change associated with trial participation may also contribute. Additionally, regional variation in study design and participant selection could introduce residual ecological confounding despite multivariable adjustment. Collectively, these factors likely explain much of the apparent pollution gradient without requiring a biologically beneficial effect of PM_2.5_ exposure itself.

Cardiorespiratory fitness showed the same directional pattern as blood pressure, with larger VO₂max improvements in higher-pollution strata. These findings reflect larger fitness gains in higher-risk, more deconditioned populations undertaking aerobic exercise, consistent with the blood pressure findings rather than diverging from them. The High stratum contained only 16 arms, and the VO₂max gradient should be interpreted with caution.

Our findings diverge from large observational cohort studies reporting that PM_2.5_ attenuates the mortality benefits of habitual physical activity. Ku et al. (2025) and Guo et al. (2020) identified attenuation of physical activity benefits above 25 µg/m³ in cohort data. Several features of the present analysis may explain this divergence. Structured supervised exercise trials deliver a controlled dose of exercise to defined patient populations; observational cohorts capture self-selected habitual activity across heterogeneous populations where who chooses to exercise more, and at what intensity, co-varies with socioeconomic and environmental factors that cannot be fully adjusted for. The baseline risk burden that drives larger benefits in high-pollution trial populations, where participants presented with higher hypertension prevalence and lower baseline fitness, is not a controllable quantity in cohort analyses of habitual activity. Additionally, cohort mortality endpoints integrate decades of cumulative exposure and competing risks that 4–52 week intervention trials are not designed to capture.

The isometric exercise training (IET) effect sizes observed here are consistent with the large blood pressure reductions reported in prior meta-analyses of isometric training (Edwards et al. 2022) and with the finding from a large-scale network meta-analysis that isometric exercise ranked as the most effective exercise modality for SBP reduction (SUCRA 98.3%; Edwards et al. 2023). Our sensitivity analysis indicates that the primary pollution–exercise findings are robust to how IET is classified.

### Clinical and Public Health Implications

These findings carry a clear public health message: exercise training delivers clinically meaningful cardiovascular benefits across the full pollution gradient studied, with pooled SBP and DBP reductions of approximately 6.1 and 3.3 mmHg exceeding established MCIDs (typically 2–5 mmHg for SBP and 2–3 mmHg for DBP; Pescatello et al. 2024) at every PM_2.5_ level tested (Tables 7 and S14). Applying the SBP-lowering risk coefficients of Ettehad et al. (2016), the pooled reduction corresponds to an estimated ∼12% reduction in major cardiovascular events, ∼16% in stroke, and ∼17% in heart failure, with proportionally larger reductions in the High stratum (9.7 mmHg SBP reduction), and the VO₂max gains observed here translate into an estimated 11% reduction in all-cause mortality (Kodama et al. 2009). Dual interventions combining exercise promotion with air quality improvement remain sensible on public health grounds and are supported by evidence that acute pollution exposure can transiently impair cardiovascular function (Sinharay et al. 2018; Rajagopalan et al. 2018); the present trial evidence does not, however, suggest that pollution reduction is necessary to realise exercise-induced blood pressure benefits.

Hypertensive patients represent the subgroup with the most to gain: across all PM_2.5_ strata they showed the largest SBP and DBP improvements in the dataset, and the concentration of high-risk patients in polluted settings means the absolute benefit of prescription is greatest exactly where air quality concerns are most acute. For clinicians, these data support continuing all forms of exercise prescription without modification based on ambient PM_2.5_ levels. Aerobic training delivered the greatest overall health gains, particularly for hypertensive patients and cardiorespiratory fitness, and retained a clear benefit even in the highest-pollution settings. Resistance training produced consistent SBP and DBP reductions across all strata with no PM_2.5_ gradient (High-stratum estimates based on k = 10 warrant caution), making it an especially reassuring option where air quality concerns exist. Combined modalities (concurrent aerobic and resistance training) were not analysed separately owing to small cell sizes but were included in pooled estimates.

### Strengths and Limitations

This study has several methodological strengths. It represents the largest meta-analysis to address the pollution–exercise interaction question, drawing on 465 studies (27,629 participants) across 43 countries, with 705 SBP arms from 442 studies contributing to the primary blood-pressure analysis. The three-level random-effects model correctly accounts for arm-within-study nesting, and CR2 cluster-robust standard errors protect against misspecified correlations. PM_2.5_ was objectively assigned from a unified global reanalysis (CAMS ERA5). Five pre-specified sensitivity analyses provided rigorous testing across multiple analytical approaches. The resistance exercise natural experiment represents a novel methodological approach: no previous exercise–pollution meta-analysis has used within-trial mode comparison to control for chronic ambient exposure and examine whether the PM_2.5_ gradient differs by exercise mode. Sensitivity analysis varying the assumed pre-post correlation from r = 0.50 to r = 0.85 confirmed that pooled estimates were virtually unchanged (maximum 1.7% variation across outcomes; Table S12).

Several limitations warrant consideration. Risk of bias was assessed for all 465 included studies using Cochrane RoB 2 (for the 316 RCTs) and ROBINS-I (for the 149 non-randomised studies). Across RCTs, 32 studies (10.1%) were rated Low overall risk, 240 (75.9%) Some concerns, and 44 (13.9%) High; the predominance of “Some concerns” was driven principally by incomplete reporting of allocation concealment and the inherent unblinding of participants in exercise interventions, both of which are well-recognised features of the exercise-training literature. Across non-RCT designs, residual confounding intrinsic to non-randomised allocation rendered every study at least Moderate risk on ROBINS-I - an unavoidable consequence of design rather than a quality issue specific to the present corpus. The consistency of the pooled exercise effect across all bias strata supports that overall bias is unlikely to materially alter the principal findings (Figures S7–S8).

Additional limitations include: (i) ambient PM_2.5_ is an ecological measure that does not reflect personal exposure during exercise, and indoor training, which comprised the majority of interventions, substantially reduces effective exposure; (ii) monthly PM_2.5_ means were matched to trial period but remain grid-cell averages (≈80 km resolution) and therefore cannot capture acute or session-level exposure variation, introducing measurement error biased toward the null; (iii) residual confounding within health condition categories (particularly hypertension severity) limits the precision of adjusted analyses as we were unable to directly test whether baseline blood pressure severity co-varies with PM_2.5_ within aerobic trials; (iv) VO₂max analyses were underpowered in the High PM_2.5_ stratum (16 arms); (v) the three-stratum PM_2.5_ categorisation (<22 / 22–60 / >60 µg/m³) was data-driven (AIC-optimised) rather than pre-specified, which should be acknowledged as a potential analytical flexibility concern, although the meta-regression results are based on continuous log(PM_2.5_); (vi) the cross-sectional exposure assignment cannot account for within-trial pollution variation; and (vii) cell sizes were small for some mode–stratum combinations (particularly resistance High stratum, k = 10), limiting statistical power.

Additionally, although all included studies required stable medication regimens throughout the trial, participant medication status varied considerably: some studies enrolled unmedicated participants, others required stable antihypertensive therapy, and many did not report medication status. Antihypertensive medication may attenuate (through ceiling effects on blood pressure reduction) or interact with exercise-induced adaptations, and we were unable to systematically account for this heterogeneity.

## Conclusions

In this meta-analysis of 465 studies spanning a >100-fold global PM_2.5_ gradient, exercise training consistently produced large, clinically meaningful cardiovascular benefits regardless of ambient pollution level. Benefits were larger in higher-pollution settings across all four outcomes, reflecting the greater cardiovascular risk burden of those populations rather than a pollution–exercise interaction; hypertensive patients stand to gain the most. Resistance training in particular delivered consistent benefits independent of ambient air quality across the full pollution range studied. These findings should reassure clinicians and patients in polluted settings that the cardiovascular benefits of exercise are preserved. Future studies should incorporate personal exposure monitoring during exercise and session-level pollution data to better characterise the conditions under which these benefits are realised.

## Supporting information

PRISMA 2020 Checklist

ROB Assessments

City Level Results

## Acknowledgments

This systematic review is supported by the National Institute for Health and Care Research (NIHR) Leicester Biomedical Research Centre (BRC). The NIHR Leicester BRC is a partnership between the University of Leicester, University Hospitals of Leicester NHS Trust, Loughborough University, and the University Hospitals of Northamptonshire NHS Group. The views expressed are those of the authors and not necessarily those of the NIHR or the Department of Health and Social Care.

## Data Availability

The analysis dataset and R code are publicly available to view in an R Shiny web application at https://8i8hck-jamesad90.shinyapps.io/shiny_map_app/. PM_2.5_ exposure data were derived from the publicly available CAMS ERA5 reanalysis (https://atmosphere.copernicus.eu/).

## Supplementary Material

Figure S1. Exercise mode-specific PM_2.5_ slopes by region for SBP (Sensitivity C).

Figure S2. Standard and contour-enhanced funnel plots for SBP, DBP, VO₂max, and HR. Panels are provided as sequential pages: standard funnel plots (pages 1–4) and contour-enhanced funnel plots (pages 5–8), each ordered SBP, DBP, VO₂max, HR.

Figure S3. Bubble plots of PM_2.5_ versus effect size for DBP, VO₂max, and HR (one outcome per page).

Figure S4. Forest plot of exercise training effects on systolic blood pressure.

Figure S5. Forest plots for DBP, VO₂max, and HR (one outcome per page).

Figure S6. Dose-response curves for DBP, VO₂max, and HR (one outcome per page).

Figure S7. Risk of bias summary. A) RoB 2 stacked bar chart for randomised trials (n = 316). B) ROBINS-I stacked bar chart for non-randomised intervention studies (n = 149).

Figure S8. Per-study risk-of-bias traffic-light judgements. A) RoB 2 (n = 316). B) ROBINS-I (n = 149).

Table S1. Sensitivity analysis: effect of outlier inclusion on pooled SBP estimate.

Table S2. Spline nonlinearity tests (likelihood ratio tests).

Table S3. Regional pooled effects for all four outcomes.

Table S4. Sensitivity A - common covariate cells: adjusted PM_2.5_ coefficients.

Table S5. Sensitivity B - marginal standardisation: adjusted PM_2.5_ coefficients.

Table S6. Sensitivity C - Wald tests and mode-specific slopes.

Table S7. Sensitivity D - duration-stratified PM_2.5_ coefficients.

Table S15. Publication bias assessment (Egger, Begg, trim-and-fill).

Table S9. Four-strata pooled effects by PM_2.5_ level.

Table S10. Moderator-stratified analysis: SBP and VO₂max by PM_2.5_ tertile.

Table S11. Regional unadjusted PM_2.5_ coefficients with exposure characteristics.

Table S12. Sensitivity analysis: effect of assumed pre-post correlation (r = 0.50, 0.70, 0.85) on pooled estimates.

Table S13. Subgroup analyses: exercise mode pooled effects, PM_2.5_ meta-regression within each mode, and exercise mode × PM_2.5_ interaction tests.

Table S14. Clinical translation: standardised mean differences converted to absolute units (mmHg, ml/kg/min) with estimated cardiovascular risk reductions by PM_2.5_ stratum.

Table S16. Per-study risk-of-bias assessments (RoB 2 / ROBINS-I) with domain-level judgements and rationales for all 465 included studies. Provided as separate CSV file: rob_assessments.csv.

Table S17. City-level results for all 301 distinct study locations: city, country, region, coordinates, numbers of studies, arms, and participants, mean PM_2.5_, and pooled SBP and DBP effect sizes with 95% CIs (REML random-effects within location; single-arm locations report the arm-level estimate). Multi-site studies are counted at each contributing location. Provided as separate CSV file: TableS17_city_level_results.csv.

Appendix S1. Supplementary Statistical Methods: model equations and estimation details.

## Declarations

### Author Contributions

JAD, GAN and JOD conceived the study, developed the web-scraping tools, conducted the data collection and statistical analyses, and drafted the manuscript. SC, JVH, and AH provided expertise in environmental health and epidemiological methods, contributed to study design and interpretation, and critically revised the manuscript. JVH contributed environmental exposure assessment expertise. TY, AA, RP, and GAN contributed to interpretation and critically revised the manuscript. JE screened studies as a reviewer and contributed to refinement of the study design and statistical approach. All authors read and approved the final version. JAD is the guarantor of this work, had full access to all data in the study, and takes responsibility for the integrity of the data and accuracy of the analysis.

