## Supplementary Materials for "Ambient PM_2.5_ Concentration and the Cardiovascular Response to Exercise Training: A Systematic Review and Meta-Analysis Across Global Pollution Gradients"

PROSPERO registration: CRD420251068843

**3 Appendix S1: Supplementary Statistical Methods** **48**

### 1 Supplementary Figures

#### Figure S1

**Figure S1.** Exercise mode-specific PM_2.5 slopes by region for SBP (Sensitivity C).

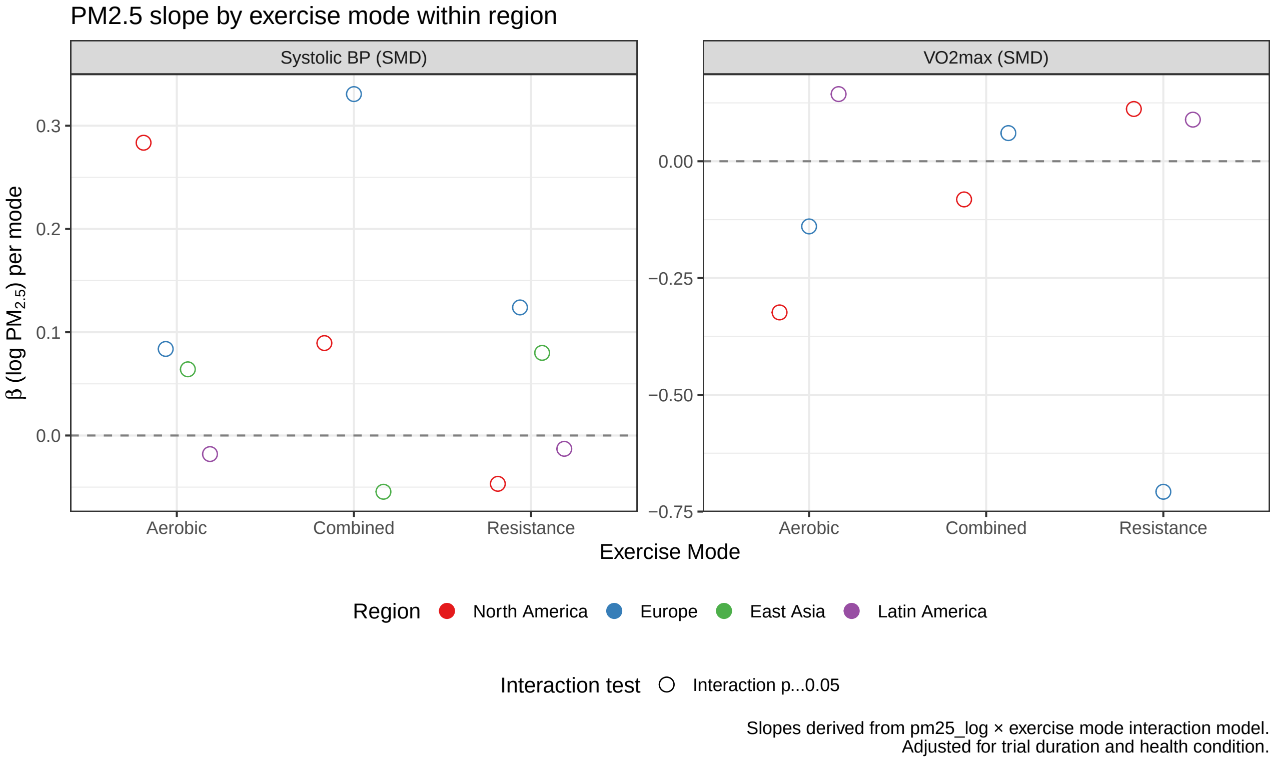

#### Figure S2

**Figure S2.** Standard and contour-enhanced funnel plots for SBP, DBP, VO_2max, and HR. Standard funnel plots (panels 1–4) and contour-enhanced funnel plots (panels 5–8), each ordered SBP, DBP, VO_2max, HR.

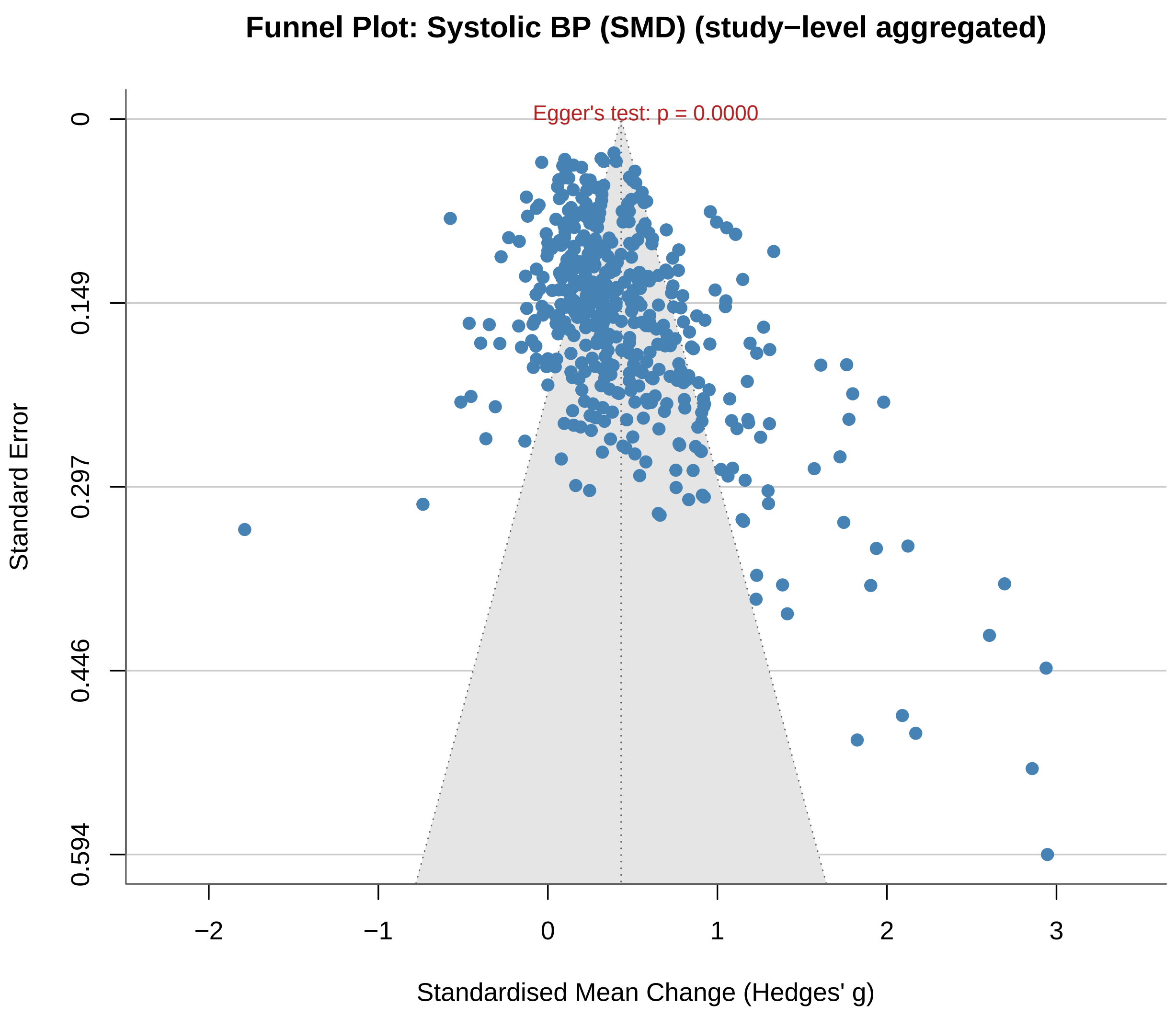

*Figure S2 (continued, panel 2 of 8)*

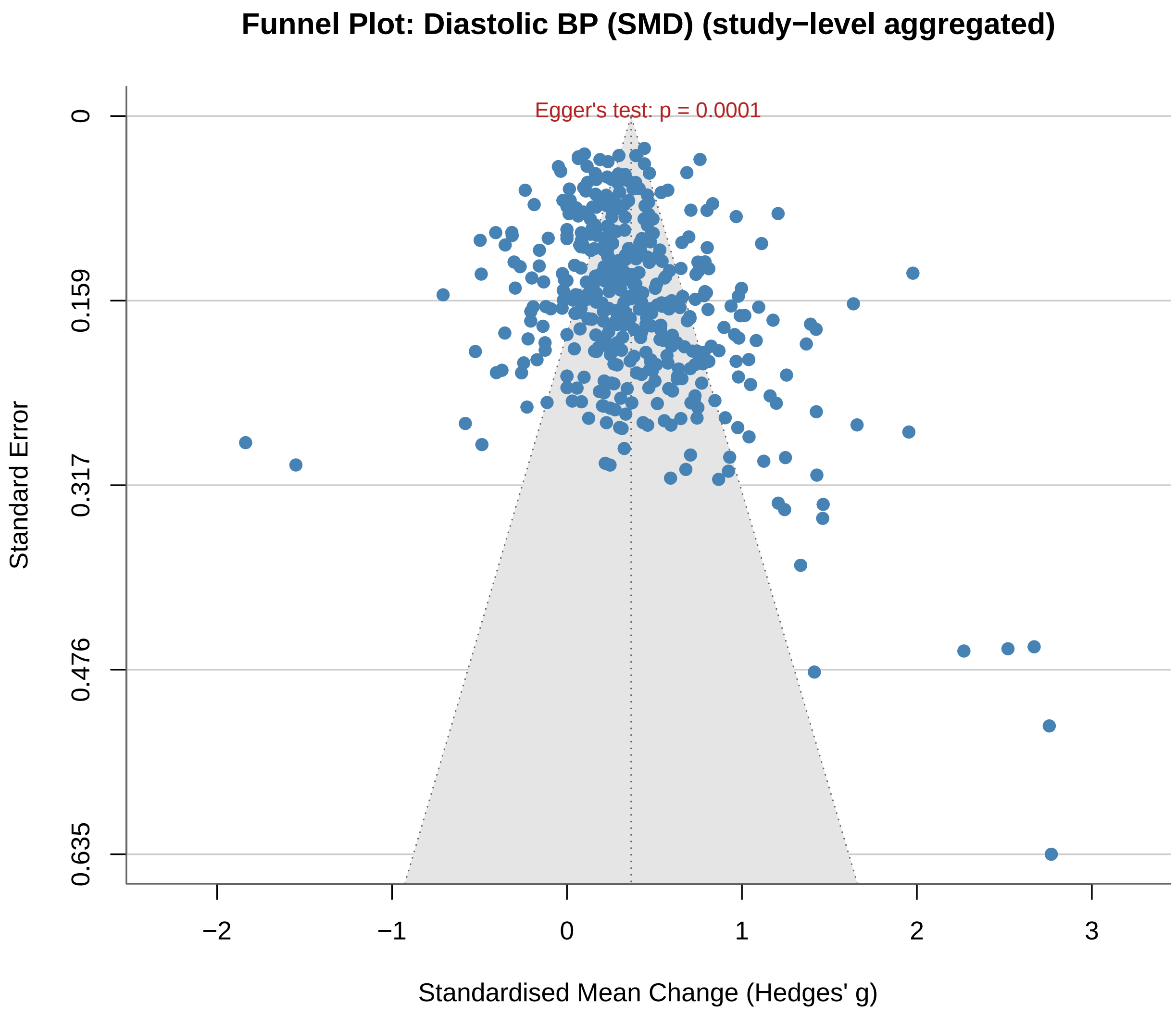

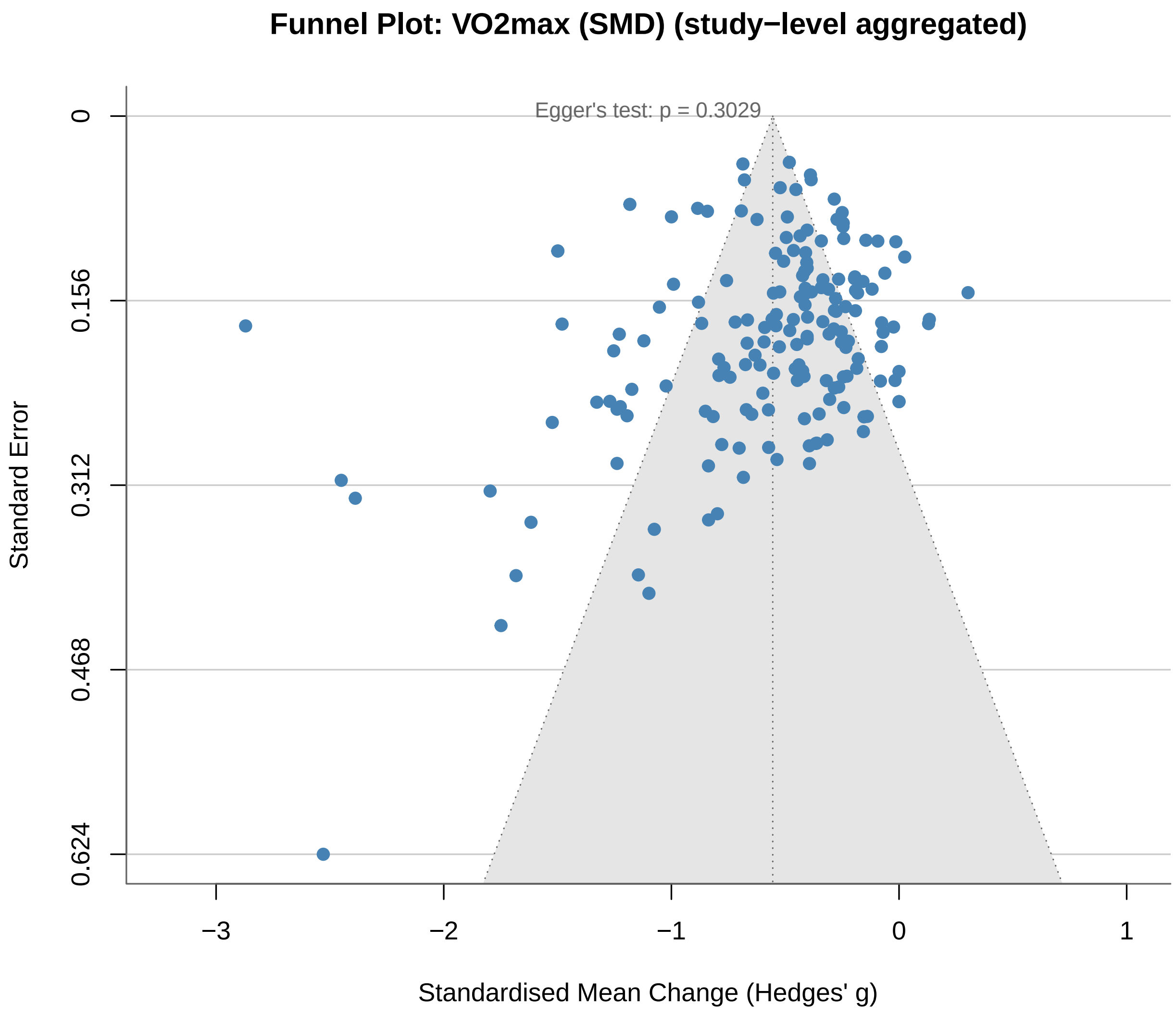
Funnel Plot: VO2max (SMD) (study−level aggregated)
Egger's test: p = 0.3029
Standard Error
0
0.156
0.312
0.468
0.624
−3
−2
−1
0
1
Standardised Mean Change (Hedges' g)

*Figure S2 (continued, panel 4 of 8)*

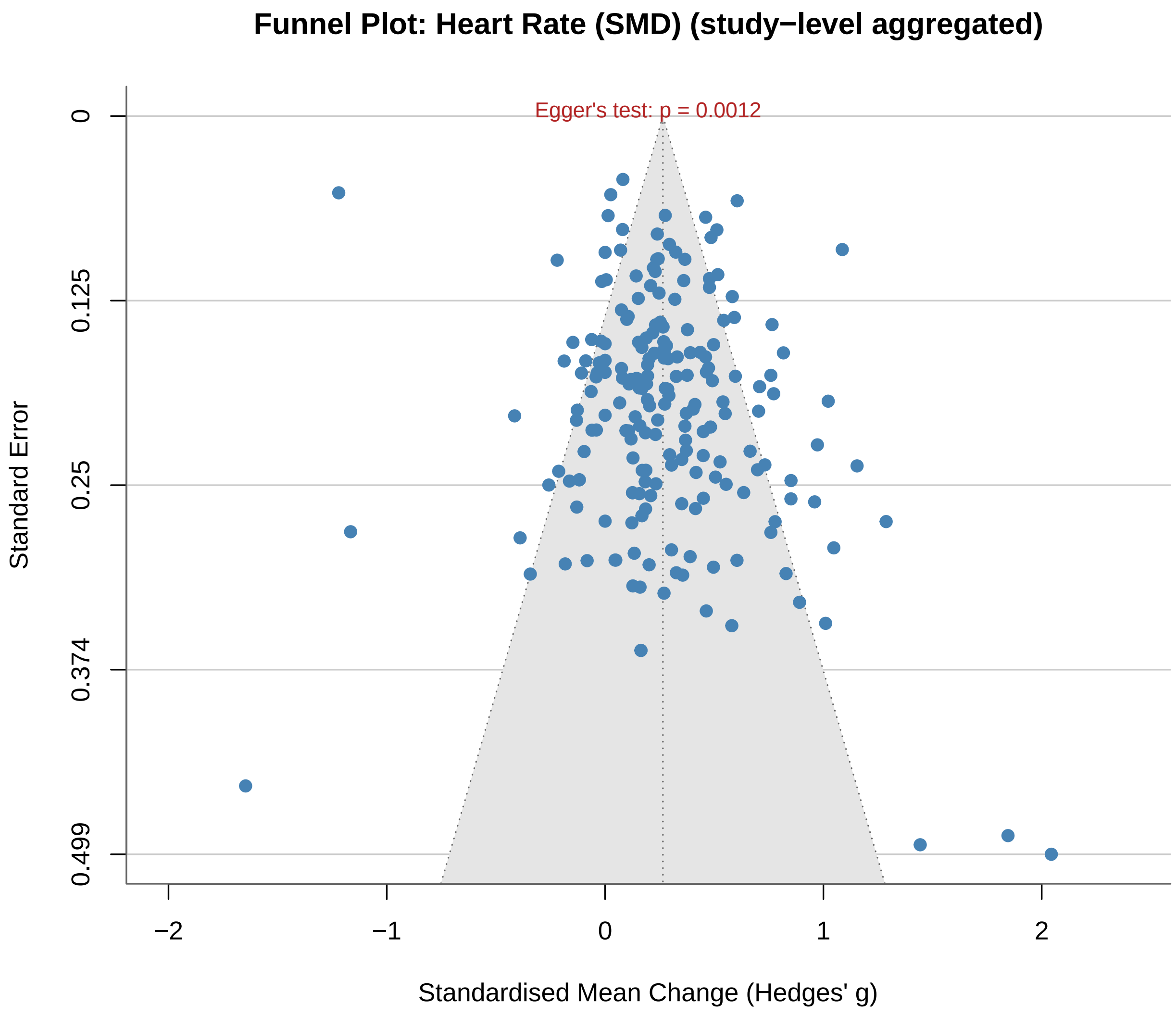

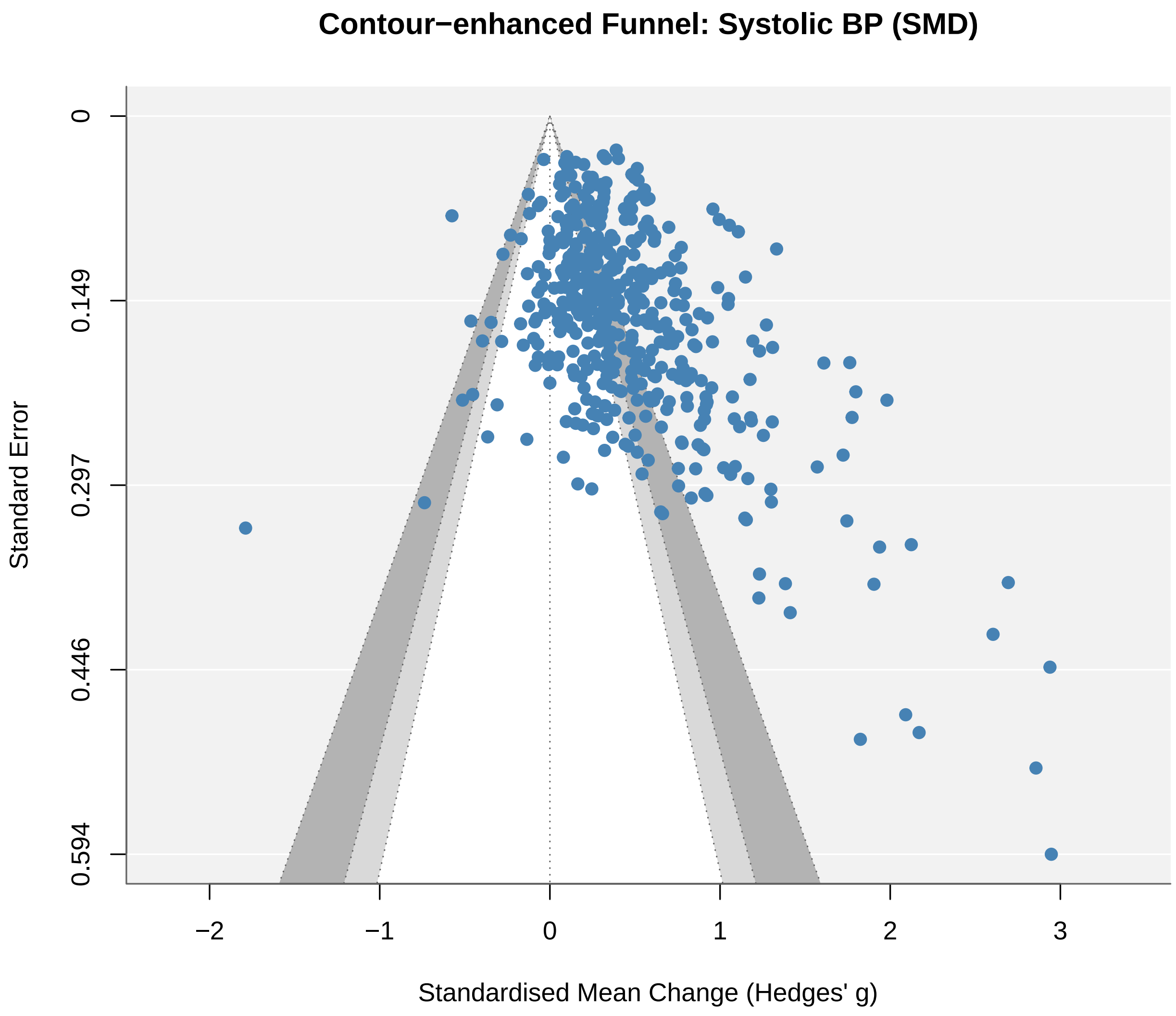
Contour−enhanced Funnel: Systolic BP (SMD)
Standard Error
0
0.149
0.297
0.446
0.594
−2
−1
0
1
2
3
Standardised Mean Change (Hedges' g)

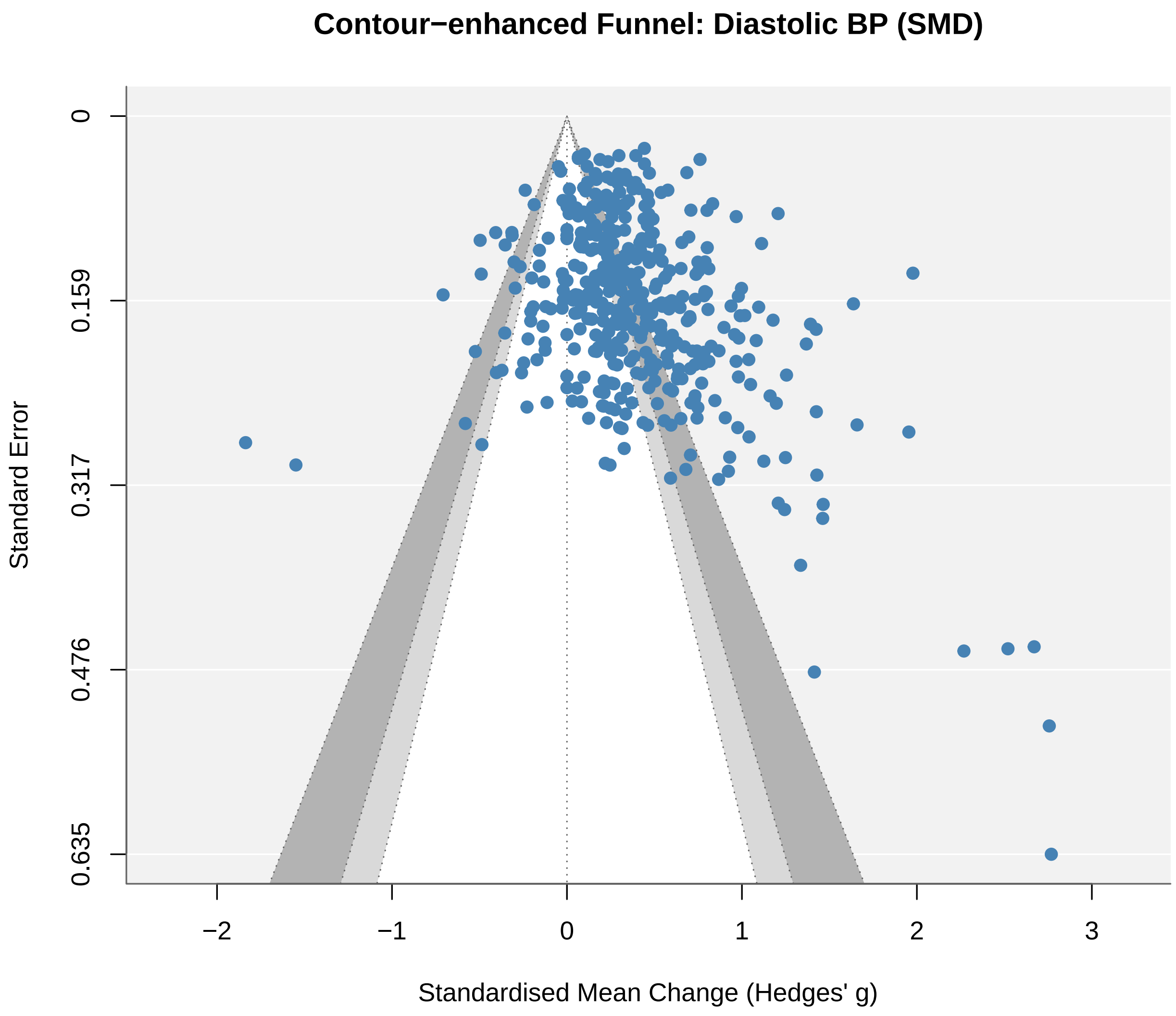
Contour−enhanced Funnel: Diastolic BP (SMD)
Standard Error
0
0.159
0.317
0.476
0.635
−2
−1
0
1
2
3
Standardised Mean Change (Hedges' g)

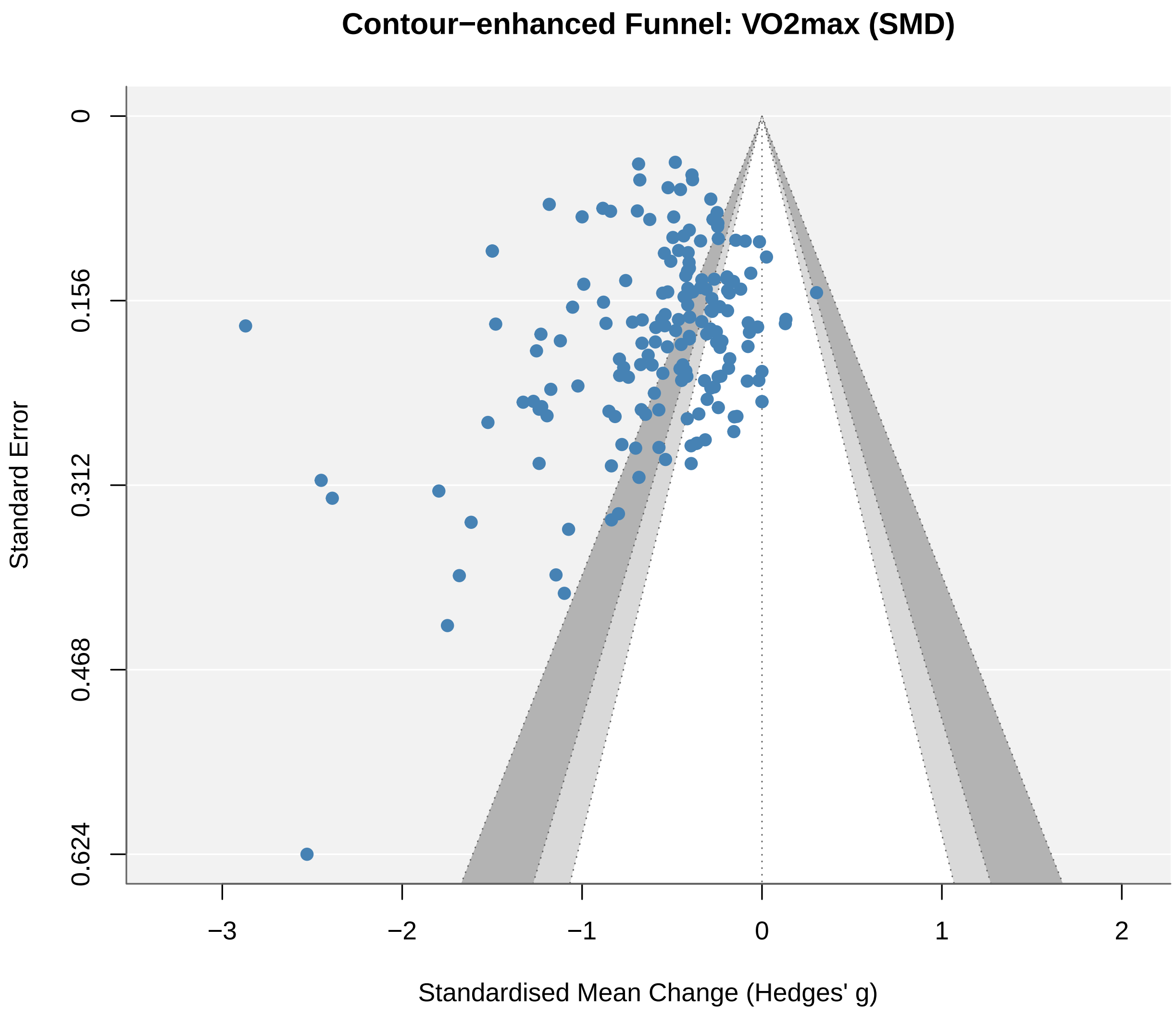
Contour−enhanced Funnel: VO2max (SMD)
Standard Error
0
0.156
0.312
0.468
0.624
−3
−2
−1
0
1
2
Standardised Mean Change (Hedges' g)

*Figure S2 (continued, panel 8 of 8)*

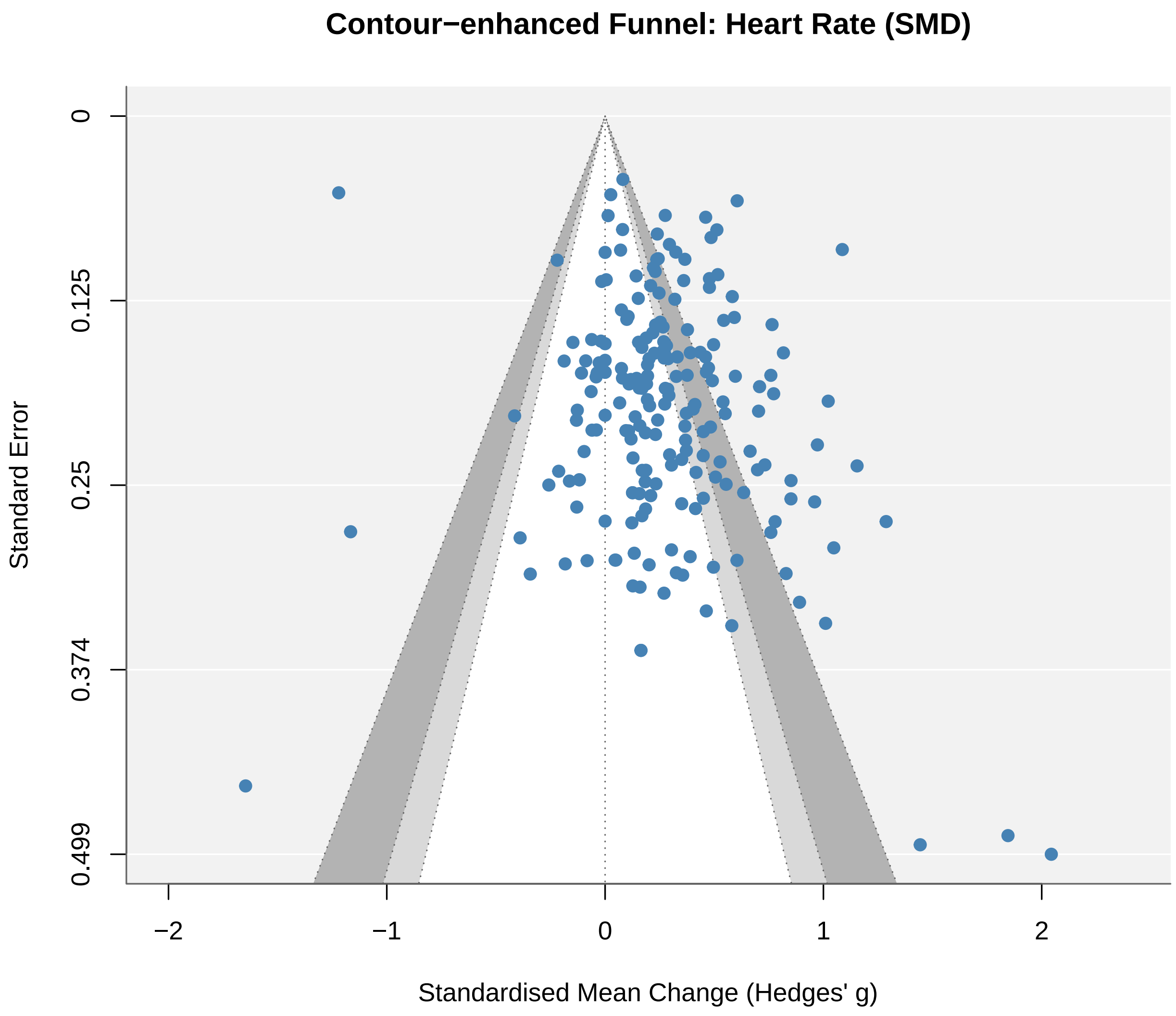

#### Figure S3

**Figure S3.** Bubble plots of PM_2.5 versus effect size for DBP, VO_2max, and HR (one outcome per panel).

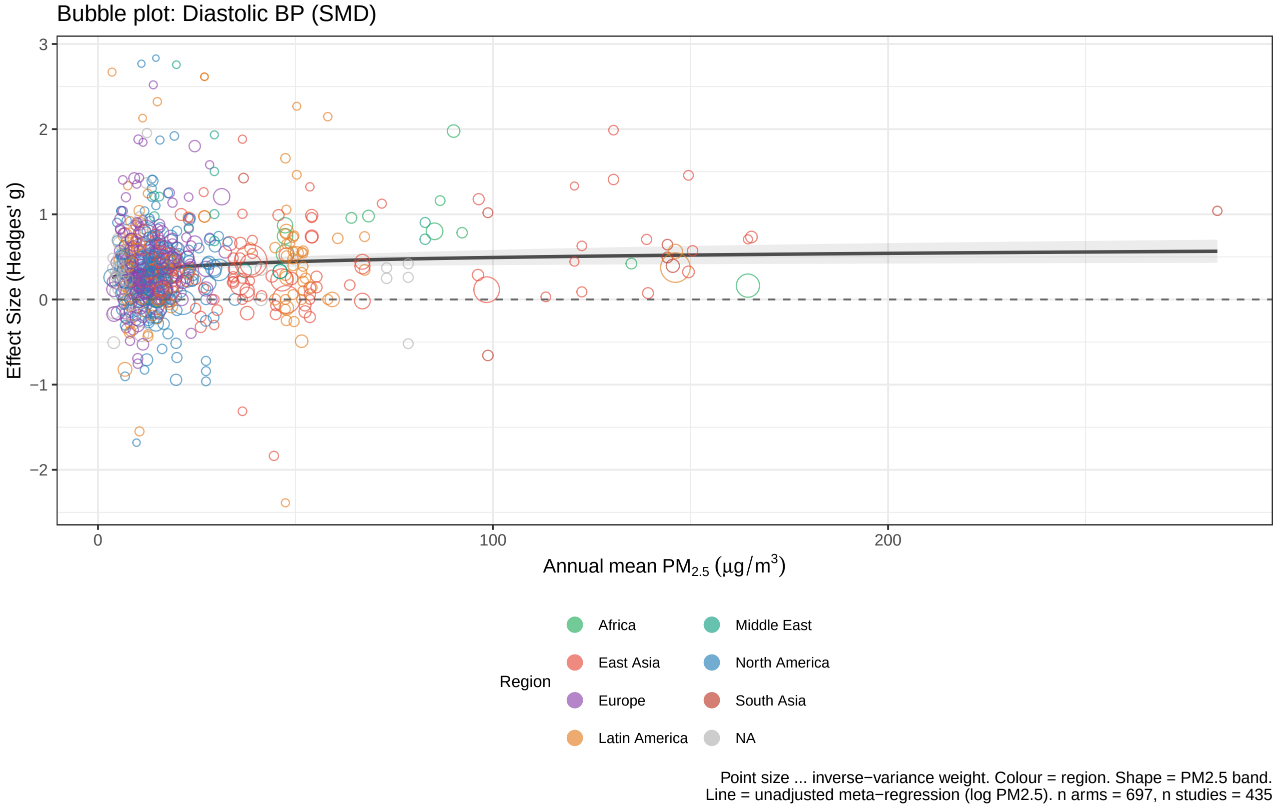

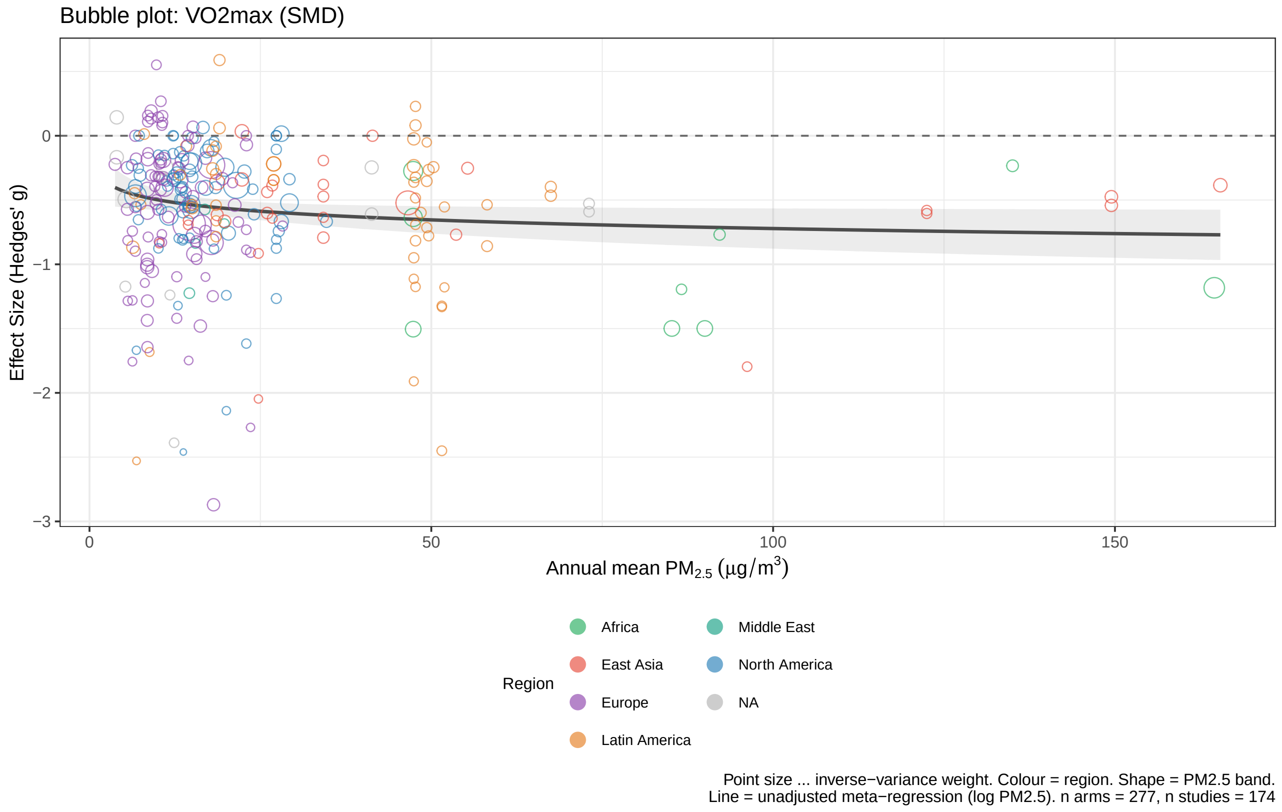
Bubble plot: VO2max (SMD)
Effect Size (Hedges' g)
0
−1
−2
−3
0
50
100
150
Annual mean PM2.5 (µg/m3)
Region
Africa
East Asia
Europe
Latin America
Middle East
North America
NA
Point size ... inverse−variance weight. Colour = region. Shape = PM2.5 band.
Line = unadjusted meta−regression (log PM2.5). n arms = 277, n studies = 174

*Figure S3 (continued, panel 3 of 3)*

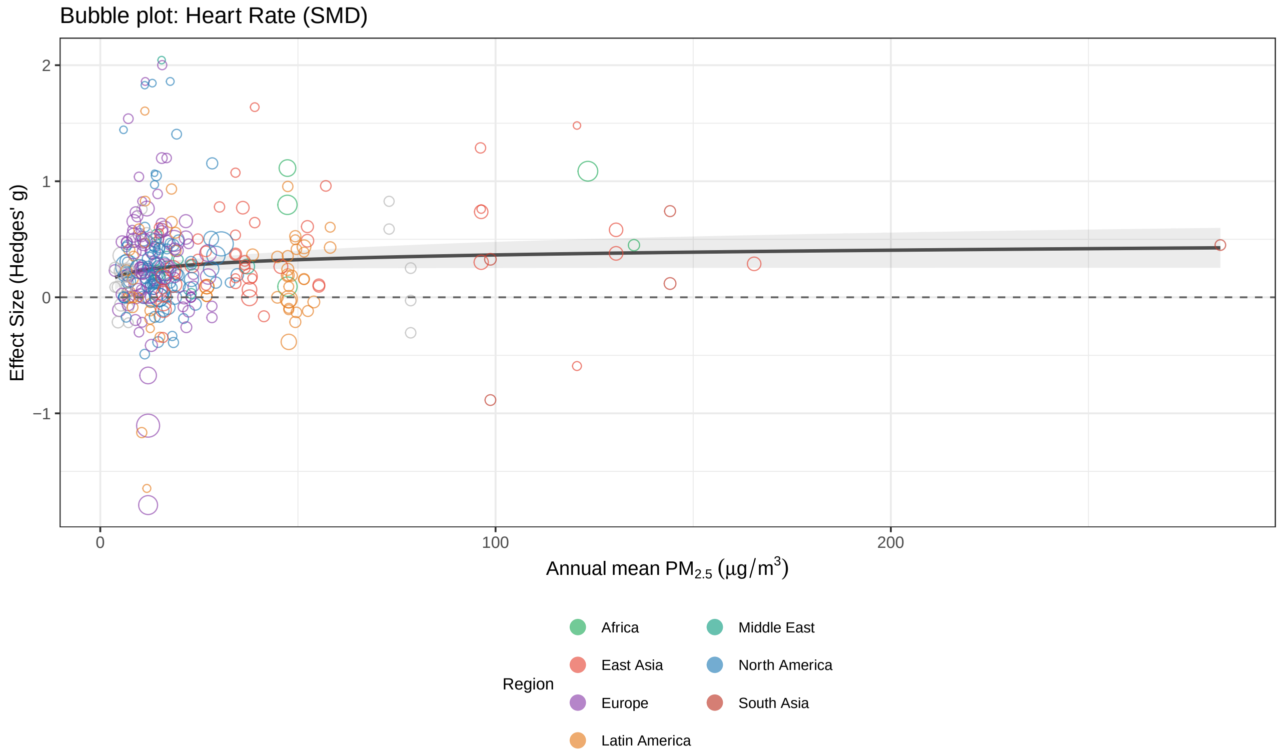

Point size ... inverse−variance weight. Colour = region. Shape = PM2.5 band.
Line = unadjusted meta−regression (log PM2.5). n arms = 327, n studies = 204

### Figure S4

**Figure S4.** Forest plot of exercise training effects on systolic blood pressure.

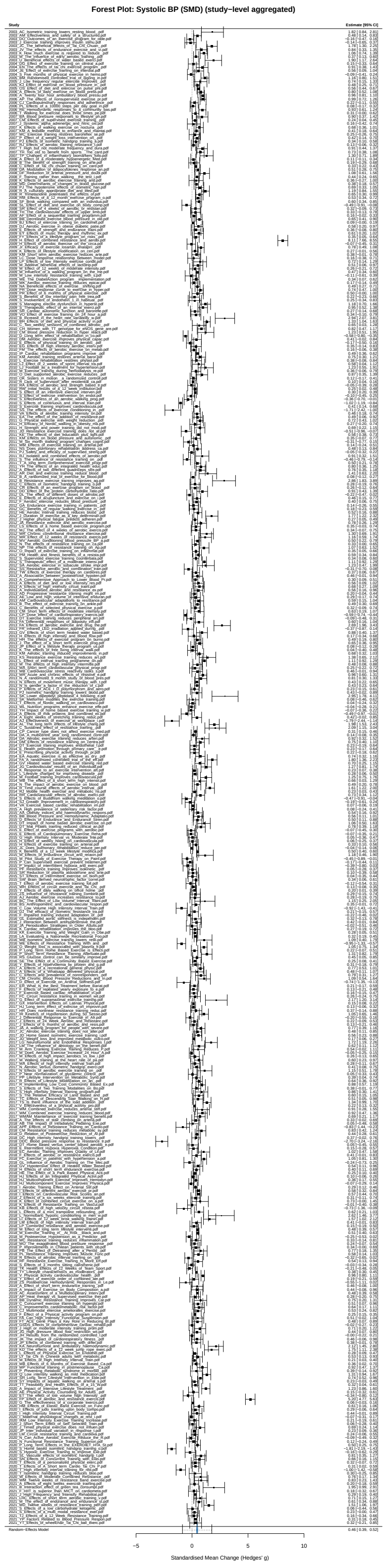

### Figure S5

**Figure S5.** Forest plots for DBP, VO_2max, and HR (one outcome per panel).

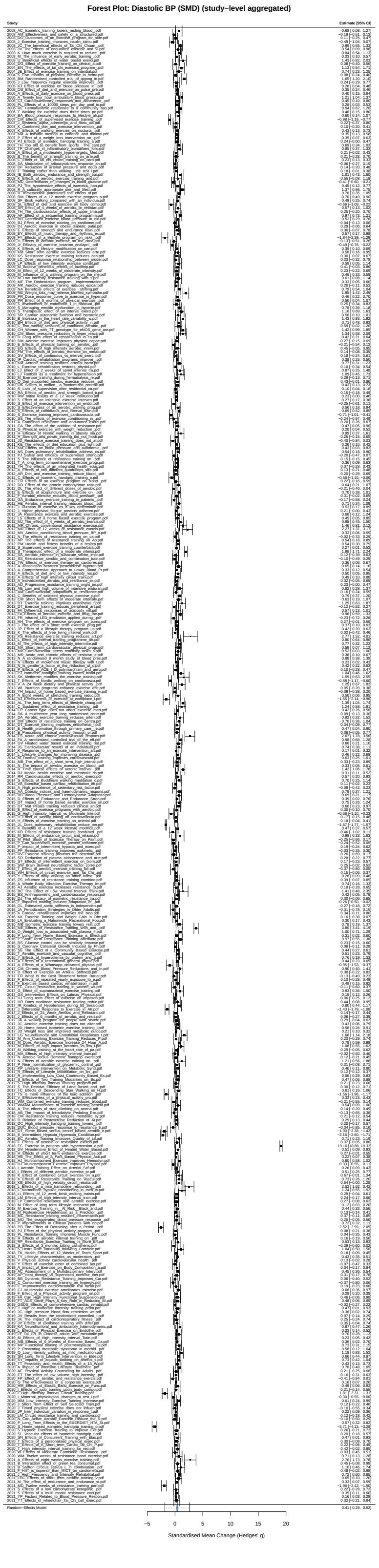

### *Figure S5 (continued, panel 2 of 3)*

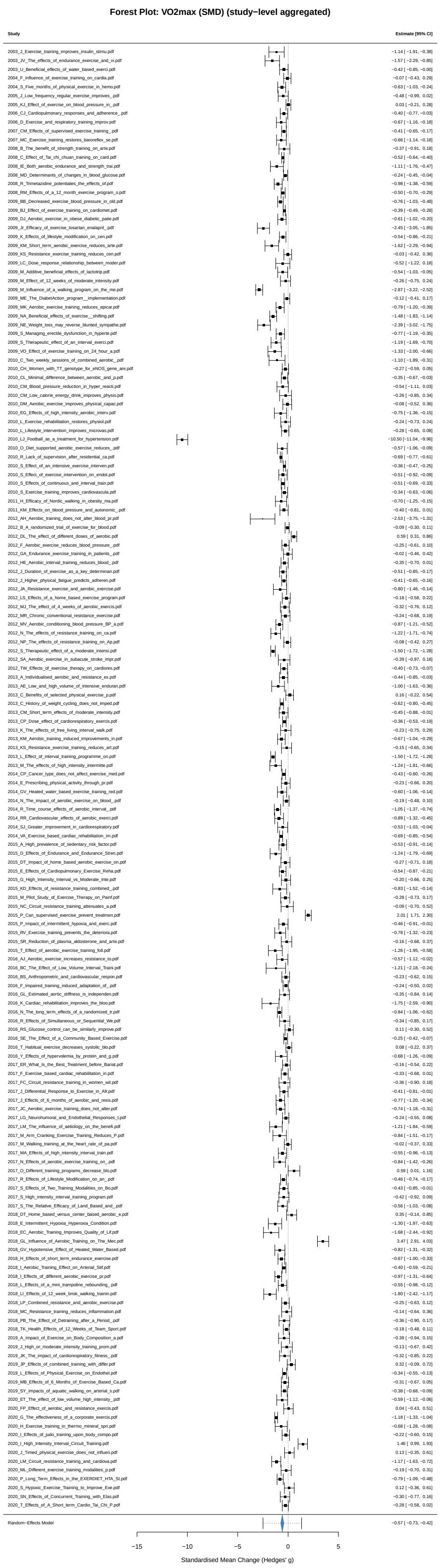

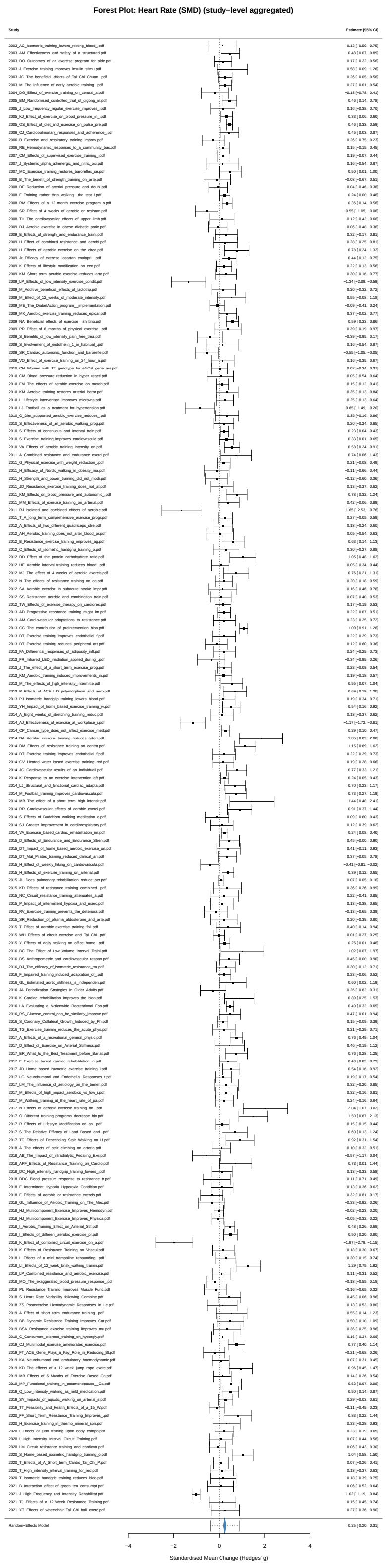
Forest Plot: Heart Rate (SMD) (study−level aggregated)
Study
Estimate [95% CI]
2020_FF_Short_Term_Resistance_Training_Improves_.pdf 0.83 [ 0.22, 1.44]
2020_H_Exercise_training_in_thermo_mineral_spri.pdf 0.33 [−0.28, 0.93]
2020_I_Effects_of_judo_training_upon_body_compo.pdf 0.23 [−0.19, 0.65]
2020_I_High_Intensity_Interval_Circuit_Training.pdf 0.07 [−0.44, 0.58]
2020_LM_Circuit_resistance_training_and_cardiova.pdf −0.06 [−0.43, 0.30]
2020_S_Home_based_isometric_handgrip_training_o.pdf 1.04 [ 0.58, 1.50]
2020_T_Effects_of_A_Short_term_Cardio_Tai_Chi_P.pdf 0.07 [−0.26, 0.41]
2020_T_High_intensity_interval_training_for_red.pdf 0.13 [−0.37, 0.63]
2020_T_Isometric_handgrip_training_reduces_bloo.pdf 0.18 [−0.39, 0.75]
2021_B_Interaction_effect_of_green_tea_consumpt.pdf 0.06 [−0.52, 0.64]
2021_J_High_Frequency_and_Intensity_Rehabilitat.pdf −1.02 [−1.19, −0.84]
2021_TJ_Effects_of_a_12_Week_Resistance_Training.pdf 0.15 [−0.45, 0.74]
2021_YT_Effects_of_wheelchair_Tai_Chi_ball_exerc.pdf 0.27 [−0.36, 0.90]
Random−Effects Model 0.25 [ 0.20, 0.31]
−4
−2
0
2
4
Standardised Mean Change (Hedges' g)

**Figure S6**

**Figure S6.** Dose-response curves for DBP, VO_2max, and HR (one outcome per panel).

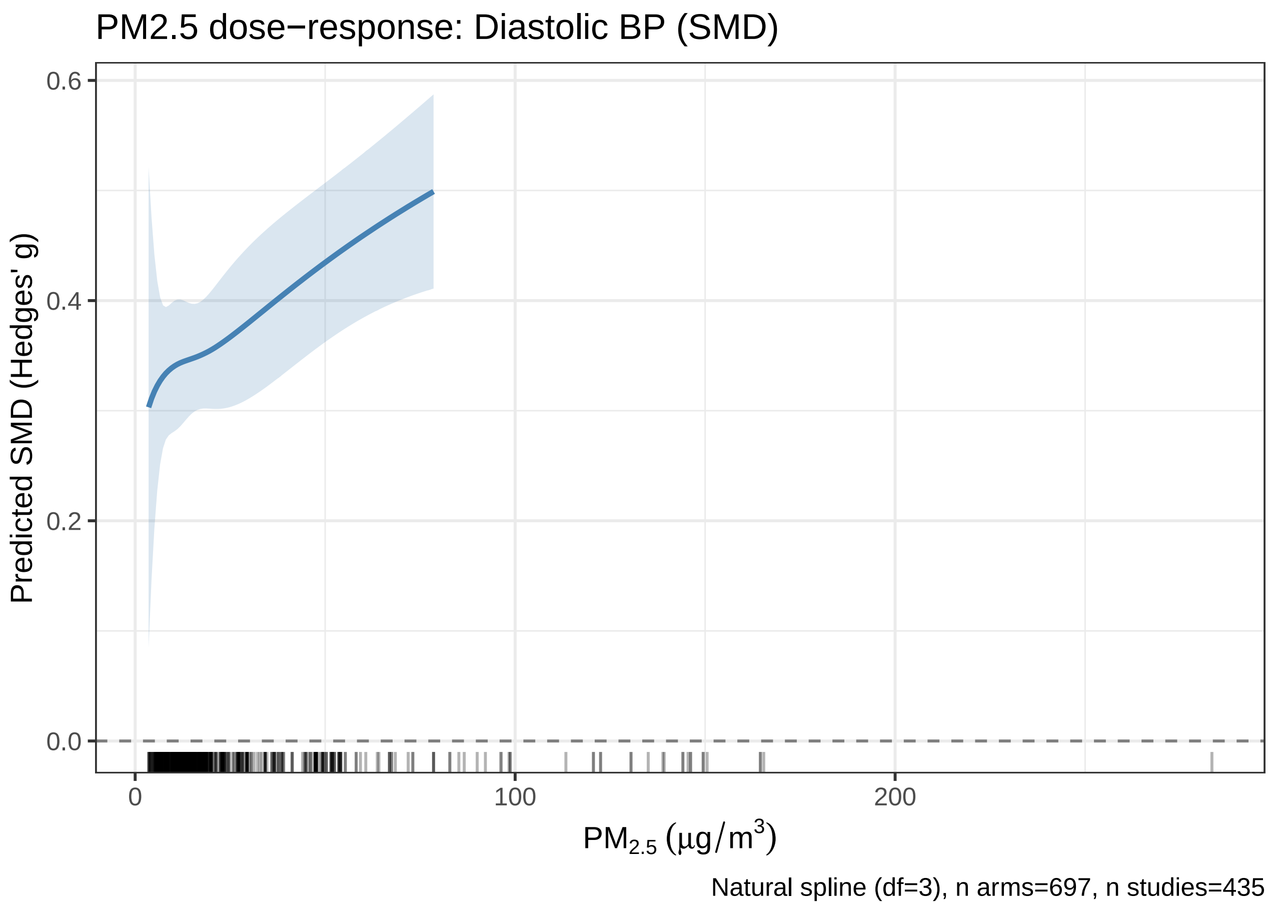

*Figure S6 (continued, panel 2 of 3)*

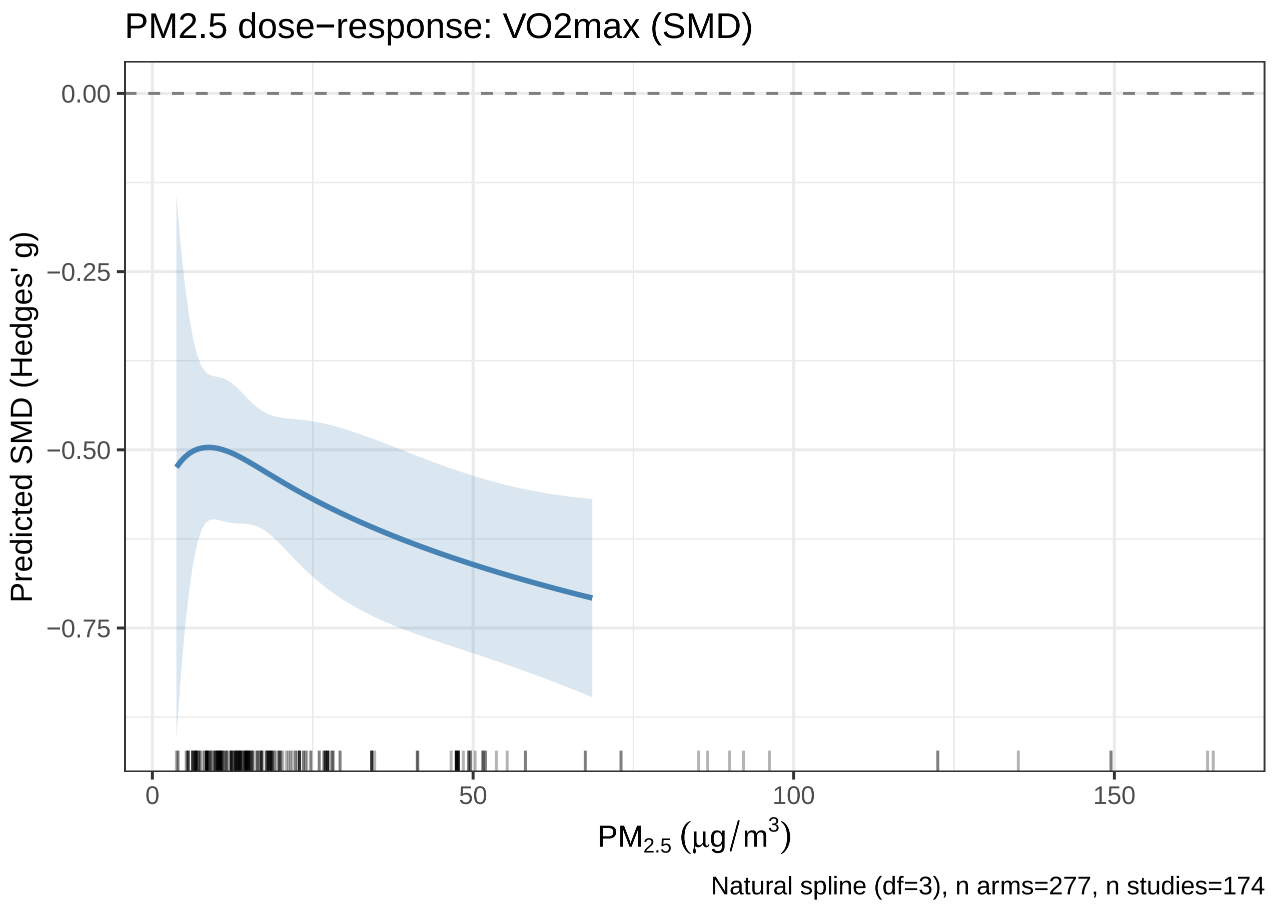

*Figure S6 (continued, panel 3 of 3)*

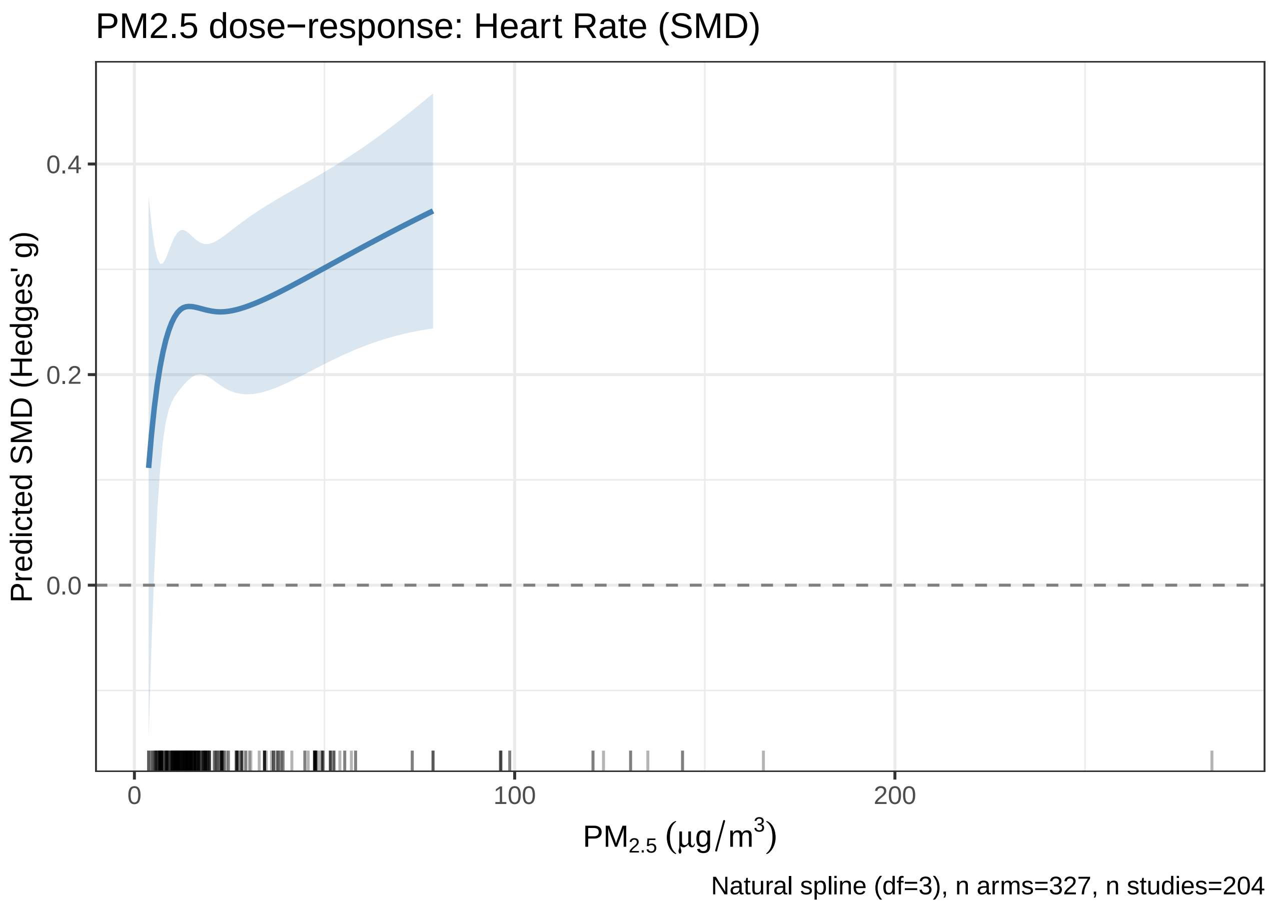

#### Figure S7

**Figure S7.** Risk of bias summary. A) RoB 2 stacked bar chart for randomised trials (n = 316). B) ROBINS-I stacked bar chart for non-randomised intervention studies (n = 149).

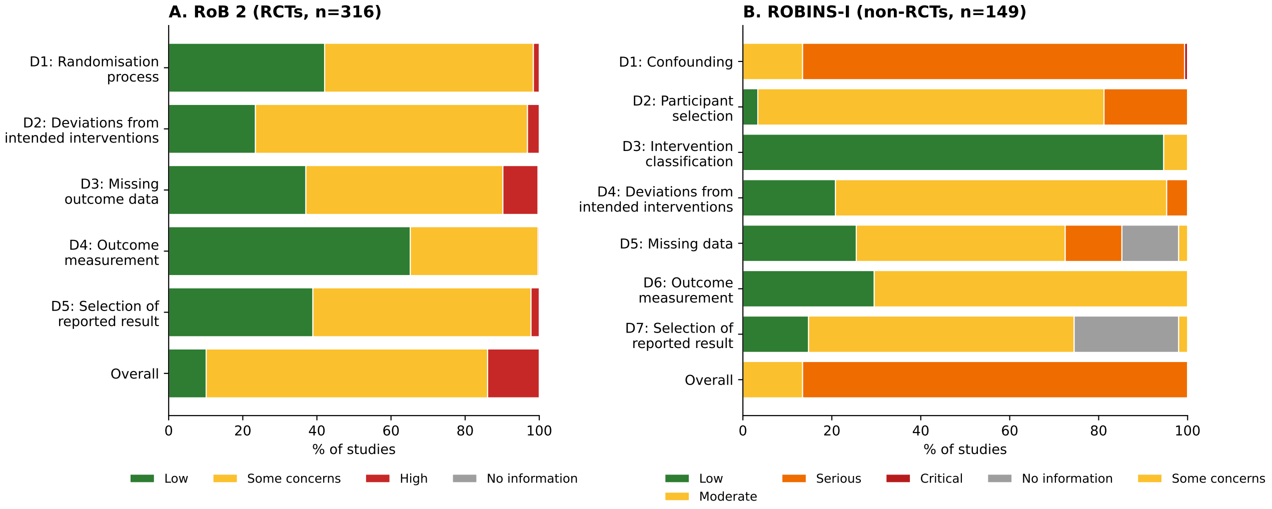

### Figure S8

**Figure S8.** Per-study risk-of-bias traffic-light judgements. A) RoB 2 (n = 316). B) ROBINS-I (n = 149).

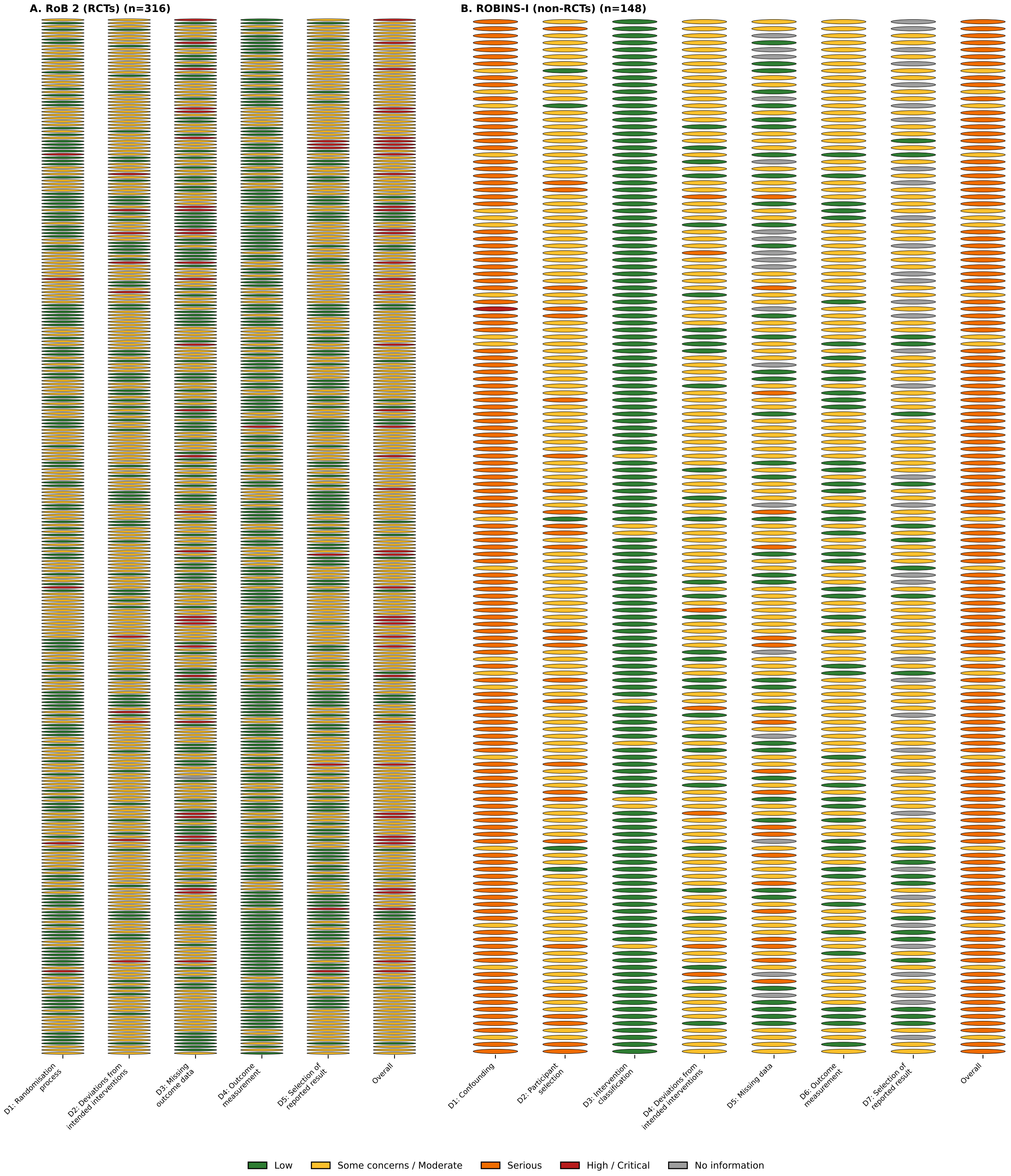

### 2 Supplementary Tables

#### Table S1

**Table S1.** Sensitivity analysis: effect of outlier inclusion on pooled estimates.

| Outcome | Analysis | k | SMD | CI_lower | CI_upper |
|---|---|---|---|---|---|
| SBP | Primary (outliers excluded) | 705 | 0.433 | 0.394 | 0.472 |
| SBP | Sensitivity (all included) | 715 | 0.46 | 0.417 | 0.503 |
| DBP | Primary (outliers excluded) | 697 | 0.372 | 0.334 | 0.41 |
| DBP | Sensitivity (all included) | 703 | 0.386 | 0.346 | 0.426 |
| VO2max | Primary (outliers excluded) | 277 | -0.558 | -0.623 | -0.492 |
| VO2max | Sensitivity (all included) | 280 | -0.569 | -0.636 | -0.503 |
| HR | Primary (outliers excluded) | 327 | 0.267 | 0.218 | 0.315 |
| HR | Sensitivity (all included) | 328 | 0.274 | 0.224 | 0.324 |

#### Table S2

**Table S2.** Spline nonlinearity tests (likelihood ratio tests).

| Outcome | LRT | df | p |
|---|---|---|---|
| SBP | 2.085 | 2 | 0.353 |
| DBP | 1.723 | 2 | 0.423 |
| VO2max | 0.757 | 2 | 0.685 |
| HR | 0.933 | 2 | 0.627 |

#### Table S3

**Table S3.** Regional pooled effects for all four outcomes.

| Outcome | Region | k | N_studies | SMD | CI_lower | CI_upper | abs_change |
|---|---|---|---|---|---|---|---|
| SBP | East Asia | 107 | 69 | 0.473 | 0.367 | 0.578 | 6.6 |
| SBP | Europe | 200 | 125 | 0.383 | 0.313 | 0.454 | 5.4 |
| SBP | Latin America | 136 | 86 | 0.503 | 0.415 | 0.591 | 7.1 |
| SBP | North America | 192 | 117 | 0.353 | 0.288 | 0.418 | 4.9 |
| DBP | East Asia | 103 | 67 | 0.333 | 0.233 | 0.433 | 3 |
| DBP | Europe | 198 | 121 | 0.377 | 0.315 | 0.438 | 3.3 |
| DBP | Latin America | 130 | 83 | 0.387 | 0.286 | 0.488 | 3.4 |
| DBP | North America | 194 | 117 | 0.31 | 0.244 | 0.376 | 2.7 |
| VO2max | East Asia | 29 | 19 | -0.549 | -0.686 | -0.413 | 3.2 |
| VO2max | Europe | 96 | 58 | -0.556 | -0.684 | -0.428 | 3.2 |
| VO2max | Latin America | 52 | 30 | -0.571 | -0.755 | -0.387 | 3.3 |
| VO2max | North America | 75 | 47 | -0.424 | -0.489 | -0.359 | 2.4 |
| HR | East Asia | 50 | 29 | 0.354 | 0.245 | 0.463 | 3.8 |
| HR | Europe | 92 | 54 | 0.276 | 0.168 | 0.384 | 2.9 |
| HR | Latin America | 57 | 37 | 0.156 | 0.036 | 0.276 | 1.7 |
| HR | North America | 86 | 57 | 0.233 | 0.171 | 0.294 | 2.5 |

#### Table S4

**Table S4.** Sensitivity A – common covariate cells: adjusted PM_2.5 coefficients.

| Outcome | Region | k | N_studies | Beta_adj | SE_adj | p_adj |
|---|---|---|---|---|---|---|
| SBP | East Asia | 70 | 52 | 0.0466 | 0.1226 | 0.709 |
| SBP | Europe | 154 | 110 | 0.1338 | 0.0857 | 0.129 |
| SBP | Latin America | 103 | 72 | -0.0155 | 0.0513 | 0.765 |
| SBP | North America | 160 | 106 | 0.095 | 0.07 | 0.184 |

| Outcome | Region | k | N_studies | Beta_adj | SE_adj | p_adj |
|---|---|---|---|---|---|---|
| DBP | East Asia | 68 | 50 | -0.005 | 0.087 | 0.955 |
| DBP | Europe | 153 | 107 | 0.2149 | 0.0927 | 0.027 |
| DBP | Latin America | 97 | 69 | 0.0157 | 0.0706 | 0.825 |
| DBP | North America | 162 | 105 | 0.0457 | 0.0658 | 0.492 |
| HR | Latin America | 45 | 32 | 0.0185 | 0.0487 | 0.711 |
| HR | North America | 57 | 42 | 0.0591 | 0.1393 | 0.677 |

#### Table S5

**Table S5.** Sensitivity B – marginal standardisation: adjusted PM_2.5 coefficients.

| Sensitivity | Outcome | Region | N_Arms | N_Studies | Beta_PM25_log | SE_robust | P_robust | Wt_min | Wt_max | Wt_mean |
|---|---|---|---|---|---|---|---|---|---|---|
| B: Marginal standardisation | Systolic BP (SMD) | North America | 162 | 108 | 0.0888 | 0.0716 | 0.2245 | 0.42 | 2.65 | 1 |
| B: Marginal standardisation | Systolic BP (SMD) | Europe | 157 | 110 | 0.1344 | 0.0884 | 0.1389 | 0.4 | 2.52 | 1 |
| B: Marginal standardisation | Systolic BP (SMD) | East Asia | 93 | 67 | 0.0494 | 0.1022 | 0.6335 | 0.18 | 3.04 | 0.94 |
| B: Marginal standardisation | Systolic BP (SMD) | Latin America | 108 | 76 | -0.0176 | 0.0496 | 0.725 | 0.58 | 2.91 | 0.88 |
| B: Marginal standardisation | Diastolic BP (SMD) | North America | 164 | 107 | 0.0376 | 0.067 | 0.5787 | 0.43 | 2.4 | 1 |
| B: Marginal standardisation | Diastolic BP (SMD) | Europe | 156 | 107 | 0.211 | 0.0915 | 0.0284 | 0.41 | 2.43 | 1 |
| B: Marginal standardisation | Diastolic BP (SMD) | East Asia | 91 | 65 | 0.1169 | 0.0889 | 0.2013 | 0.18 | 3.19 | 0.94 |
| B: Marginal standardisation | Diastolic BP (SMD) | Latin America | 102 | 73 | 0.0174 | 0.0566 | 0.7608 | 0.56 | 2.78 | 0.88 |
| B: Marginal standardisation | VO2max (SMD) | North America | 65 | 43 | -0.0527 | 0.0962 | 0.5954 | 0.31 | 1.88 | 0.95 |
| B: Marginal standardisation | VO2max (SMD) | Europe | 74 | 50 | -0.2124 | 0.2027 | 0.3112 | 0.36 | 3.22 | 0.95 |
| B: Marginal standardisation | VO2max (SMD) | East Asia | 27 | 19 | 0.0273 | 0.1348 | 0.8472 | 0.13 | 1.17 | 0.82 |
| B: Marginal standardisation | VO2max (SMD) | Latin America | 41 | 27 | 0.1135 | 0.1543 | 0.4897 | 0.36 | 1.58 | 0.81 |
| B: Marginal standardisation | Heart Rate (SMD) | North America | 73 | 53 | 0.08 | 0.1101 | 0.4779 | 0.32 | 1.92 | 0.95 |
| B: Marginal standardisation | Heart Rate (SMD) | Europe | 65 | 45 | -0.0609 | 0.1406 | 0.6741 | 0.29 | 2 | 0.93 |
| B: Marginal standardisation | Heart Rate (SMD) | East Asia | 42 | 28 | 0.2365 | 0.1224 | 0.0902 | 0.23 | 2.58 | 0.87 |

#### Table S6

**Table S6.** Sensitivity C – Wald tests (a) and mode-specific slopes (b).
*(a)*

| Outcome | Region | N_Arms | N_Studies | Interaction_p | Significant |
|---|---|---|---|---|---|
| Systolic BP (SMD) | North America | 160 | 106 | 0.1883 | FALSE |
| Systolic BP (SMD) | Europe | 154 | 110 | 0.6456 | FALSE |
| Systolic BP (SMD) | East Asia | 86 | 62 | 0.8843 | FALSE |
| Systolic BP (SMD) | Latin America | 107 | 72 | 0.9652 | FALSE |
| VO2max (SMD) | North America | 64 | 42 | 0.2933 | FALSE |
| VO2max (SMD) | Europe | 74 | 50 | 0.2652 | FALSE |
| VO2max (SMD) | Latin America | 41 | 26 | 0.8065 | FALSE |

*(b)*

| Outcome | Region | Exercise_Mode | N_Arms | Beta_PM25_log | Interaction_p |
|---|---|---|---|---|---|
| Systolic BP (SMD) | North America | Aerobic | 71 | 0.2835 | 0.1883 |
| Systolic BP (SMD) | North America | Resistance | 61 | -0.0468 | 0.1883 |
| Systolic BP (SMD) | North America | Combined | 28 | 0.0895 | 0.1883 |
| Systolic BP (SMD) | Europe | Aerobic | 86 | 0.0838 | 0.6456 |
| Systolic BP (SMD) | Europe | Resistance | 46 | 0.124 | 0.6456 |
| Systolic BP (SMD) | Europe | Combined | 22 | 0.3306 | 0.6456 |
| Systolic BP (SMD) | East Asia | Aerobic | 36 | 0.0641 | 0.8843 |
| Systolic BP (SMD) | East Asia | Resistance | 34 | 0.08 | 0.8843 |
| Systolic BP (SMD) | East Asia | Combined | 16 | -0.0545 | 0.8843 |
| Systolic BP (SMD) | Latin America | Aerobic | 51 | -0.018 | 0.9652 |
| Systolic BP (SMD) | Latin America | Resistance | 52 | -0.0129 | 0.9652 |
| VO2max (SMD) | North America | Aerobic | 34 | -0.3236 | 0.2933 |
| VO2max (SMD) | North America | Resistance | 17 | 0.1119 | 0.2933 |
| VO2max (SMD) | North America | Combined | 13 | -0.0817 | 0.2933 |
| VO2max (SMD) | Europe | Aerobic | 44 | -0.1395 | 0.2652 |
| VO2max (SMD) | Europe | Resistance | 19 | -0.7076 | 0.2652 |
| VO2max (SMD) | Europe | Combined | 11 | 0.0603 | 0.2652 |
| VO2max (SMD) | Latin America | Aerobic | 31 | 0.1438 | 0.8065 |
| VO2max (SMD) | Latin America | Resistance | 8 | 0.0891 | 0.8065 |

#### Table S7

**Table S7.** Sensitivity D – duration-stratified PM_2.5 coefficients (a) and duration × PM_2.5 interaction tests (b).
*(a)*

| Outcome | Duration | k | Beta_adj | SE | p |
|---|---|---|---|---|---|
| SBP | Short (¡12 weeks) | 198 | 0.0721 | 0.0655 | 0.277 |
| SBP | Medium (12-24 weeks) | 270 | 0.0255 | 0.0481 | 0.597 |
| SBP | Long (¿24 weeks) | 51 | 0.2675 | 0.1485 | 0.112 |
| DBP | Short (¡12 weeks) | 195 | 0.0982 | 0.0561 | 0.088 |
| DBP | Medium (12-24 weeks) | 265 | 0.0637 | 0.0564 | 0.262 |
| DBP | Long (¿24 weeks) | 52 | 0.1101 | 0.1405 | 0.457 |

*(b)*

| Outcome | LRT | df | p |
|---|---|---|---|
| SBP | 0.091 | 2 | 0.955 |
| DBP | 2.273 | 2 | 0.321 |
| VO2max | 3.513 | 2 | 0.173 |
| HR | 1.829 | 2 | 0.401 |

#### Table S8

**Table S8.** Sequential covariate adjustment for the T3 vs T1 SBP effect size contrast.

| Model | T3_vs_T1_beta | T3_vs_T1_p | Attenuation_pct |
|---|---|---|---|
| Unadjusted (tertile only) | 0.1319 | 0.0063 | 0 |
| + Health condition | 0.1311 | 0.0058 | 0.6 |
| + Region | 0.0845 | 0.1402 | 35.9 |
| + Exercise mode | 0.0482 | 0.437 | 63.4 |
| + Trial duration | 0.0328 | 0.5988 | 75.1 |
| + Baseline SBP | 0.0268 | 0.6468 | 79.7 |

#### Table S9

**Table S9.** Four-strata pooled effects by PM_2.5 level.

| **Outcome** | **Stratum** | **N_Arms** | **N_Studies** | **ES** | **SE_robust** | **CI_Lower** | **CI_Upper** | **P_robust** | **I2_L3** | **I2_L2** |
|---|---|---|---|---|---|---|---|---|---|---|
| Systolic BP (SMD) | Low (¡22) | 485 | 300 | 0.375 | 0.022 | 0.333 | 0.418 | 0 | 66.2 | 33.8 |
| Systolic BP (SMD) | Moderate (22-60) | 170 | 108 | 0.51 | 0.043 | 0.426 | 0.594 | 0 | 43.1 | 56.9 |
| Systolic BP (SMD) | High (¿60) | 50 | 34 | 0.692 | 0.098 | 0.499 | 0.885 | 0 | 85 | 15 |
| Diastolic BP (SMD) | Low (¡22) | 483 | 296 | 0.335 | 0.021 | 0.293 | 0.377 | 0 | 56.2 | 43.8 |
| Diastolic BP (SMD) | Moderate (22-60) | 166 | 105 | 0.392 | 0.046 | 0.302 | 0.482 | 0 | 43.6 | 56.4 |
| Diastolic BP (SMD) | High (¿60) | 48 | 34 | 0.616 | 0.08 | 0.46 | 0.773 | 0 | 28.5 | 71.5 |
| VO2max (SMD) | Low (¡22) | 185 | 119 | -0.519 | 0.04 | -0.598 | -0.44 | 0 | 63.2 | 36.8 |
| VO2max (SMD) | Moderate (22-60) | 76 | 43 | -0.58 | 0.059 | -0.696 | -0.463 | 0 | 23.8 | 76.2 |
| VO2max (SMD) | High (¿60) | 16 | 12 | -0.876 | 0.146 | -1.163 | -0.589 | 1e-04 | 100 | 0 |
| Heart Rate (SMD) | Low (¡22) | 216 | 134 | 0.25 | 0.032 | 0.188 | 0.313 | 0 | 64.1 | 35.9 |
| Heart Rate (SMD) | Moderate (22-60) | 90 | 58 | 0.266 | 0.038 | 0.192 | 0.34 | 0 | 43.4 | 56.6 |
| Heart Rate (SMD) | High (¿60) | 21 | 12 | 0.42 | 0.118 | 0.189 | 0.651 | 0.0054 | 9.6 | 90.4 |

#### Table S10

**Table S10.** Moderator-stratified analysis: SBP and $VO_2max$ by $PM_{2.5}$ tertile.

| **Outcome** | **PM25_Stratum** | **Moderator** | **Level** | **N_Arms** | **N_Studies** | **ES** | **CI_Lower** | **CI_Upper** | **P** |
|---|---|---|---|---|---|---|---|---|---|
| SBP | Low (¡22) | Exercise_Type | Aerobic | 180 | 131 | 0.396 | 0.323 | 0.469 | 0 |
| SBP | Low (¡22) | Exercise_Type | Resistance | 157 | 119 | 0.426 | 0.36 | 0.492 | 0 |
| SBP | Low (¡22) | Exercise_Type | Combined | 49 | 36 | 0.345 | 0.242 | 0.449 | 0 |
| SBP | Low (¡22) | Exercise_Type | Mind-Body | 4 | 3 | 0.058 | -0.558 | 0.673 | 0.8546 |
| SBP | Low (¡22) | Exercise_Location | Indoor | 347 | 233 | 0.401 | 0.351 | 0.451 | 0 |
| SBP | Low (¡22) | Exercise_Location | Outdoor | 26 | 22 | 0.413 | 0.255 | 0.571 | 0 |
| SBP | Low (¡22) | Intensity | Vigorous | 132 | 104 | 0.391 | 0.317 | 0.465 | 0 |
| SBP | Low (¡22) | Intensity | Moderate | 149 | 111 | 0.353 | 0.278 | 0.427 | 0 |
| SBP | Low (¡22) | Intensity | Light | 117 | 88 | 0.429 | 0.34 | 0.518 | 0 |
| SBP | Low (¡22) | Health_Condition | CVD | 19 | 13 | 0.249 | 0.025 | 0.474 | 0.0295 |
| SBP | Low (¡22) | Health_Condition | Metabolic | 77 | 47 | 0.461 | 0.329 | 0.593 | 0 |
| SBP | Low (¡22) | Health_Condition | Healthy | 68 | 41 | 0.319 | 0.213 | 0.424 | 0 |
| SBP | Low (¡22) | Health_Condition | Diabetes | 38 | 27 | 0.331 | 0.19 | 0.471 | 0 |
| SBP | Low (¡22) | Health_Condition | Other | 47 | 36 | 0.221 | 0.083 | 0.359 | 0.0016 |
| SBP | Low (¡22) | Health_Condition | Hypertensive | 236 | 144 | 0.412 | 0.356 | 0.468 | 0 |
| SBP | Moderate (22-60) | Exercise_Type | Aerobic | 70 | 49 | 0.552 | 0.415 | 0.688 | 0 |
| SBP | Moderate (22-60) | Exercise_Type | Resistance | 52 | 42 | 0.495 | 0.362 | 0.628 | 0 |
| SBP | Moderate (22-60) | Exercise_Type | Combined | 20 | 13 | 0.453 | 0.266 | 0.64 | 0 |
| SBP | Moderate (22-60) | Exercise_Type | Mind-Body | 6 | 6 | 0.627 | 0.178 | 1.077 | 0.0062 |
| SBP | Moderate (22-60) | Exercise_Location | Indoor | 122 | 84 | 0.56 | 0.459 | 0.66 | 0 |
| SBP | Moderate (22-60) | Exercise_Location | Outdoor | 8 | 6 | 0.469 | 0.235 | 0.703 | 1e-04 |
| SBP | Moderate (22-60) | Intensity | Moderate | 71 | 52 | 0.547 | 0.428 | 0.667 | 0 |
| SBP | Moderate (22-60) | Intensity | Light | 37 | 29 | 0.468 | 0.305 | 0.631 | 0 |
| SBP | Moderate (22-60) | Intensity | Vigorous | 34 | 29 | 0.44 | 0.275 | 0.605 | 0 |
| SBP | Moderate (22-60) | Health_Condition | Hypertensive | 81 | 58 | 0.691 | 0.552 | 0.831 | 0 |
| SBP | Moderate (22-60) | Health_Condition | Other | 22 | 15 | 0.483 | 0.364 | 0.603 | 0 |
| SBP | Moderate (22-60) | Health_Condition | Diabetes | 17 | 8 | 0.251 | 0.007 | 0.495 | 0.044 |
| SBP | Moderate (22-60) | Health_Condition | Healthy | 22 | 16 | 0.231 | 0.058 | 0.403 | 0.0088 |
| SBP | Moderate (22-60) | Health_Condition | Metabolic | 20 | 14 | 0.442 | 0.304 | 0.58 | 0 |
| SBP | Moderate (22-60) | Health_Condition | CVD | 8 | 4 | 0.029 | -0.619 | 0.677 | 0.9297 |
| SBP | High (¿60) | Exercise_Type | Aerobic | 23 | 18 | 0.828 | 0.487 | 1.168 | 0 |
| SBP | High (¿60) | Exercise_Type | Resistance | 10 | 10 | 0.489 | 0.267 | 0.711 | 0 |
| SBP | High (¿60) | Exercise_Type | Mind-Body | 3 | 3 | 0.932 | 0.44 | 1.424 | 2e-04 |
| SBP | High (¿60) | Exercise_Type | Combined | 3 | 3 | 0.676 | 0.353 | 0.998 | 0 |
| SBP | High (¿60) | Exercise_Location | Outdoor | 4 | 4 | 0.877 | 0.157 | 1.596 | 0.0169 |
| SBP | High (¿60) | Exercise_Location | Indoor | 36 | 26 | 0.778 | 0.547 | 1.01 | 0 |
| SBP | High (¿60) | Intensity | Light | 13 | 10 | 0.613 | 0.353 | 0.872 | 0 |
| SBP | High (¿60) | Intensity | Moderate | 24 | 21 | 0.799 | 0.51 | 1.089 | 0 |
| SBP | High (¿60) | Intensity | Vigorous | 3 | 3 | 0.82 | -0.511 | 2.151 | 0.227 |
| SBP | High (¿60) | Health_Condition | Diabetes | 8 | 4 | 0.722 | 0.026 | 1.419 | 0.0421 |
| SBP | High (¿60) | Health_Condition | Hypertensive | 28 | 22 | 0.821 | 0.577 | 1.065 | 0 |
| SBP | High (¿60) | Health_Condition | Healthy | 3 | 1 | 0.123 | 0.03 | 0.216 | 0.0099 |
| SBP | High (¿60) | Health_Condition | Metabolic | 10 | 6 | 0.348 | 0.029 | 0.668 | 0.0327 |
| VO2max | Low (¡22) | Exercise_Type | Aerobic | 84 | 59 | -0.668 | -0.807 | -0.528 | 0 |
| VO2max | Low (¡22) | Exercise_Type | Resistance | 44 | 34 | -0.421 | -0.518 | -0.325 | 0 |
| VO2max | Low (¡22) | Exercise_Type | Combined | 25 | 19 | -0.501 | -0.654 | -0.349 | 0 |
| VO2max | Low (¡22) | Exercise_Location | Indoor | 148 | 98 | -0.557 | -0.649 | -0.465 | 0 |
| VO2max | Low (¡22) | Exercise_Location | Outdoor | 6 | 6 | -0.554 | -0.917 | -0.191 | 0.0028 |
| VO2max | Low (¡22) | Intensity | Light | 31 | 22 | -0.404 | -0.561 | -0.247 | 0 |
| VO2max | Low (¡22) | Intensity | Moderate | 66 | 51 | -0.573 | -0.682 | -0.463 | 0 |

| Outcome | PM25_Stratum | Moderator | Level | N_Arms | N_Studies | ES | CI_Lower | CI_Upper | P |
|---|---|---|---|---|---|---|---|---|---|
| VO2max | Low (¡22) | Intensity | Vigorous | 65 | 48 | -0.624 | -0.783 | -0.464 | 0 |
| VO2max | Low (¡22) | Health_Condition | Healthy | 16 | 12 | -0.383 | -0.565 | -0.202 | 0 |
| VO2max | Low (¡22) | Health_Condition | Diabetes | 18 | 14 | -0.327 | -0.494 | -0.161 | 1e-04 |
| VO2max | Low (¡22) | Health_Condition | Other | 27 | 21 | -0.669 | -0.941 | -0.397 | 0 |
| VO2max | Low (¡22) | Health_Condition | Hypertensive | 83 | 51 | -0.469 | -0.569 | -0.369 | 0 |
| VO2max | Low (¡22) | Health_Condition | Metabolic | 30 | 19 | -0.675 | -0.931 | -0.418 | 0 |
| VO2max | Low (¡22) | Health_Condition | CVD | 11 | 7 | -0.663 | -0.87 | -0.455 | 0 |
| VO2max | Moderate (22-60) | Exercise_Type | Resistance | 8 | 8 | -0.331 | -0.497 | -0.164 | 1e-04 |
| VO2max | Moderate (22-60) | Exercise_Type | Aerobic | 43 | 28 | -0.748 | -0.895 | -0.601 | 0 |
| VO2max | Moderate (22-60) | Exercise_Type | Combined | 7 | 4 | -0.545 | -0.86 | -0.231 | 7e-04 |
| VO2max | Moderate (22-60) | Exercise_Location | Indoor | 58 | 37 | -0.62 | -0.769 | -0.47 | 0 |
| VO2max | Moderate (22-60) | Exercise_Location | Outdoor | 4 | 4 | -0.526 | -0.803 | -0.249 | 2e-04 |
| VO2max | Moderate (22-60) | Intensity | Moderate | 29 | 21 | -0.64 | -0.828 | -0.452 | 0 |
| VO2max | Moderate (22-60) | Intensity | Light | 14 | 11 | -0.754 | -1.123 | -0.386 | 1e-04 |
| VO2max | Moderate (22-60) | Intensity | Vigorous | 22 | 18 | -0.508 | -0.647 | -0.368 | 0 |
| VO2max | Moderate (22-60) | Health_Condition | Other | 15 | 10 | -0.652 | -1.012 | -0.292 | 4e-04 |
| VO2max | Moderate (22-60) | Health_Condition | Diabetes | 11 | 4 | -0.471 | -0.68 | -0.262 | 0 |
| VO2max | Moderate (22-60) | Health_Condition | Hypertensive | 30 | 20 | -0.67 | -0.856 | -0.484 | 0 |
| VO2max | Moderate (22-60) | Health_Condition | Metabolic | 8 | 6 | -0.448 | -0.634 | -0.263 | 0 |
| VO2max | Moderate (22-60) | Health_Condition | Healthy | 6 | 4 | -0.428 | -0.855 | -0.001 | 0.0497 |
| VO2max | Moderate (22-60) | Health_Condition | CVD | 6 | 3 | -0.984 | -1.79 | -0.179 | 0.0166 |
| VO2max | High (¿60) | Exercise_Type | Aerobic | 12 | 9 | -0.838 | -1.155 | -0.521 | 0 |
| VO2max | High (¿60) | Exercise_Type | Resistance | 3 | 3 | -0.631 | -1.23 | -0.031 | 0.0393 |
| VO2max | High (¿60) | Exercise_Location | Indoor | 15 | 11 | -0.813 | -1.09 | -0.535 | 0 |
| VO2max | High (¿60) | Intensity | Light | 3 | 2 | -0.497 | -0.763 | -0.231 | 2e-04 |
| VO2max | High (¿60) | Intensity | Moderate | 9 | 9 | -0.862 | -1.232 | -0.491 | 0 |
| VO2max | High (¿60) | Intensity | Vigorous | 3 | 3 | -0.729 | -1.165 | -0.292 | 0.0011 |
| VO2max | High (¿60) | Health_Condition | Diabetes | 4 | 2 | -0.524 | -0.72 | -0.328 | 0 |
| VO2max | High (¿60) | Health_Condition | Hypertensive | 7 | 7 | -1.175 | -1.523 | -0.826 | 0 |
| VO2max | High (¿60) | Health_Condition | Metabolic | 4 | 2 | -0.496 | -0.729 | -0.262 | 0 |

##### Table S11

**Table S11.** Regional unadjusted PM_2.5 coefficients with exposure characteristics.

| **Region** | **Outcome** | **N_Arms** | **N_Studies** | **Beta_PM25_log** | **SE_robust** | **P_robust** | **PM25_mean** | **PM25_range** |
|---|---|---|---|---|---|---|---|---|
| North America | Systolic BP (SMD) | 192 | 117 | 0.0377 | 0.07 | 0.594 | 15 | 3.6-34.7 |
| North America | Diastolic BP (SMD) | 194 | 117 | 0.0083 | 0.0687 | 0.9047 | 14.9 | 3.6-34.7 |
| North America | VO2max (SMD) | 75 | 47 | -0.0738 | 0.0662 | 0.2871 | 16.3 | 6.2-34.7 |
| North America | Heart Rate (SMD) | 86 | 57 | 0.0595 | 0.0619 | 0.3519 | 15.4 | 5.9-34.7 |
| Europe | Systolic BP (SMD) | 200 | 125 | 0.0658 | 0.0754 | 0.388 | 13.4 | 3.7-32.2 |
| Europe | Diastolic BP (SMD) | 198 | 121 | 0.1498 | 0.0782 | 0.0629 | 13.2 | 3.7-32.2 |
| Europe | VO2max (SMD) | 96 | 58 | -0.194 | 0.1607 | 0.2419 | 12.5 | 3.7-28.3 |
| Europe | Heart Rate (SMD) | 92 | 54 | -0.0327 | 0.0843 | 0.7032 | 13.7 | 3.7-28.3 |
| East Asia | Systolic BP (SMD) | 107 | 69 | 0.1879 | 0.0832 | 0.032 | 51 | 10.2-165.4 |
| East Asia | Diastolic BP (SMD) | 103 | 67 | 0.1355 | 0.0703 | 0.0642 | 50.6 | 10.2-165.4 |
| East Asia | VO2max (SMD) | 29 | 19 | -0.0093 | 0.0886 | 0.9192 | 50.7 | 10.2-165.4 |
| East Asia | Heart Rate (SMD) | 50 | 29 | 0.1416 | 0.0782 | 0.1057 | 46.9 | 14.8-165.4 |
| Latin America | Systolic BP (SMD) | 136 | 86 | 0.0123 | 0.0495 | 0.8051 | 27.6 | 5.8-146.2 |
| Latin America | Diastolic BP (SMD) | 130 | 83 | 0.0378 | 0.0594 | 0.5279 | 27.3 | 3.5-146.2 |
| Latin America | VO2max (SMD) | 52 | 30 | -0.0395 | 0.1388 | 0.78 | 35.6 | 6.3-67.5 |
| Latin America | Heart Rate (SMD) | 57 | 37 | 0.0491 | 0.0763 | 0.5264 | 31 | 6.6-58.2 |

#### Table S12

**Table S12.** Sensitivity analysis: effect of assumed pre–post correlation (r = 0.50, 0.70, 0.85) on pooled estimates.

| Outcome | k | r_primary | SMD_primary | CI_primary | SMD_r050 | SMD_r070 | SMD_r085 | Max_variation_pct |
|---|---|---|---|---|---|---|---|---|
| SBP | 705 | 0.7 | 0.4329 | [0.394, 0.472] | 0.4338 | 0.4329 | 0.4317 | 0.47 |
| DBP | 697 | 0.7 | 0.3719 | [0.334, 0.410] | 0.3728 | 0.3719 | 0.371 | 0.5 |
| VO2max | 277 | 0.65 | -0.5577 | [-0.623, -0.492] | -0.5616 | -0.5564 | -0.5521 | 1.72 |
| HR | 327 | 0.55 | 0.2665 | [0.218, 0.315] | 0.2671 | 0.2646 | 0.2625 | 1.72 |

#### Table S13

**Table S13.** Subgroup analyses: exercise mode pooled effects (a), PM_2.5 meta-regression within each mode (b), and exercise mode $\times$ PM_2.5 interaction tests (c).
*(a)*

| **Outcome** | **Mode** | **N_Arms** | **N_Studies** | **ES** | **CI_Lower** | **CI_Upper** | **SE** | **P** | **tau2** | **I2** |
|---|---|---|---|---|---|---|---|---|---|---|
| SBP | Aerobic | 273 | 198 | 0.4775 | 0.4105 | 0.5445 | 0.0342 | ¡0.001 | 0.2058 | 80.9 |
| SBP | Resistance | 219 | 171 | 0.4464 | 0.3886 | 0.5042 | 0.0295 | ¡0.001 | 0.1197 | 69 |
| SBP | Combined | 72 | 52 | 0.3941 | 0.3032 | 0.485 | 0.0464 | ¡0.001 | 0.0943 | 72.5 |
| SBP | Mind-Body | 13 | 12 | 0.5502 | 0.2205 | 0.88 | 0.1679 | 0.0074 | 0.3139 | 92.5 |
| SBP | Other | 103 | 82 | 0.2705 | 0.1784 | 0.3627 | 0.047 | ¡0.001 | 0.1484 | 74.1 |
| DBP | Aerobic | 267 | 193 | 0.4223 | 0.3643 | 0.4803 | 0.0296 | ¡0.001 | 0.1489 | 77.2 |
| DBP | Resistance | 219 | 171 | 0.3844 | 0.3273 | 0.4415 | 0.0291 | ¡0.001 | 0.1131 | 67.3 |
| DBP | Combined | 73 | 52 | 0.323 | 0.1951 | 0.4508 | 0.0652 | ¡0.001 | 0.2196 | 86.2 |
| DBP | Mind-Body | 13 | 12 | 0.5314 | 0.2346 | 0.8281 | 0.1512 | 0.0049 | 0.2501 | 90.9 |
| DBP | Other | 100 | 79 | 0.231 | 0.1186 | 0.3434 | 0.0574 | ¡0.001 | 0.2482 | 81.3 |
| VO2max | Aerobic | 139 | 96 | -0.6983 | -0.7977 | -0.599 | 0.0507 | ¡0.001 | 0.2147 | 75.4 |
| VO2max | Resistance | 55 | 45 | -0.4279 | -0.5193 | -0.3364 | 0.0467 | ¡0.001 | 0.0707 | 56.2 |
| VO2max | Combined | 32 | 23 | -0.5248 | -0.6701 | -0.3795 | 0.074 | ¡0.001 | 0.1032 | 67.4 |
| VO2max | Other | 44 | 33 | -0.314 | -0.4673 | -0.1606 | 0.0783 | ¡0.001 | 0.1588 | 73.1 |
| HR | Aerobic | 129 | 97 | 0.3607 | 0.2887 | 0.4327 | 0.0367 | ¡0.001 | 0.0913 | 57.1 |
| HR | Resistance | 95 | 75 | 0.2268 | 0.1542 | 0.2994 | 0.037 | ¡0.001 | 0.0549 | 44.1 |
| HR | Combined | 32 | 23 | 0.2657 | 0.1771 | 0.3543 | 0.0449 | ¡0.001 | 0.0169 | 23.2 |
| HR | Mind-Body | 7 | 6 | -0.1397 | -0.7421 | 0.4628 | 0.3112 | 0.6724 | 0.5617 | 96.7 |
| HR | Other | 53 | 42 | 0.1932 | 0.0719 | 0.3145 | 0.0619 | 0.0034 | 0.1112 | 64.2 |

*(b)*

| Outcome | Mode | N_Arms | N_Studies | PM25_Mean | Beta_PM25_log | SE | P |
|---|---|---|---|---|---|---|---|
| SBP | Aerobic | 273 | 198 | 26.4 | 0.1527 | 0.0489 | 0.0026 |
| SBP | Resistance | 219 | 171 | 22.6 | 0.0154 | 0.0356 | 0.6662 |
| SBP | Combined | 72 | 52 | 24.9 | 0.1109 | 0.0618 | 0.0944 |
| SBP | Mind-Body | 13 | 12 | 47.6 | 0.3995 | 0.1929 | 0.0985 |
| SBP | Other | 103 | 82 | 22.6 | -0.0455 | 0.0663 | 0.4998 |
| DBP | Aerobic | 267 | 193 | 26.1 | 0.093 | 0.038 | 0.017 |
| DBP | Resistance | 219 | 171 | 23.6 | 0.0376 | 0.0391 | 0.3412 |
| DBP | Combined | 73 | 52 | 24.9 | 0.0675 | 0.1227 | 0.5904 |
| DBP | Mind-Body | 13 | 12 | 47.6 | 0.3492 | 0.21 | 0.1625 |
| DBP | Other | 100 | 79 | 21.6 | -0.0249 | 0.0731 | 0.7372 |
| VO2max | Aerobic | 139 | 96 | 29.1 | -0.0515 | 0.0603 | 0.399 |
| VO2max | Resistance | 55 | 45 | 22.6 | -0.0391 | 0.0877 | 0.6652 |
| VO2max | Combined | 32 | 23 | 18 | -0.0927 | 0.1267 | 0.4859 |
| VO2max | Other | 44 | 33 | 19.9 | -0.117 | 0.2008 | 0.5773 |
| HR | Aerobic | 129 | 97 | 28.2 | 0.09 | 0.0445 | 0.0523 |
| HR | Resistance | 95 | 75 | 24.4 | 0.0295 | 0.0394 | 0.4624 |
| HR | Combined | 32 | 23 | 23.4 | 0.0754 | 0.0623 | 0.2843 |
| HR | Other | 53 | 42 | 21.4 | -0.0174 | 0.1443 | 0.9064 |

*(c)*

| Outcome | N_Arms | N_Studies | LRT_P |
|---|---|---|---|
| SBP | 564 | 396 | 0.0742 |
| DBP | 559 | 389 | 0.6362 |
| VO2max | 226 | 155 | 0.991 |
| HR | 256 | 181 | 0.5995 |

#### Table S14

**Table S14.** Clinical translation: standardised mean differences converted to absolute units with estimated cardiovascular risk reductions by PM_2.5 stratum.

| Outcome | PM25_Stratum | k | Studies | SMD | SMD_95CI | Abs_change | Abs_95CI | Units | RRR_MACE_pct | RRR_CHD_pct | RRR_stroke_pct | RRR_HF_pct | RRR_all_cause_mortality_pct |
|---|---|---|---|---|---|---|---|---|---|---|---|---|---|
| SBP | Low (¡22) | 485 | 300 | 0.375 | [0.333, 0.418] | 5.2 | [4.7, 5.9] | mmHg | 10.5 | 8.9 | 14.2 | 14.7 | |
| SBP | Moderate (22-60) | 170 | 108 | 0.51 | [0.426, 0.594] | 7.2 | [6.0, 8.3] | mmHg | 14.4 | 12.2 | 19.4 | 20.2 | |
| SBP | High (¿60) | 50 | 34 | 0.692 | [0.499, 0.886] | 9.7 | [7.0, 12.4] | mmHg | 19.4 | 16.5 | 26.2 | 27.1 | |
| DBP | Low (¡22) | 483 | 296 | 0.335 | [0.293, 0.377] | 3.0 | [2.6, 3.4] | mmHg | | | | | |
| DBP | Moderate (22-60) | 166 | 105 | 0.392 | [0.302, 0.482] | 3.5 | [2.7, 4.3] | mmHg | | | | | |
| DBP | High (¿60) | 48 | 34 | 0.616 | [0.460, 0.772] | 5.5 | [4.1, 6.9] | mmHg | | | | | |
| VO2max | Low (¡22) | 185 | 119 | -0.519 | [-0.598, -0.440] | 3.0 | [2.6, 3.5] | ml/kg/min | | | | | 10.2 |
| VO2max | Moderate (22-60) | 76 | 43 | -0.58 | [-0.696, -0.463] | 3.4 | [2.7, 4.0] | ml/kg/min | | | | | 11.4 |
| VO2max | High (¿60) | 16 | 12 | -0.876 | [-1.164, -0.589] | 5.1 | [3.4, 6.8] | ml/kg/min | | | | | 17.3 |
| HR | Low (¡22) | 216 | 134 | 0.25 | [0.188, 0.313] | 2.7 | [2.0, 3.3] | bpm | | | | | |
| HR | Moderate (22-60) | 90 | 58 | 0.266 | [0.192, 0.339] | 2.8 | [2.0, 3.6] | bpm | | | | | |
| HR | High (¿60) | 21 | 12 | 0.42 | [0.190, 0.651] | 4.5 | [2.0, 6.9] | bpm | | | | | |

#### Table S15

**Table S15.** Publication bias assessment: arm-level Egger and Begg tests (a); study-aggregated tests with trim-and-fill (b).

*(a)*

| **Outcome** | **k** | **Egger_z** | **Egger_p** | **Begg_tau** | **Begg_p** |
|---|---|---|---|---|---|
| SBP | 705 | 12 | ¡0.001 | 0.242 | ¡0.001 |
| DBP | 697 | 9.72 | ¡0.001 | 0.207 | ¡0.001 |
| VO2max | 277 | -6.5 | ¡0.001 | -0.216 | ¡0.001 |
| HR | 327 | 4.43 | ¡0.001 | 0.102 | 0.030 |

*(b)*

| Outcome | N_Studies_Agg | Egger_z | Egger_p | Begg_tau | Begg_p | TrimFill_k0 | TrimFill_ES_adj | TrimFill_CI_Lo | TrimFill_CI_Hi | FailsafeN_Rosenthal |
|---|---|---|---|---|---|---|---|---|---|---|
| Systolic BP (SMD) | 442 | 7.261 | 0 | 0.237 | 0 | 0 | 0.432 | 0.394 | 0.47 | NA |
| Diastolic BP (SMD) | 435 | 4.046 | 1e-04 | 0.181 | 0 | 0 | 0.367 | 0.329 | 0.404 | NA |
| VO2max (SMD) | 174 | -1.033 | 0.3029 | -0.239 | 0 | 0 | -0.555 | -0.621 | -0.488 | NA |
| Heart Rate (SMD) | 204 | 3.285 | 0.0012 | 0.029 | 0.5416 | 0 | 0.265 | 0.218 | 0.312 | NA |

#### Table S16

**Table S16.** Per-study risk-of-bias assessments (RoB 2 / ROBINS-I) with domain-level judgements and rationales for all 465 included studies. Provided as a separate CSV file (TableS16_rob_assessments.csv) owing to size.

#### Table S17

**Table S17.** City-level results for all 301 distinct study locations. Pooled SBP and DBP effect sizes are REML random-effects within location; single-arm locations report the arm-level estimate. Also provided as a separate CSV file.

| City | Country | Region | Latitude | Longitude | Studies | Arms | Participants | Mean_PM25 | SBP_k | SBP_SMD | SBP_95CI | DBP_k | DBP_SMD | DBP_95CI |
|---|---|---|---|---|---|---|---|---|---|---|---|---|---|---|
| Brisbane | Australia | Australasia | -27.47 | 153.02 | 1 | 1 | 68 | 11 | 1 | 0.377 | [0.182, 0.572] | 1 | -0.407 | [-0.604, -0.21 |
| Joondalup | Australia | Australasia | -31.78 | 115.76 | 1 | 3 | 33 | 7.1 | 3 | 0.258 | [-0.155, 0.670] | 3 | 0.163 | [-0.107, 0.432 |
| Melbourne | Australia | Australasia | -37.81 | 144.96 | 2 | 2 | 74 | 12.3 | 1 | 1.193 | [0.838, 1.548] | 2 | 1.631 | [1.060, 2.202] |
| NSW | Australia | Australasia | -31.88 | 147.29 | 2 | 4 | 68 | 4.1 | 4 | 0.22 | [-0.089, 0.530] | 4 | 0.344 | [0.150, 0.537] |
| Perth | Australia | Australasia | -31.96 | 115.86 | 1 | 3 | 48 | 6.7 | 3 | 0.169 | [-0.098, 0.436] | 3 | 0.155 | [-0.068, 0.379 |
| Perth and Bunbury | Australia | Australasia | -31.95 | 115.86 | 1 | 2 | 63 | 4 | 2 | -0.169 | [-0.362, 0.025] | 2 | -0.325 | [-0.668, 0.019 |
| Subiaco | Australia | Australasia | -31.95 | 115.82 | 1 | 1 | 23 | 8.6 | 1 | 0.483 | [0.137, 0.829] | 1 | -0.193 | [-0.515, 0.128 |
| Sydney | Australia | Australasia | -33.87 | 151.21 | 2 | 2 | 32 | 11.2 | 2 | 0.41 | [0.123, 0.697] | 2 | 0.857 | [0.516, 1.199] |
| Tasmania | Australia | Australasia | -42.04 | 146.64 | 1 | 1 | 43 | 7.2 | 1 | 0.2 | [-0.035, 0.436] | 1 | 0.233 | [-0.004, 0.469 |
| Townsville | Australia | Australasia | -19.26 | 146.82 | 1 | 1 | 78 | 5.4 | 1 | 0.53 | [0.339, 0.721] | 1 | 0.696 | [0.493, 0.900] |
| Victoria | Australia | Australasia | -36.6 | 144.68 | 2 | 5 | 203 | 5.6 | 5 | 0.395 | [0.186, 0.604] | 5 | 0.401 | [0.242, 0.560] |
| Bad Schallerbach | Austria | Europe | 48.23 | 13.92 | 1 | 1 | 570 | 14.6 | 1 | 0.1 | [0.036, 0.164] | 1 | 0.1 | [0.036, 0.164] |
| Department of Physical Medicine and Rehabilitation | Austria | Europe | 47.07 | 15.44 | 1 | 2 | 54 | 6.6 | 2 | 0.353 | [0.136, 0.570] | 2 | 0 | [-0.207, 0.207 |
| Innsbruck | Austria | Europe | 47.27 | 11.4 | 1 | 1 | 24 | 12.9 | 1 | 0.062 | [-0.248, 0.373] | 1 | 0.167 | [-0.147, 0.480 |
| Leipzig | Austria | Europe | 48.18 | 15.08 | 1 | 1 | 18 | 13.9 | 1 | 0.907 | [0.443, 1.372] | 0 | NA | |
| Vienna | Austria | Europe | 48.21 | 16.37 | 1 | 1 | 10 | 20.9 | 1 | 0.193 | [-0.294, 0.681] | 1 | 0.211 | [-0.278, 0.700 |
| Antwerp and Leuven | Belgium | Europe | 50.88 | 4.7 | 3 | 5 | 381 | 18.1 | 3 | 0.331 | [0.008, 0.654] | 1 | 0.108 | [-0.018, 0.234 |
| Hasselt | Belgium | Europe | 50.93 | 5.33 | 1 | 2 | 119 | 20.1 | 2 | -0.576 | [-0.733, -0.418] | 2 | 0.456 | [0.198, 0.713] |
| Alagoas | Brazil | Latin America | -9.66 | -36.65 | 1 | 1 | 11 | 6.8 | 1 | 0.249 | [-0.220, 0.719] | 1 | 0.595 | [0.074, 1.116] |
| Bahia | Brazil | Latin America | -12.29 | -41.93 | 1 | 2 | 40 | 3.5 | 0 | NA | | 1 | 2.67 | [1.776, 3.565] |
| Bauru | Brazil | Latin America | -22.32 | -49.07 | 1 | 2 | 38 | 12.7 | 2 | 0.742 | [0.444, 1.039] | 0 | NA | |
| Belo Horizonte | Brazil | Latin America | -19.92 | -43.95 | 1 | 3 | 115 | 18.5 | 3 | 0.131 | [-0.124, 0.386] | 3 | 0.018 | [-0.124, 0.161 |
| Brasilia | Brazil | Latin America | -10.33 | -53.2 | 3 | 6 | 83 | 13 | 6 | 0.533 | [-0.180, 1.245] | 6 | 0.135 | [-0.331, 0.602 |
| Cruz Alta | Brazil | Latin America | -28.64 | -53.61 | 1 | 1 | 10 | 7.5 | 1 | 1.232 | [0.509, 1.954] | 1 | 1.336 | [0.579, 2.093] |
| Cruz Alta | Brazil | Latin America | -28.65 | -53.61 | 1 | 1 | 48 | 5.8 | 1 | 0.738 | [0.474, 1.002] | 1 | 0.459 | [0.222, 0.697] |
| DF | Brazil | Latin America | -15.78 | -47.8 | 1 | 1 | 22 | 15.3 | 1 | 0.049 | [-0.275, 0.373] | 1 | -0.028 | [-0.351, 0.296 |
| Fortaleza | Brazil | Latin America | -3.73 | -38.52 | 1 | 2 | 28 | 8.1 | 2 | 0.702 | [0.360, 1.043] | 2 | 0.546 | [0.225, 0.866] |
| Iguatu | Brazil | Latin America | -6.36 | -39.3 | 1 | 1 | 10 | 6.9 | 0 | NA | | 0 | NA | |
| Juiz De Fora | Brazil | Latin America | -21.76 | -43.35 | 2 | 2 | 28 | 14.1 | 2 | 0.386 | [0.082, 0.690] | 2 | 0.278 | [-0.018, 0.574 |
| Londrina | Brazil | Latin America | -23.31 | -51.16 | 5 | 7 | 106 | 11.1 | 7 | 0.266 | [-0.460, 0.992] | 7 | 0.076 | [-0.505, 0.657 |
| Minas Gerais | Brazil | Latin America | -18.53 | -44.16 | 2 | 3 | 57 | 7.6 | 3 | 0.433 | [0.153, 0.714] | 3 | 0.382 | [0.169, 0.596] |
| Mogi Das Cruzes | Brazil | Latin America | -23.52 | -46.19 | 1 | 1 | 15 | 50.3 | 1 | 1.163 | [0.592, 1.735] | 1 | 2.269 | [1.367, 3.170] |
| Natal | Brazil | Latin America | -5.81 | -35.21 | 4 | 5 | 83 | 8.3 | 5 | 0.198 | [-0.172, 0.567] | 5 | 0.273 | [-0.151, 0.696 |
| Niteroi | Brazil | Latin America | -22.88 | -43.1 | 1 | 1 | 10 | 8.4 | 1 | 1.412 | [0.629, 2.196] | 1 | 0.203 | [-0.285, 0.691 |
| POA | Brazil | Latin America | -23.52 | -46.34 | 1 | 1 | 11 | 49.3 | 1 | 0.755 | [0.199, 1.311] | 1 | 0.058 | [-0.401, 0.516 |
| Paraiba | Brazil | Latin America | -7.12 | -36.72 | 1 | 1 | 13 | 11.4 | 0 | NA | | 0 | NA | |
| Parana | Brazil | Latin America | -24.48 | -51.81 | 3 | 4 | 60 | 7.4 | 4 | 0.595 | [0.371, 0.819] | 4 | 0.537 | [0.230, 0.844] |
| Pelotas | Brazil | Latin America | -31.77 | -52.34 | 1 | 2 | 150 | 6.8 | 2 | 1.055 | [0.883, 1.227] | 2 | -0.032 | [-1.576, 1.512 |
| Porto Alegre | Brazil | Latin America | -30.03 | -51.23 | 4 | 9 | 141 | 18.5 | 6 | 0.434 | [0.252, 0.616] | 6 | 0.293 | [0.117, 0.469] |
| Presidente Prudente | Brazil | Latin America | -22.12 | -51.39 | 1 | 1 | 10 | 8.4 | 0 | NA | | 0 | NA | |
| RS | Brazil | Latin America | -29.84 | -53.77 | 2 | 4 | 84 | 7.7 | 4 | 0.191 | [-0.390, 0.772] | 4 | 0.37 | [0.077, 0.663] |
| Recife | Brazil | Latin America | -8.06 | -34.88 | 3 | 5 | 93 | 10.9 | 5 | 0.847 | [0.195, 1.498] | 5 | 0.72 | [0.090, 1.351] |
| Ribeirao Preto | Brazil | Latin America | -21.18 | -47.81 | 1 | 4 | 61 | 13.8 | 4 | 0.159 | [-0.133, 0.451] | 4 | 0.135 | [-0.097, 0.367 |
| Rio Claro | Brazil | Latin America | -22.41 | -47.57 | 1 | 4 | 110 | 26.9 | 4 | 0.878 | [0.464, 1.293] | 4 | 1.674 | [0.763, 2.585] |
| Rio De Janeiro | Brazil | Latin America | -22.91 | -43.21 | 3 | 3 | 70 | 22.7 | 3 | 0.576 | [-0.101, 1.253] | 3 | 0.484 | [0.038, 0.929] |
| Santa Catarina | Brazil | Latin America | -27.06 | -51.11 | 1 | 1 | 13 | 13.3 | 1 | 0.655 | [0.165, 1.146] | 1 | 0.468 | [0.010, 0.926] |
| Santos | Brazil | Latin America | -23.93 | -46.33 | 1 | 1 | 22 | 38.5 | 1 | 0.784 | [0.386, 1.182] | 1 | 0.602 | [0.233, 0.972] |
| Sao Carlos | Brazil | Latin America | -21.88 | -47.86 | 1 | 1 | 10 | 15.1 | 1 | -0.366 | [-0.872, 0.141] | 1 | -0.228 | [-0.719, 0.262 |
| Sao Paulo | Brazil | Latin America | -23.55 | -46.63 | 28 | 44 | 933 | 50 | 42 | 0.478 | [0.316, 0.640] | 38 | 0.36 | [0.174, 0.547] |
| Uberaba | Brazil | Latin America | -19.75 | -47.94 | 1 | 1 | 10 | 8.8 | 1 | 0.369 | [-0.137, 0.876] | 1 | 0.435 | [-0.081, 0.952 |
| Vitoria | Brazil | Latin America | -20.32 | -40.34 | 1 | 2 | 21 | 9.8 | 2 | 0.769 | [0.081, 1.457] | 2 | 0.671 | [0.283, 1.060] |
| BC | Canada | North America | 55 | -125 | 2 | 7 | 97 | 28.4 | 7 | -0.131 | [-0.486, 0.225] | 7 | -0.323 | [-0.633, -0.01 |
| Guelph | Canada | North America | 43.55 | -80.25 | 1 | 1 | 17 | 14.2 | 1 | 0.909 | [0.430, 1.387] | 1 | 0.639 | [0.213, 1.065] |
| Hamilton | Canada | North America | 43.26 | -79.87 | 3 | 3 | 77 | 26.5 | 3 | 0.825 | [0.078, 1.573] | 3 | 0.36 | [0.147, 0.573] |
| Kelowna | Canada | North America | 49.89 | -119.5 | 1 | 1 | 20 | 27.7 | 1 | 0.48 | [0.109, 0.851] | 1 | 0.427 | [0.062, 0.791] |
| Kingston | Canada | North America | 44.23 | -76.49 | 1 | 4 | 22 | 9.7 | 4 | -0.319 | [-0.780, 0.142] | 4 | -0.395 | [-0.843, 0.054 |
| Kingston | Canada | North America | 44.31 | -76.43 | 1 | 2 | 20 | 11.2 | 2 | 0.721 | [0.313, 1.128] | 2 | 0.599 | [0.211, 0.987] |
| London | Canada | North America | 42.98 | -81.24 | 1 | 2 | 63 | 13.7 | 2 | 0.323 | [0.077, 0.569] | 2 | 0.428 | [0.193, 0.663] |
| Moncton | Canada | North America | 46.1 | -64.8 | 1 | 1 | 12 | 7.2 | 1 | 2.17 | [1.197, 3.142] | 1 | 0.93 | [0.355, 1.505] |
| Montreal | Canada | North America | 45.5 | -73.57 | 1 | 1 | 33 | 17.2 | 1 | 0.345 | [0.068, 0.622] | 1 | 0.325 | [0.050, 0.601] |
| NL | Canada | North America | 53.82 | -61.23 | 1 | 1 | 8 | 5.9 | 1 | 0.662 | [0.035, 1.289] | 1 | 0.327 | [-0.233, 0.887 |
| Ontario | Canada | North America | 50 | -86 | 10 | 17 | 336 | 6.2 | 17 | 0.327 | [0.219, 0.435] | 17 | 0.347 | [0.154, 0.539] |
| Ottawa | Canada | North America | 45.42 | -75.69 | 1 | 3 | 188 | 10.6 | 3 | 0.149 | [0.037, 0.261] | 3 | 0.119 | [-0.005, 0.242 |
| Quebec | Canada | North America | 52.48 | -71.83 | 1 | 2 | 20 | 6.6 | 2 | 0.406 | [-0.382, 1.195] | 2 | 0.274 | [-0.409, 0.957 |
| Quebec City | Canada | North America | 46.81 | -71.21 | 1 | 2 | 16 | 12.3 | 2 | 0.592 | [-0.131, 1.315] | 2 | 0.631 | [0.188, 1.074] |
| Saskatoon | Canada | North America | 52.13 | -106.66 | 1 | 4 | 92 | 8.1 | 4 | 0.047 | [-0.111, 0.206] | 4 | 0.109 | [-0.129, 0.346 |
| Toronto | Canada | North America | 43.65 | -79.38 | 1 | 1 | 16 | 17.7 | 1 | 0 | [-0.380, 0.380] | 1 | 1.249 | [0.673, 1.824] |

| City | Country | Region | Latitude | Longitude | Studies | Arms | Participants | Mean_PM25 | SBP_k | SBP_SMD | SBP_95CI | DBP_k | DBP_SMD | DBP_95CI |
|---|---|---|---|---|---|---|---|---|---|---|---|---|---|---|
| Vancouver | Canada | North America | 49.26 | -123.11 | 2 | 2 | 52 | 18.3 | 1 | 0.385 | [-0.005, 0.775] | 2 | 0.533 | [0.298, 0.767] |
| Vancouver | Canada | North America | 49.28 | -123.12 | 1 | 2 | 40 | 19.3 | 2 | 1.092 | [-0.161, 2.345] | 2 | 1.17 | [-0.240, 2.580 |
| Windsor | Canada | North America | 42.31 | -83.04 | 1 | 1 | 11 | 16.2 | 1 | 0.283 | [-0.189, 0.756] | 1 | -0.581 | [-1.099, -0.06 |
| Los Lagos | Chile | Latin America | -41.47 | -72.94 | 4 | 8 | 118 | 7.8 | 8 | 0.51 | [0.200, 0.819] | 7 | 0.099 | [-0.053, 0.251 |
| Osorno | Chile | Latin America | -40.57 | -73.14 | 1 | 1 | 14 | 8.2 | 1 | 0.513 | [0.065, 0.961] | 1 | -0.371 | [-0.800, 0.057 |
| Rio Bueno | Chile | Latin America | -40.33 | -72.96 | 1 | 2 | 28 | 7.7 | 2 | 0.64 | [0.308, 0.973] | 2 | 0.242 | [-0.054, 0.537 |
| Santiago | Chile | Latin America | -33.44 | -70.65 | 2 | 3 | 55 | 65.2 | 3 | 0.369 | [0.153, 0.585] | 3 | 0.583 | [0.320, 0.847] |
| Temuco | Chile | Latin America | -38.74 | -72.59 | 2 | 4 | 59 | 13.2 | 3 | 0.41 | [0.129, 0.691] | 3 | 0.93 | [-0.415, 2.276 |
| Hong Kong | China | East Asia | 22.35 | 114.18 | 1 | 2 | 76 | 130.5 | 2 | 1.163 | [0.824, 1.503] | 2 | 1.679 | [1.112, 2.245] |
| Shaanxi Province | China | East Asia | 35.59 | 109.3 | 1 | 1 | 325 | 98.4 | 1 | 0.482 | [0.390, 0.574] | 1 | 0.115 | [0.030, 0.200] |
| Shanghai | China | East Asia | 31.23 | 121.47 | 2 | 4 | 115 | 96.3 | 2 | 1.388 | [1.084, 1.693] | 2 | 0.731 | [-0.142, 1.604 |
| Tianjin | China | East Asia | 39.08 | 117.2 | 1 | 2 | 55 | 149.5 | 2 | 1.762 | [1.373, 2.151] | 2 | 0.875 | [-0.236, 1.986 |
| Xiamen | China | East Asia | 24.54 | 118.08 | 1 | 3 | 220 | 66.9 | 3 | 0.317 | [0.044, 0.591] | 3 | 0.269 | [-0.018, 0.557 |
| Copenhagen | Denmark | Europe | 55.68 | 12.57 | 3 | 5 | 168 | 6.5 | 5 | 0.456 | [-0.103, 1.016] | 5 | 0.444 | [-0.046, 0.933 |
| Copenhagen | Denmark | Europe | 55.69 | 12.57 | 3 | 5 | 114 | 9.1 | 5 | 0.028 | [-0.140, 0.196] | 5 | 0.166 | [0.018, 0.313] |
| Hellerup | Denmark | Europe | 55.73 | 12.57 | 2 | 2 | 36 | 8.9 | 1 | 0.052 | [-0.328, 0.432] | 1 | 0.312 | [-0.083, 0.706 |
| Cairo | Egypt | Middle East | 30.04 | 31.24 | 1 | 2 | 40 | 82.8 | 2 | 0.545 | [0.277, 0.813] | 2 | 0.797 | [0.500, 1.095] |
| Tartu | Estonia | Europe | 58.38 | 26.72 | 1 | 1 | 17 | 10.4 | 1 | 1.905 | [1.166, 2.643] | 1 | 1.428 | [0.823, 2.033] |
| Local Football Clubs | Faroe Islands | Europe | 62.01 | -6.77 | 2 | 2 | 172 | 13.8 | 2 | 0.753 | [-0.164, 1.670] | 2 | 0.413 | [0.109, 0.716] |
| Jyvaskyla | Finland | Europe | 62.24 | 25.75 | 2 | 4 | 182 | 6.3 | 4 | 0.443 | [0.121, 0.764] | 4 | 0.379 | [0.128, 0.629] |
| Turku | Finland | Europe | 60.45 | 22.27 | 2 | 2 | 26 | 7.5 | 2 | 0.139 | [-0.338, 0.615] | 2 | 0.003 | [-0.914, 0.921 |
| Ballan Mire | France | Europe | 47.34 | 0.61 | 1 | 1 | 12 | 12.9 | 1 | 0.38 | [-0.084, 0.844] | 1 | 0.098 | [-0.342, 0.538 |
| Grenoble | France | Europe | 45.19 | 5.74 | 1 | 2 | 23 | 10.7 | 2 | 0.099 | [-0.222, 0.420] | 2 | 0.298 | [-0.032, 0.628 |
| Osseja | France | Europe | 42.42 | 1.98 | 1 | 2 | 84 | 10.8 | 2 | 0.14 | [-0.316, 0.596] | 2 | 0.094 | [-0.281, 0.469 |
| Paris | France | Europe | 48.86 | 2.32 | 1 | 1 | 30 | 21.2 | 1 | -0.07 | [-0.347, 0.208] | 1 | 0 | [-0.277, 0.277 |
| Saint-Etienne | France | Europe | 45.44 | 4.39 | 1 | 1 | 21 | 23 | 0 | NA | | 0 | NA | |
| Toulouse | France | Europe | 43.6 | 1.44 | 1 | 1 | 153 | 18.1 | 1 | 0.471 | [0.338, 0.605] | 1 | 0.459 | [0.326, 0.592] |
| Berlin | Germany | Europe | 52.51 | 13.4 | 6 | 9 | 183 | 12.7 | 9 | 0.7 | [0.158, 1.241] | 9 | 0.413 | [0.190, 0.635] |
| Cologne | Germany | Europe | 50.94 | 6.96 | 2 | 2 | 36 | 15.3 | 2 | 0.001 | [-0.956, 0.958] | 2 | 0.23 | [-0.037, 0.497 |
| Freiberg | Germany | Europe | 50.92 | 13.34 | 1 | 2 | 72 | 17.1 | 2 | 0.486 | [0.216, 0.757] | 2 | 0.445 | [0.080, 0.811] |
| Hamburg | Germany | Europe | 53.55 | 10 | 1 | 2 | 24 | 8.5 | 2 | -0.084 | [-0.594, 0.426] | 2 | 0.087 | [-0.226, 0.400 |
| Heidelberg | Germany | Europe | 49.41 | 8.69 | 1 | 1 | 15 | 21.8 | 0 | NA | | 0 | NA | |
| Leipzig | Germany | Europe | 51.34 | 12.37 | 1 | 1 | 60 | 12.1 | 1 | 0 | [-0.196, 0.196] | 1 | 0.082 | [-0.114, 0.279 |
| Saarbrucken | Germany | Europe | 49.23 | 7 | 2 | 5 | 139 | 12.8 | 5 | -0.003 | [-0.195, 0.188] | 5 | 0.218 | [0.085, 0.350] |
| Serres | Greece | Europe | 41.09 | 23.55 | 1 | 1 | 23 | 14.4 | 1 | 1.073 | [0.630, 1.516] | 1 | 0.536 | [0.184, 0.889] |
| Thessaloniki | Greece | Europe | 40.64 | 22.94 | 1 | 1 | 38 | 10.5 | 1 | 1.232 | [0.861, 1.603] | 1 | 0.784 | [0.481, 1.086] |
| Shatin | Hong Kong | East Asia | 22.38 | 114.19 | 1 | 2 | 24 | 122.5 | 2 | -0.346 | [-0.671, -0.020] | 2 | 0.344 | [-0.184, 0.872 |
| Unspecified | Hong Kong | East Asia | 22.32 | 114.17 | 1 | 1 | 18 | 138.9 | 1 | 0.531 | [0.134, 0.929] | 1 | 0.705 | [0.279, 1.131] |
| Kaposvar | Hungary | Europe | 46.36 | 17.79 | 1 | 3 | 641 | 12.1 | 3 | 0.311 | [0.212, 0.409] | 3 | 0.761 | [0.687, 0.834] |
| Veszprem | Hungary | Europe | 47.09 | 17.91 | 1 | 2 | 50 | 15.6 | 2 | 1.273 | [0.943, 1.602] | 2 | 0.751 | [0.490, 1.013] |
| Amritsar | India | South Asia | 31.63 | 74.87 | 1 | 2 | 40 | 144.1 | 2 | 0.071 | [-0.276, 0.417] | 2 | 0.564 | [0.294, 0.835] |
| Haryana | India | South Asia | 29 | 76 | 1 | 2 | 40 | 98.7 | 2 | 0.157 | [-0.947, 1.260] | 2 | 0.176 | [-1.466, 1.819 |
| Mangalore | India | South Asia | 12.91 | 74.86 | 1 | 1 | 25 | 36.8 | 1 | 2.939 | [2.069, 3.808] | 1 | 1.425 | [0.927, 1.924] |
| New Delhi | India | South Asia | 28.61 | 77.21 | 1 | 1 | 15 | 283.4 | 0 | NA | | 1 | 1.041 | [0.500, 1.581] |
| West Bengal | India | South Asia | 23 | 87.69 | 1 | 1 | 45 | 145.5 | 1 | 0.796 | [0.516, 1.075] | 1 | 0.395 | [0.154, 0.636] |
| Ardebil Province | Iran | Middle East | 38.25 | 48.29 | 1 | 1 | 10 | 19.7 | 0 | NA | | 0 | NA | |
| Mashhad | Iran | Middle East | 36.3 | 59.61 | 1 | 4 | 48 | 29.4 | 4 | 0.411 | [0.090, 0.732] | 4 | 1.198 | [0.682, 1.715] |
| Rasht | Iran | Middle East | 37.28 | 49.58 | 1 | 2 | 21 | 23 | 2 | 1.938 | [1.258, 2.618] | 2 | 0.549 | [0.036, 1.061] |
| Jerusalem | Israel | Middle East | 31.78 | 35.23 | 1 | 1 | 46 | 37.2 | 1 | 0.369 | [0.133, 0.605] | 1 | 0.335 | [0.101, 0.569] |
| Zerifin | Israel | Middle East | 31.93 | 34.8 | 1 | 2 | 113 | 46.1 | 2 | 0.306 | [0.157, 0.454] | 2 | 0.329 | [0.180, 0.478] |
| Ancona | Italy | Europe | 43.48 | 13.22 | 1 | 2 | 60 | 18 | 2 | 0.835 | [0.019, 1.651] | 2 | 0.855 | [0.106, 1.604] |
| Bologna | Italy | Europe | 44.49 | 11.34 | 1 | 2 | 160 | 24.5 | 1 | 0.156 | [-0.015, 0.328] | 2 | 1.223 | [0.105, 2.341] |
| Chieti | Italy | Europe | 42.1 | 14.42 | 1 | 2 | 36 | 16.8 | 2 | 0.36 | [-0.109, 0.828] | 2 | 0.268 | [-0.193, 0.728 |
| Ferrara | Italy | Europe | 44.77 | 11.83 | 1 | 2 | 126 | 27.3 | 2 | 0.438 | [0.292, 0.584] | 2 | 0.247 | [0.015, 0.479] |
| Larino | Italy | Europe | 41.8 | 14.91 | 1 | 2 | 40 | 15.1 | 2 | 0.769 | [-0.413, 1.951] | 2 | 0.788 | [0.493, 1.084] |
| Mantua | Italy | Europe | 45.17 | 10.67 | 1 | 1 | 12 | 28.9 | 1 | -0.31 | [-0.765, 0.145] | 1 | 0 | [-0.438, 0.438 |
| Milan | Italy | Europe | 45.46 | 9.19 | 2 | 4 | 98 | 20.5 | 4 | 0.933 | [0.314, 1.552] | 4 | 0.744 | [0.318, 1.169] |
| Padova | Italy | Europe | 45.39 | 11.81 | 1 | 3 | 58 | 23.4 | 3 | 0.704 | [0.432, 0.976] | 3 | 0.557 | [0.220, 0.894] |
| Palmero | Italy | Europe | 44.97 | 7.55 | 1 | 1 | 189 | 31.3 | 1 | 0.958 | [0.811, 1.105] | 1 | 1.207 | [1.042, 1.371] |
| Perugia | Italy | Europe | 43.11 | 12.39 | 2 | 3 | 56 | 17 | 3 | 0.209 | [-0.078, 0.497] | 3 | 0.139 | [-0.069, 0.348 |
| Rome | Italy | Europe | 41.89 | 12.48 | 1 | 1 | 47 | 21.4 | 1 | 0.113 | [-0.110, 0.335] | 1 | 0.208 | [-0.018, 0.433 |
| Rome | Italy | Europe | 41.9 | 12.5 | 2 | 3 | 600 | 17.7 | 3 | 0.334 | [0.180, 0.488] | 3 | 0.293 | [0.151, 0.434] |
| Rovereto | Italy | Europe | 45.89 | 11.03 | 1 | 2 | 23 | 28.3 | 2 | 0.322 | [-0.011, 0.655] | 2 | 1.096 | [0.262, 1.931] |
| Sardinia | Italy | Europe | 40.09 | 9.03 | 1 | 1 | 18 | 14 | 1 | 0.62 | [0.209, 1.031] | 1 | 2.52 | [1.623, 3.418] |
| Verona | Italy | Europe | 45.44 | 10.99 | 1 | 2 | 30 | 22.4 | 2 | 0.402 | [0.106, 0.698] | 2 | 0.443 | [-0.015, 0.900 |
| Fukui | Japan | East Asia | 35.93 | 136.61 | 1 | 1 | 58 | 18.5 | 1 | 0.431 | [0.217, 0.646] | 1 | 0.228 | [0.024, 0.431] |
| Gunma | Japan | East Asia | 36.52 | 139.03 | 1 | 3 | 26 | 14.8 | 3 | 0.05 | [-0.415, 0.516] | 3 | 0.145 | [-0.158, 0.447 |

| City | Country | Region | Latitude | Longitude | Studies | Arms | Participants | Mean_PM25 | SBP_k | SBP_SMD | SBP_95CI | DBP_k | DBP_SMD | DBP_95CI |
|---|---|---|---|---|---|---|---|---|---|---|---|---|---|---|
| Hokkaido | Japan | East Asia | 43.46 | 143.33 | 1 | 1 | 9 | 10.2 | 1 | 0.542 | [-0.023, 1.106] | 1 | 0.226 | [-0.291, 0.742 |
| Ibaraki | Japan | East Asia | 36.29 | 140.47 | 1 | 2 | 66 | 22.3 | 0 | NA | | 0 | NA | |
| Kitakyushu | Japan | East Asia | 33.88 | 130.87 | 1 | 2 | 78 | 18.7 | 2 | 0.614 | [0.416, 0.811] | 2 | 0.134 | [-0.040, 0.307 |
| Kitakyushu | Japan | East Asia | 33.88 | 130.88 | 1 | 1 | 35 | 23.2 | 1 | 0.496 | [0.214, 0.777] | 1 | 0.362 | [0.091, 0.632] |
| Kobe | Japan | East Asia | 34.69 | 135.19 | 1 | 1 | 16 | 30.2 | 1 | -0.068 | [-0.448, 0.312] | 1 | -0.125 | [-0.507, 0.257 |
| Kure | Japan | East Asia | 34.24 | 132.56 | 1 | 1 | 65 | 18.4 | 1 | 0.198 | [0.006, 0.389] | 1 | 0.282 | [0.088, 0.477] |
| Kyoto | Japan | East Asia | 35.01 | 135.77 | 2 | 4 | 55 | 22.2 | 4 | 0.594 | [0.017, 1.171] | 4 | 0.399 | [-0.130, 0.928 |
| Matsumoto | Japan | East Asia | 36.24 | 137.97 | 1 | 2 | 21 | 14.4 | 2 | 0.307 | [-0.037, 0.652] | 2 | 0.755 | [0.353, 1.158] |
| Niigata | Japan | East Asia | 37.65 | 138.77 | 1 | 1 | 295 | 18.2 | 1 | 0.499 | [0.402, 0.596] | 1 | 0.471 | [0.375, 0.567] |
| Okinawa | Japan | East Asia | 26.57 | 128.03 | 1 | 1 | 30 | 15.1 | 1 | 0.682 | [0.355, 1.008] | 1 | 0.433 | [0.135, 0.731] |
| Osaka | Japan | East Asia | 34.69 | 135.5 | 1 | 2 | 38 | 26 | 2 | -0.132 | [-0.381, 0.117] | 2 | -0.267 | [-0.521, -0.01 |
| Saitama | Japan | East Asia | 35.98 | 139.42 | 1 | 1 | 31 | 19.8 | 1 | 0.402 | [0.112, 0.693] | 1 | 0.267 | [-0.013, 0.548 |
| Sapporo | Japan | East Asia | 43.06 | 141.35 | 1 | 2 | 131 | 13.5 | 2 | 0.483 | [0.176, 0.789] | 2 | 0.468 | [0.183, 0.752] |
| Tokushima and Utsunomiya | Japan | East Asia | 33.92 | 134.25 | 1 | 4 | 200 | 16 | 4 | 0.255 | [-0.056, 0.567] | 4 | 0.304 | [0.082, 0.527] |
| Tokyo | Japan | East Asia | 35.68 | 139.76 | 5 | 13 | 272 | 37.7 | 13 | 0.426 | [0.007, 0.845] | 13 | 0.432 | [0.039, 0.824] |
| Tsukuba | Japan | East Asia | 36.08 | 140.08 | 6 | 9 | 114 | 32.6 | 9 | 0.199 | [0.042, 0.356] | 9 | 0.229 | [0.083, 0.376] |
| Yokohama | Japan | East Asia | 35.45 | 139.63 | 1 | 1 | 50 | 21 | 1 | 1.047 | [0.750, 1.344] | 1 | 0.998 | [0.707, 1.288] |
| Vilnius | Lithuania | Europe | 54.69 | 25.28 | 1 | 1 | 84 | 11 | 1 | 0.293 | [0.121, 0.464] | 1 | 0.257 | [0.087, 0.427] |
| Mexico City | Mexico | Latin America | 19.32 | -99.15 | 1 | 2 | 587 | 146.2 | 2 | 0.511 | [0.210, 0.813] | 2 | 0.443 | [0.264, 0.622] |
| Maastricht | Netherlands | Europe | 50.85 | 5.69 | 1 | 2 | 92 | 22.6 | 2 | 0.595 | [0.415, 0.776] | 2 | 0.492 | [0.318, 0.665] |
| Nijmegen | Netherlands | Europe | 51.84 | 5.84 | 1 | 2 | 44 | 15.7 | 2 | 0.73 | [0.454, 1.005] | 2 | 0.587 | [0.326, 0.847] |
| Utrecht | Netherlands | Europe | 52.09 | 5.12 | 2 | 2 | 97 | 12.5 | 2 | 0.253 | [0.094, 0.411] | 2 | 0.266 | [0.107, 0.425] |
| Dunedin | New Zealand | Australasia | -45.87 | 170.5 | 2 | 2 | 37 | 5.5 | 2 | 0.253 | [-0.004, 0.509] | 2 | 0.174 | [-0.079, 0.428 |
| Wellington | New Zealand | Australasia | -41.29 | 174.78 | 1 | 1 | 36 | 5.3 | 1 | 0.652 | [0.358, 0.947] | 1 | 0.367 | [0.100, 0.634] |
| Wellington | New Zealand | Australasia | -41.32 | 174.81 | 1 | 1 | 50 | 4.7 | 1 | 0.136 | [-0.081, 0.352] | 1 | 0.085 | [-0.130, 0.300 |
| Ibadan | Nigeria | Africa | 7.38 | 3.9 | 2 | 2 | 75 | 66.3 | 2 | 2.195 | [1.323, 3.067] | 2 | 0.971 | [0.737, 1.205] |
| Kano | Nigeria | Africa | 11.99 | 8.52 | 4 | 4 | 324 | 88.5 | 4 | 1.1 | [0.980, 1.219] | 4 | 1.184 | [0.619, 1.749] |
| Minna | Nigeria | Africa | 9.61 | 6.56 | 1 | 3 | 357 | 47.4 | 3 | 0.672 | [-0.197, 1.541] | 3 | 0.709 | [0.514, 0.905] |
| Trondheim | Norway | Europe | 63.43 | 10.4 | 4 | 8 | 157 | 5.8 | 5 | 0.421 | [0.180, 0.663] | 8 | 0.729 | [0.387, 1.070] |
| Bydgoszcz | Poland | Europe | 53.13 | 18.03 | 1 | 2 | 55 | 11.4 | 2 | -0.304 | [-0.878, 0.270] | 2 | -0.359 | [-0.677, -0.04 |
| Glucholazy | Poland | Europe | 50.32 | 17.38 | 1 | 1 | 101 | 14.6 | 1 | 0.443 | [0.280, 0.606] | 1 | 0.289 | [0.132, 0.445] |
| Lodz | Poland | Europe | 51.77 | 19.46 | 1 | 1 | 12 | 14.5 | 1 | 0.266 | [-0.185, 0.717] | 1 | 0.344 | [-0.115, 0.804 |
| Poznan | Poland | Europe | 52.4 | 16.92 | 2 | 4 | 123 | 12.2 | 4 | 0.58 | [0.425, 0.736] | 4 | 0.457 | [0.183, 0.730] |
| Rzasnia | Poland | Europe | 51.22 | 19.04 | 1 | 2 | 53 | 15.3 | 2 | 0 | [-0.209, 0.209] | 2 | 0.083 | [-0.126, 0.292 |
| Coimbra | Portugal | Europe | 40.2 | -8.41 | 1 | 3 | 63 | 17.2 | 3 | 0.068 | [-0.125, 0.260] | 3 | 0.165 | [-0.029, 0.359 |
| Covilha | Portugal | Europe | 40.28 | -7.5 | 1 | 1 | 39 | 32.2 | 1 | 0.878 | [0.567, 1.190] | 1 | 0.558 | [0.285, 0.831] |
| Lisbon | Portugal | Europe | 38.71 | -9.14 | 1 | 3 | 51 | 9 | 3 | 0.396 | [0.169, 0.622] | 3 | 0.353 | [0.128, 0.578] |
| Maia | Portugal | Europe | 41.24 | -8.62 | 1 | 2 | 31 | 18.9 | 2 | 1.002 | [0.072, 1.932] | 2 | 0.904 | [0.500, 1.309] |
| Porto | Portugal | Europe | 41.16 | -8.63 | 1 | 1 | 20 | 3.7 | 1 | 0.291 | [-0.060, 0.642] | 1 | 0.208 | [-0.138, 0.553 |
| University of Porto | Portugal | Europe | 41.18 | -8.6 | 1 | 2 | 31 | 18.9 | 2 | 0.601 | [0.290, 0.912] | 2 | 0.599 | [0.288, 0.910] |
| Vila Nova De Gaia | Portugal | Europe | 41.13 | -8.61 | 1 | 1 | 25 | 19.4 | 1 | 0.161 | [-0.146, 0.468] | 1 | 0.484 | [0.152, 0.816] |
| Vilarinho | Portugal | Europe | 41.71 | -8.41 | 1 | 1 | 16 | 10.1 | 1 | 0.322 | [-0.074, 0.718] | 1 | 0.302 | [-0.092, 0.695 |
| Moscow | Russia | Europe | 55.63 | 37.61 | 1 | 2 | 29 | 23.6 | 2 | 0.341 | [-0.182, 0.864] | 2 | 0.114 | [-0.893, 1.121 |
| Unspecified | Singapore | South/Southeast Asia | 1.28 | 103.85 | 1 | 2 | 60 | 41.3 | 2 | 0.342 | [-0.340, 1.024] | 2 | 0.174 | [-0.173, 0.521 |
| Gauteng | South Africa | Africa | -25.94 | 28.08 | 1 | 1 | 183 | 123.4 | 0 | NA | | 0 | NA | |
| Johannesburg | South Africa | Africa | -26.2 | 28.05 | 2 | 2 | 270 | 149.8 | 2 | 0.154 | [-0.108, 0.416] | 2 | 0.227 | [0.007, 0.446] |
| Busan | South Korea | East Asia | 35.13 | 129.1 | 4 | 5 | 76 | 56 | 5 | 0.411 | [0.046, 0.775] | 5 | 0.232 | [-0.071, 0.535 |
| Busan | South Korea | East Asia | 35.18 | 129.08 | 5 | 5 | 99 | 58.3 | 5 | 0.747 | [0.199, 1.295] | 4 | 0.292 | [-0.216, 0.799 |
| Daegu | South Korea | East Asia | 35.86 | 128.58 | 1 | 2 | 55 | 52.4 | 2 | 0.023 | [-0.182, 0.228] | 2 | -0.107 | [-0.313, 0.099 |
| Daegu | South Korea | East Asia | 35.87 | 128.6 | 1 | 2 | 30 | 45 | 2 | 0.211 | [-0.328, 0.750] | 2 | -0.133 | [-0.412, 0.147 |
| Gwangju/Jeonju | South Korea | East Asia | 35.16 | 126.85 | 1 | 1 | 15 | 36 | 1 | 0.61 | [0.161, 1.059] | 1 | 0.488 | [0.059, 0.917] |
| Incheon | South Korea | East Asia | 37.51 | 126.72 | 1 | 2 | 16 | 120.6 | 2 | 1.145 | [-0.371, 2.661] | 2 | 0.833 | [-0.031, 1.696 |
| Manufacturing Workplace Departments | South Korea | East Asia | 37.57 | 126.98 | 4 | 6 | 314 | 97.2 | 6 | 0.349 | [0.097, 0.602] | 3 | 0.231 | [-0.110, 0.572 |
| Metropolitan City | South Korea | East Asia | -26.04 | 27.93 | 2 | 2 | 45 | 165 | 2 | 0.466 | [-0.112, 1.044] | 2 | 0.727 | [0.456, 0.999] |
| Barcelona | Spain | Europe | 41.38 | 2.18 | 1 | 4 | 39 | 10.6 | 0 | NA | | 0 | NA | |
| Basque Country | Spain | Europe | 42.99 | -2.55 | 2 | 5 | 357 | 8.6 | 5 | 0.55 | [0.434, 0.666] | 4 | 0.577 | [0.452, 0.701] |
| Catalonia | Spain | Europe | 41.85 | 1.57 | 2 | 2 | 412 | 11.1 | 2 | 0.203 | [0.127, 0.279] | 2 | 0.268 | [0.132, 0.404] |
| Elche | Spain | Europe | 38.27 | -0.7 | 1 | 1 | 10 | 9.6 | 1 | 0.442 | [-0.075, 0.960] | 1 | 0.336 | [-0.166, 0.838 |
| Granada | Spain | Europe | 37.17 | -3.6 | 1 | 1 | 60 | 11.8 | 1 | 0.77 | [0.530, 1.010] | 1 | 0.443 | [0.231, 0.654] |
| Madrid | Spain | Europe | 40.42 | -3.7 | 2 | 2 | 212 | 12.6 | 2 | 0.222 | [-0.327, 0.770] | 2 | 0.046 | [-0.619, 0.712 |
| Malaga | Spain | Europe | 36.72 | -4.42 | 1 | 2 | 21 | 13.2 | 2 | 0.332 | [-0.299, 0.964] | 2 | 0.338 | [-0.104, 0.781 |
| Mallorca | Spain | Europe | 39.61 | 2.88 | 1 | 2 | 23 | 7.1 | 2 | 0.794 | [-0.178, 1.766] | 2 | 0.434 | [-1.075, 1.943 |
| Murcia | Spain | Europe | 37.99 | -1.13 | 1 | 3 | 41 | 10.1 | 3 | 0.451 | [0.193, 0.710] | 3 | -0.191 | [-1.244, 0.862 |
| Reus-Tarragona | Spain | Europe | 41.16 | 1.11 | 1 | 1 | 260 | 11 | 1 | 0.249 | [0.153, 0.346] | 1 | 0.294 | [0.196, 0.391] |
| Toledo | Spain | Europe | 39.86 | -4.02 | 4 | 7 | 153 | 9.9 | 4 | 0.733 | [0.014, 1.451] | 4 | 0.991 | [0.223, 1.760] |
| Toledo | Spain | Europe | 39.86 | -4.03 | 1 | 1 | 26 | 10.2 | 1 | 0.071 | [-0.228, 0.369] | 1 | 0.051 | [-0.247, 0.349 |
| Unspecified | Spain | Europe | 33.18 | -86.81 | 1 | 1 | 55 | 11.6 | 1 | -0.256 | [-0.467, -0.046] | 1 | 0.099 | [-0.107, 0.304 |

| City | Country | Region | Latitude | Longitude | Studies | Arms | Participants | Mean_PM25 | SBP_k | SBP_SMD | SBP_95CI | DBP_k | DBP_SMD | DBP_95CI |
|---|---|---|---|---|---|---|---|---|---|---|---|---|---|---|
| Valladolid | Spain | Europe | 41.65 | -4.73 | 1 | 2 | 107 | 10.3 | 2 | 0.573 | [0.408, 0.739] | 2 | -0.187 | [-0.336, -0.03 |
| Vitoria-Gasteiz | Spain | Europe | 42.85 | -2.67 | 2 | 7 | 216 | 9.3 | 7 | 0.441 | [0.265, 0.617] | 7 | 0.467 | [0.304, 0.629] |
| Gothenburg | Sweden | Europe | 57.71 | 11.97 | 1 | 3 | 74 | 8.9 | 3 | 0.101 | [-0.077, 0.280] | 3 | 0.096 | [-0.299, 0.490 |
| Gustavsberg and Stockholm | Sweden | Europe | 59.33 | 18.07 | 2 | 4 | 113 | 7.9 | 4 | 0.152 | [0.007, 0.297] | 4 | 0.074 | [-0.225, 0.374 |
| Jarfalla | Sweden | Europe | 59.42 | 17.83 | 1 | 1 | 22 | 8.6 | 1 | -0.172 | [-0.500, 0.156] | 1 | -0.207 | [-0.536, 0.123 |
| Linkoping | Sweden | Europe | 58.41 | 15.62 | 1 | 2 | 42 | 7.1 | 0 | NA |  | 0 | NA |  |
| Norrkoping | Sweden | Europe | 58.59 | 16.19 | 1 | 2 | 53 | 7.1 | 2 | 0.238 | [0.024, 0.451] | 2 | 0.261 | [0.047, 0.476] |
| Basel | Switzerland | Europe | 47.56 | 7.59 | 1 | 2 | 88 | 14.6 | 2 | 0.923 | [0.056, 1.789] | 2 | 0.34 | [0.051, 0.629] |
| Zurich | Switzerland | Europe | 47.37 | 8.54 | 1 | 1 | 9 | 10.8 | 1 | -0.737 | [-1.347, -0.127] | 1 | 0.301 | [-0.224, 0.826 |
| Chia Yi | Taiwan | East Asia | 23.49 | 120.46 | 1 | 3 | 154 | 37.8 | 3 | 0.201 | [0.076, 0.326] | 3 | 0.01 | [-0.142, 0.162 |
| Fooyin | Taiwan | East Asia | 22.61 | 120.39 | 1 | 1 | 51 | 20.1 | 1 | 0.409 | [0.182, 0.636] | 1 | 0.352 | [0.128, 0.575] |
| Taichung | Taiwan | East Asia | 24.16 | 120.65 | 1 | 1 | 33 | 49 | 1 | 0.337 | [0.061, 0.614] | 1 | 0.246 | [-0.024, 0.517 |
| Tainan | Taiwan | East Asia | 22.99 | 120.18 | 1 | 1 | 34 | 36.1 | 1 | 0.533 | [0.243, 0.822] | 1 | 0.661 | [0.357, 0.965] |
| Taipei | Taiwan | East Asia | 25.03 | 121.57 | 2 | 3 | 532 | 42.9 | 3 | 0.685 | [-0.081, 1.450] | 3 | 0.382 | [-0.019, 0.783 |
| Taipei | Taiwan | East Asia | 25.04 | 121.56 | 5 | 5 | 339 | 44.4 | 5 | 0.681 | [0.175, 1.186] | 5 | -0.042 | [-0.943, 0.859 |
| Taipei City | Taiwan | East Asia | 25.03 | 121.42 | 2 | 3 | 166 | 38 | 3 | 0.507 | [0.096, 0.919] | 3 | 0.406 | [0.062, 0.750] |
| Taoyuan | Taiwan | East Asia | 24.99 | 121.3 | 1 | 2 | 904 | 38.7 | 2 | 0.39 | [0.336, 0.443] | 2 | 0.443 | [0.388, 0.497] |
| Bangkok | Thailand | South/Southeast Asia | 13.75 | 100.49 | 2 | 5 | 69 | 76.3 | 5 | 0.368 | [0.116, 0.620] | 5 | 0.164 | [-0.158, 0.487 |
| Kef and Ksar-Said | Tunisia | Middle East | 36.81 | 10.18 | 1 | 2 | 32 | 14.2 | 2 | 0.262 | [-0.015, 0.538] | 2 | 1.082 | [0.704, 1.460] |
| Adiyaman | Turkey | Middle East | 37.76 | 38.28 | 1 | 1 | 16 | 19.8 | 1 | 0.201 | [-0.185, 0.587] | 1 | 2.757 | [1.729, 3.784] |
| Ankara | Turkey | Middle East | 39.92 | 32.85 | 2 | 2 | 32 | 15.2 | 2 | 0.877 | [0.531, 1.223] | 2 | 0.633 | [-0.417, 1.683 |
| Istanbul | Turkey | Middle East | 41.01 | 28.98 | 1 | 1 | 14 | 16.9 | 1 | 0.871 | [0.353, 1.390] | 1 | 0.426 | [-0.009, 0.862 |
| Izmir | Turkey | Middle East | 38.42 | 27.13 | 1 | 1 | 24 | 14.6 | 1 | 0.326 | [0.003, 0.650] | 1 | -0.02 | [-0.330, 0.289 |
| Cambridge | UK | Europe | 52.21 | 0.12 | 1 | 2 | 100 | 13.3 | 2 | 0.167 | [0.013, 0.320] | 2 | 0.147 | [-0.007, 0.300 |
| Canterbury | UK | Europe | 51.28 | 1.08 | 2 | 2 | 52 | 9.8 | 2 | 1.31 | [-0.226, 2.846] | 2 | 0.703 | [0.427, 0.979] |
| Cardiff | UK | Europe | 51.48 | -3.18 | 1 | 1 | 22 | 12.9 | 1 | 0.482 | [0.128, 0.836] | 1 | 0.535 | [0.175, 0.896] |
| Derby | UK | Europe | 52.92 | -1.48 | 1 | 1 | 17 | 15.5 | 1 | 2.857 | [1.828, 3.885] | 0 | NA |  |
| Dudley | UK | Europe | 52.51 | -2.08 | 1 | 1 | 18 | 14.1 | 1 | 0.557 | [0.156, 0.959] | 1 | 0.318 | [-0.054, 0.691 |
| Dundee | UK | Europe | 56.46 | -2.97 | 1 | 1 | 22 | 9.7 | 1 | 0.081 | [-0.243, 0.406] | 1 | -0.093 | [-0.417, 0.232 |
| Glasgow | UK | Europe | 55.86 | -4.25 | 2 | 3 | 44 | 10.1 | 3 | 0.266 | [-0.087, 0.619] | 3 | 0.543 | [0.122, 0.964] |
| Home-Based Intervention | UK | Europe | 51.51 | -0.13 | 1 | 1 | 74 | 14.7 | 1 | -0.045 | [-0.221, 0.132] | 1 | -0.082 | [-0.259, 0.095 |
| Leeds | UK | Europe | 53.8 | -1.54 | 1 | 1 | 58 | 16.2 | 1 | 0.493 | [0.275, 0.712] | 1 | 0.789 | [0.544, 1.035] |
| Leicester | UK | Europe | 52.64 | -1.13 | 1 | 1 | 14 | 14.7 | 1 | 0.183 | [-0.228, 0.594] | 0 | NA |  |
| Liverpool | UK | Europe | 53.41 | -2.99 | 1 | 1 | 14 | 11.2 | 0 | NA |  | 0 | NA |  |
| Middlesex | UK | Europe | 51.55 | -0.25 | 1 | 1 | 418 | 11.8 | 1 | -0.035 | [-0.109, 0.039] | 1 | 0.092 | [0.017, 0.166] |
| Sheffield | UK | Europe | 53.38 | -1.47 | 2 | 2 | 64 | 15.2 | 2 | 0.327 | [0.128, 0.527] | 2 | 0.215 | [-0.502, 0.931 |
| Unspecified | UK | Europe | 53.38 | -1.49 | 1 | 1 | 50 | 17.7 | 1 | 0.222 | [0.003, 0.441] | 1 | 0.384 | [0.157, 0.612] |
| Wales | UK | Europe | 52.29 | -3.74 | 1 | 1 | 10 | 9.6 | 1 | 0.46 | [-0.061, 0.981] | 1 | 0.248 | [-0.244, 0.740 |
| AL | USA | North America | 33.26 | -86.83 | 1 | 2 | 28 | 13.3 | 2 | 0.167 | [-0.135, 0.469] | 2 | 0.192 | [-0.631, 1.015 |
| Amherst | USA | North America | 42.37 | -72.52 | 2 | 6 | 78 | 18.2 | 5 | 0.171 | [-0.181, 0.523] | 6 | 0.604 | [-0.223, 1.431 |
| Athens | USA | North America | 33.96 | -83.38 | 1 | 1 | 32 | 14.9 | 1 | -0.046 | [-0.315, 0.222] | 1 | -0.201 | [-0.474, 0.072 |
| Austin | USA | North America | 30.27 | -97.74 | 1 | 2 | 48 | 10.9 | 2 | 0.193 | [-0.194, 0.580] | 2 | 0.097 | [-0.124, 0.318 |
| Baltimore | USA | North America | 39.29 | -76.61 | 5 | 6 | 301 | 23.5 | 6 | 0.44 | [0.264, 0.617] | 5 | 0.334 | [0.099, 0.568] |
| Baton Rouge | USA | North America | 30.45 | -91.19 | 1 | 3 | 132 | 14.7 | 3 | -0.103 | [-0.293, 0.086] | 3 | -0.231 | [-0.366, -0.09 |
| Birmingham | USA | North America | 33.52 | -86.8 | 2 | 3 | 431 | 13.7 | 3 | 0.196 | [0.095, 0.296] | 3 | 0.245 | [0.158, 0.333] |
| Bloomington | USA | North America | 39.17 | -86.53 | 1 | 3 | 71 | 19.8 | 3 | -0.01 | [-0.191, 0.172] | 3 | -0.478 | [-1.039, 0.084 |
| Boulder | USA | North America | 40.01 | -105.27 | 1 | 2 | 113 | 29.2 | 2 | 0.209 | [0.063, 0.354] | 2 | 0.106 | [-0.464, 0.675 |
| CA | USA | North America | 36.7 | -118.76 | 1 | 1 | 31 | 22.7 | 1 | 0.308 | [0.025, 0.591] | 1 | 0.217 | [-0.061, 0.495 |
| CO | USA | North America | 38.73 | -105.61 | 1 | 2 | 208 | 6.7 | 2 | 0.281 | [0.173, 0.390] | 2 | 0.392 | [0.280, 0.504] |
| Champaign | USA | North America | 40.12 | -88.24 | 2 | 3 | 86 | 12.1 | 3 | 0.07 | [-0.113, 0.253] | 3 | 0.201 | [-0.013, 0.416 |
| Chicago | USA | North America | 41.88 | -87.62 | 2 | 3 | 32 | 20.7 | 3 | 0.245 | [-0.397, 0.886] | 3 | 0.407 | [0.028, 0.785] |
| Cleveland | USA | North America | 41.5 | -81.69 | 1 | 1 | 14 | 14.3 | 1 | 2.947 | [1.782, 4.111] | 0 | NA |  |
| Columbus | USA | North America | 39.96 | -83 | 1 | 1 | 47 | 16.7 | 1 | 0.251 | [0.024, 0.479] | 1 | 0.231 | [0.005, 0.457] |
| Cullowhee | USA | North America | 35.31 | -83.18 | 1 | 1 | 25 | 16.4 | 1 | 0 | [-0.304, 0.304] | 1 | 0.101 | [-0.204, 0.406 |
| Dallas | USA | North America | 32.78 | -96.8 | 3 | 5 | 603 | 16.2 | 5 | 0.126 | [-0.015, 0.266] | 5 | -0.042 | [-0.104, 0.020 |
| Detroit/Livonia/Clinton Township | USA | North America | 42.33 | -83.05 | 1 | 2 | 28 | 13.6 | 2 | 0.196 | [-0.343, 0.736] | 2 | 0.36 | [-0.361, 1.080 |
| Durham | USA | North America | 36 | -78.9 | 2 | 3 | 130 | 12.2 | 3 | 0.677 | [-0.576, 1.930] | 3 | 0.541 | [-0.275, 1.357 |
| Eau Claire | USA | North America | 44.81 | -91.5 | 1 | 2 | 16 | 12.9 | 2 | 0.198 | [-0.189, 0.585] | 2 | 0.232 | [-0.156, 0.620 |
| FL | USA | North America | 27.76 | -81.46 | 4 | 8 | 94 | 13 | 6 | 0.357 | [0.098, 0.616] | 8 | 0.524 | [0.082, 0.966] |
| Flagstaff | USA | North America | 35.2 | -111.65 | 1 | 1 | 11 | 10.5 | 1 | 0.278 | [-0.194, 0.751] | 1 | 0.185 | [-0.279, 0.650 |
| Gainesville | USA | North America | 29.65 | -82.32 | 4 | 5 | 66 | 19.1 | 5 | 0.39 | [-0.016, 0.795] | 5 | 0.202 | [-0.081, 0.485 |
| Grand Rapids | USA | North America | 42.96 | -85.67 | 1 | 2 | 22 | 13.1 | 1 | 0.777 | [0.261, 1.294] | 2 | 0.743 | [0.333, 1.152] |
| Gunnison | USA | North America | 38.65 | -107.06 | 1 | 2 | 22 | 7.2 | 2 | 0.841 | [0.264, 1.419] | 2 | 0.034 | [-0.462, 0.531 |
| Hines | USA | North America | 41.85 | -87.84 | 1 | 1 | 20 | 24.1 | 1 | 0.583 | [0.198, 0.967] | 1 | 0.288 | [-0.063, 0.639 |
| Iowa City | USA | North America | 41.66 | -91.53 | 1 | 1 | 12 | 11.3 | 1 | 0.322 | [-0.135, 0.779] | 1 | 0.268 | [-0.183, 0.720 |
| KS | USA | North America | 39.05 | -95.68 | 1 | 2 | 12 | 13.7 | 2 | 1.155 | [0.518, 1.791] | 1 | 1.414 | [0.477, 2.351] |
| Kansas City | USA | North America | 39.1 | -94.58 | 2 | 2 | 23 | 12.1 | 2 | 0.357 | [-1.290, 2.005] | 2 | -0.092 | [-0.451, 0.267 |

| City | Country | Region | Latitude | Longitude | Studies | Arms | Participants | Mean_PM25 | SBP_k | SBP_SMD | SBP_95CI | DBP_k | DBP_SMD | DBP_95CI |
|---|---|---|---|---|---|---|---|---|---|---|---|---|---|---|
| Kingston | USA | North America | 44.31 | -76.43 | 1 | 1 | 15 | 10.4 | 1 | 0.49 | [0.061, 0.920] | 1 | 0.473 | [0.046, 0.900] |
| Knoxville | USA | North America | 35.96 | -83.92 | 1 | 1 | 38 | 18.8 | 1 | 0.075 | [-0.172, 0.322] | 1 | 0.28 | [0.026, 0.534] |
| Los Angeles | USA | North America | 34.05 | -118.24 | 2 | 3 | 90 | 29.2 | 3 | 0.472 | [0.057, 0.886] | 3 | 0.544 | [0.317, 0.771] |
| Louisiana | USA | North America | 30.87 | -92.01 | 1 | 1 | 23 | 16.6 | 1 | -0.132 | [-0.450, 0.187] | 1 | -0.285 | [-0.612, 0.042 |
| MA | USA | North America | 42.36 | -71.06 | 1 | 3 | 121 | 15.6 | 3 | 0.138 | [-0.049, 0.325] | 3 | 0.308 | [-0.019, 0.636 |
| MI | USA | North America | 43.62 | -84.68 | 1 | 2 | 413 | 11.1 | 2 | 0.513 | [0.431, 0.596] | 2 | 0.443 | [0.362, 0.523] |
| Maryland | USA | North America | 39.52 | -76.94 | 2 | 2 | 21 | 12.4 | 2 | 0.213 | [-0.126, 0.553] | 2 | 0.184 | [-0.179, 0.548 |
| Miami | USA | North America | 25.77 | -80.19 | 1 | 1 | 89 | 19.8 | 1 | 0.248 | [0.083, 0.413] | 1 | 0.441 | [0.267, 0.614] |
| Miami-Dade County | USA | North America | 25.64 | -80.5 | 1 | 1 | 106 | 13.2 | 1 | 0.253 | [0.102, 0.405] | 1 | 0.225 | [0.074, 0.375] |
| Minneapolis | USA | North America | 44.98 | -93.27 | 1 | 1 | 16 | 15.7 | 1 | 1.299 | [0.710, 1.887] | 1 | 0.846 | [0.366, 1.325] |
| Morgantown | USA | North America | 39.63 | -79.96 | 1 | 1 | 11 | 13.2 | 1 | 0.923 | [0.324, 1.521] | 1 | 0.923 | [0.324, 1.521] |
| NC | USA | North America | 35.78 | -78.64 | 2 | 3 | 37 | 12.8 | 3 | 0.001 | [-0.251, 0.254] | 3 | 0.056 | [-0.196, 0.309 |
| Norman | USA | North America | 35.22 | -97.44 | 1 | 2 | 19 | 10.1 | 0 | NA |  | 0 | NA |  |
| North Carolina | USA | North America | 35.67 | -79.04 | 1 | 2 | 79 | 11.4 | 2 | 0.132 | [-0.041, 0.304] | 2 | 0.062 | [-0.110, 0.233 |
| PA | USA | North America | 35.68 | 139.76 | 1 | 1 | 12 | 23.9 | 1 | 0.857 | [0.300, 1.413] | 1 | 0.213 | [-0.234, 0.659 |
| Philadelphia | USA | North America | 39.95 | -75.16 | 1 | 2 | 37 | 13.4 | 2 | 0.084 | [-0.168, 0.335] | 2 | 0.044 | [-0.208, 0.295 |
| Phoenix | USA | North America | 33.45 | -112.07 | 1 | 2 | 16 | 11.8 | 2 | 0.155 | [-0.517, 0.826] | 2 | 0.179 | [-0.211, 0.570 |
| Pittsburgh | USA | North America | 40.44 | -80 | 1 | 2 | 130 | 18.4 | 2 | 1.332 | [1.123, 1.542] | 2 | 0.708 | [0.550, 0.867] |
| Rhode Island | USA | North America | 41.82 | -71.41 | 1 | 1 | 96 | 3.6 | 1 | 0.555 | [0.381, 0.729] | 1 | 0.259 | [0.100, 0.418] |
| Rochester | USA | North America | 43.16 | -77.62 | 1 | 1 | 58 | 10.7 | 1 | 0.079 | [-0.121, 0.279] | 1 | 0.417 | [0.204, 0.630] |
| Royal Oak | USA | North America | 42.49 | -83.14 | 1 | 1 | 15 | 12.8 | 1 | -0.008 | [-0.400, 0.384] | 1 | 0.732 | [0.260, 1.203] |
| Salisbury | USA | North America | 35.67 | -80.47 | 1 | 2 | 20 | 13.2 | 2 | 0.319 | [-0.039, 0.676] | 2 | 0.392 | [0.031, 0.752] |
| San Diego | USA | North America | 32.72 | -117.16 | 3 | 6 | 128 | 14.5 | 6 | 0.406 | [0.196, 0.616] | 6 | 0.332 | [0.191, 0.473] |
| San Francisco | USA | North America | 37.78 | -122.42 | 1 | 1 | 39 | 23.1 | 1 | 0.926 | [0.608, 1.245] | 1 | 0.938 | [0.618, 1.258] |
| South San Diego County | USA | North America | 32.58 | -117.1 | 1 | 1 | 442 | 14.6 | 1 | 0.151 | [0.078, 0.224] | 1 | 0.189 | [0.115, 0.262] |
| Syracuse | USA | North America | 43.05 | -76.15 | 4 | 6 | 106 | 13.5 | 6 | 0.386 | [0.227, 0.546] | 6 | 0.631 | [-0.149, 1.412 |
| TX | USA | North America | 31.26 | -98.55 | 1 | 1 | 100 | 9.9 | 1 | 0.227 | [0.071, 0.382] | 1 | 0.167 | [0.014, 0.321] |
| Tallahassee | USA | North America | 30.44 | -84.28 | 5 | 10 | 124 | 16.8 | 10 | 0.365 | [-0.021, 0.751] | 10 | 0.378 | [0.082, 0.674] |
| Tyler | USA | North America | 32.35 | -95.3 | 1 | 1 | 9 | 12.1 | 1 | 0.321 | [-0.206, 0.849] | 1 | 0.315 | [-0.212, 0.842 |
| University | USA | North America | 34.36 | -89.54 | 1 | 1 | 20 | 11.4 | 1 | 0.436 | [0.071, 0.802] | 1 | 0.384 | [0.024, 0.744] |
| WA | USA | North America | 47.29 | -120.21 | 1 | 1 | 117 | 11.6 | 0 | NA |  | 0 | NA |  |
| Washington | USA | North America | 38.9 | -77.04 | 1 | 1 | 215 | 30.6 | 1 | 0.556 | [0.440, 0.672] | 1 | 0.347 | [0.238, 0.456] |
| Winston-Salem | USA | North America | 36.1 | -80.24 | 1 | 2 | 32 | 12.9 | 2 | 0.026 | [-0.293, 0.345] | 2 | -0.016 | [-0.515, 0.482 |

#### 3 Appendix S1: Supplementary Statistical Methods

##### Effect Size Calculation

Standardised mean change scores using raw-score standardisation (SMCR; Becker 1988; Morris and DeShon 2002) were computed for each arm–outcome combination:

$$g_{ij} = c(n)\,\frac{\bar{M}_{\text{post}} - \bar{M}_{\text{pre}}}{SD_{\text{pre}}}$$

where $\bar{M}_{\text{pre}}$ and $\bar{M}_{\text{post}}$ are the pre- and post-intervention means, $SD_{\text{pre}}$ is the pre-intervention standard deviation, and $c(n)$ is the small-sample bias correction. The sampling variance is

$$v_{ij} = \frac{2(1-r)}{n} + \frac{g^2}{2n}$$

where $r$ is the assumed pre–post correlation and $n$ is the sample size. Pre–post correlations were assumed as $r = 0.70$ for SBP and DBP, $r = 0.65$ for $\text{VO}_2$max, and $r = 0.55$ for HR. Study-level dispersion reporting was verified, with SEM-to-SD correction applied where required, before effect sizes and sampling variances were computed.

##### Three-Level Random-Effects Model

Because many studies contributed multiple arms, three-level random-effects models were fitted:

$$y_{ij} = \mu + u_i + w_{ij} + \varepsilon_{ij}$$

where $y_{ij}$ is the observed effect size for arm $j$ in study $i$; $\mu$ is the overall pooled effect; $u_i \sim N(0, \tau^2_{\text{between}})$ captures between-study heterogeneity; $w_{ij} \sim N(0, \tau^2_{\text{within}})$ captures within-study (between-arm) heterogeneity; and $\varepsilon_{ij}$ is sampling error. Models were estimated by REML in metafor.

##### Meta-Regression

The three-level model was extended with fixed-effect moderators:

$$y_{ij} = \beta_0 + \beta_1 \log(\text{PM}_{2.5,i}) + u_i + w_{ij} + \varepsilon_{ij}$$

and, in adjusted models,

$$y_{ij} = \beta_0 + \beta_1 \log(\text{PM}_{2.5,i}) + \beta_2\,\text{Region}_i + \beta_3\,\text{ExMode}_{ij} + \beta_4\,\text{Duration}_i + \beta_5\,\text{Health}_i + u_i + w_{ij} + \varepsilon_{ij}$$

where Region is a four-level factor (North America [ref], Europe, East Asia, Latin America), ExMode is exercise category (Aerobic [ref], Resistance, Combined, Other), Duration is trial duration category, and Health is health condition category.

##### Cluster-Robust Variance Estimation

All inference used CR2 bias-reduced linearisation standard errors clustered at the study level (clubSandwich; Pustejovsky 2022), with Satterthwaite degrees of freedom.

##### Sensitivity B: Marginal Standardisation

To address potential covariate imbalance across $\text{PM}_{2.5}$ levels within regions, stabilised standardisation weights were applied:

$$sw_{ij} = \min\left(\frac{p_{\text{target}}(\text{cell})}{p_{\text{region}}(\text{cell})},\ 5\right)$$

Weights were capped at 5× to prevent extreme leverage from sparse cells and absorbed into the variance term ($v^*_{ij} = v_{ij}/sw_{ij}$) before model fitting.

#### Nonlinear Dose–Response (Spline) Models

Potential nonlinearity was assessed by replacing the linear $\log(\mathrm{PM}_{2.5})$ term with a natural cubic spline basis with 3 degrees of freedom:

$$y_{ij} = \beta_0 + \sum_k \beta_k B_k(\log \mathrm{PM}_{2.5,i}) + u_i + w_{ij} + \varepsilon_{ij}$$

Nonlinearity was assessed with a likelihood ratio test against the linear model, both fitted with maximum likelihood (Table S2).

#### Outlier Handling

Effect sizes with $|y_{ij}| > 3$ were excluded from primary analyses as outliers; a sensitivity analysis retaining all observations is reported in Table S1.
