## Supplementary material for "Ambient PM_2.5_ Concentration and the Cardiovascular Response to Exercise Training: A Systematic Review and Meta-Analysis Across Global Pollution Gradients": PRISMA 2020 Checklist

| **Section and Topic** | **Item #** | **Checklist item** | **Location where item is reported** |
| --- | --- | --- | --- |
| **TITLE** | | |  |
| Title | 1 | Identify the report as a systematic review. | Title page |
| **ABSTRACT** | | |  |
| Abstract | 2 | See the PRISMA 2020 for Abstracts checklist. | Abstract (structured) |
| **INTRODUCTION** | | |  |
| Rationale | 3 | Describe the rationale for the review in the context of existing knowledge. | Introduction, paras 1–3 |
| Objectives | 4 | Provide an explicit statement of the objective(s) or question(s) the review addresses. | Introduction, final para |
| **METHODS** | | |  |
| Eligibility criteria | 5 | Specify the inclusion and exclusion criteria for the review and how studies were grouped for the syntheses. | Methods, 'Inclusion and Exclusion Criteria' |
| Information sources | 6 | Specify all databases, registers, websites, organisations, reference lists and other sources searched or consulted to identify studies. Specify the date when each source was last searched or consulted. | Methods, 'Search Strategy' (MEDLINE, Embase, PubMed; searched 3 Nov 2025) |
| Search strategy | 7 | Present the full search strategies for all databases, registers and websites, including any filters and limits used. | Full search strategies for all databases were published with the registered protocol on PROSPERO (CRD420251068843); summarised in Methods, 'Search Strategy' |
| Selection process | 8 | Specify the methods used to decide whether a study met the inclusion criteria of the review, including how many reviewers screened each record and each report retrieved, whether they worked independently, and if applicable, details of automation tools used in the process. | Methods, 'Search Strategy' (three reviewers screened records in Rayyan across four systematic stages) |
| Data collection process | 9 | Specify the methods used to collect data from reports, including how many reviewers collected data from each report, whether they worked independently, any processes for obtaining or confirming data from study investigators, and if applicable, details of automation tools used in the process. | Methods, 'Data Extraction and Air Pollution Exposure Assessment' |
| Data items | 10a | List and define all outcomes for which data were sought. Specify whether all results that were compatible with each outcome domain in each study were sought (e.g. for all measures, time points, analyses), and if not, the methods used to decide which results to collect. | Methods, 'Inclusion and Exclusion Criteria' and 'Data Extraction' (SBP, DBP, VO₂max, resting HR) |
|  | 10b | List and define all other variables for which data were sought (e.g. participant and intervention characteristics, funding sources). Describe any assumptions made about any missing or unclear information. | Methods, 'Data Extraction and Air Pollution Exposure Assessment' |
| Study risk of bias assessment | 11 | Specify the methods used to assess risk of bias in the included studies, including details of the tool(s) used, how many reviewers assessed each study and whether they worked independently, and if applicable, details of automation tools used in the process. | Methods, 'Risk of Bias Assessment' (RoB 2 / ROBINS-I; two reviewers) |
| Effect measures | 12 | Specify for each outcome the effect measure(s) (e.g. risk ratio, mean difference) used in the synthesis or presentation of results. | Methods, 'Effect Size Calculation' (standardised mean change, Hedges' g) |
| Synthesis methods | 13a | Describe the processes used to decide which studies were eligible for each synthesis (e.g. tabulating the study intervention characteristics and comparing against the planned groups for each synthesis (item #5)). | Methods, 'Inclusion and Exclusion Criteria'; 'Statistical Analysis' |
|  | 13b | Describe any methods required to prepare the data for presentation or synthesis, such as handling of missing summary statistics, or data conversions. | Methods, 'Effect Size Calculation' (assumed pre–post correlations) |
|  | 13c | Describe any methods used to tabulate or visually display results of individual studies and syntheses. | Figures S4–S5 (forest plots); Figure 6 and Figure S6 (dose–response) |
|  | 13d | Describe any methods used to synthesize results and provide a rationale for the choice(s). If meta-analysis was performed, describe the model(s), method(s) to identify the presence and extent of statistical heterogeneity, and software package(s) used. | Methods, 'Statistical Analysis' (three-level REML models, CR2 cluster-robust variance) |
|  | 13e | Describe any methods used to explore possible causes of heterogeneity among study results (e.g. subgroup analysis, meta-regression). | Methods, 'Statistical Analysis' and subgroup analysis sections (meta-regression, PM₂.₅ strata, exercise mode, hypertension status) |
|  | 13f | Describe any sensitivity analyses conducted to assess robustness of the synthesized results. | Methods, 'Statistical Analysis' (five pre-specified sensitivity analyses; outlier and IET analyses) |
| Reporting bias assessment | 14 | Describe any methods used to assess risk of bias due to missing results in a synthesis (arising from reporting biases). | Methods, 'Statistical Analysis' (Egger's regression, Begg's rank correlation, trim-and-fill) |
| Certainty assessment | 15 | Describe any methods used to assess certainty (or confidence) in the body of evidence for an outcome. | Not reported (formal certainty assessment, e.g. GRADE, was not performed) |
| **RESULTS** | | |  |
| Study selection | 16a | Describe the results of the search and selection process, from the number of records identified in the search to the number of studies included in the review, ideally using a flow diagram. | Results, first para; Figure 1 (PRISMA flow diagram) |
|  | 16b | Cite studies that might appear to meet the inclusion criteria, but which were excluded, and explain why they were excluded. | Figure 1; post hoc exclusions described in Methods, 'Inclusion and Exclusion Criteria' |
| Study characteristics | 17 | Cite each included study and present its characteristics. | Table 1; Table S17 (city-level results); study-level data via Shiny application (Data Availability) |
| Risk of bias in studies | 18 | Present assessments of risk of bias for each included study. | Results, 'Risk of Bias'; Figures S7–S8; Table S16 |
| Results of individual studies | 19 | For all outcomes, present, for each study: (a) summary statistics for each group (where appropriate) and (b) an effect estimate and its precision (e.g. confidence/credible interval), ideally using structured tables or plots. | Figures S4–S5 (forest plots); Table S17 |
| Results of syntheses | 20a | For each synthesis, briefly summarise the characteristics and risk of bias among contributing studies. | Results, 'Pooled Effects' and 'Risk of Bias' |
|  | 20b | Present results of all statistical syntheses conducted. If meta-analysis was done, present for each the summary estimate and its precision (e.g. confidence/credible interval) and measures of statistical heterogeneity. If comparing groups, describe the direction of the effect. | Results; Tables 2–3 (pooled estimates, 95% CIs, prediction intervals, heterogeneity) |
|  | 20c | Present results of all investigations of possible causes of heterogeneity among study results. | Results, 'PM₂.₅ Meta-regression', 'Subgroup Analyses', 'Composition of the High-Pollution Stratum' |
|  | 20d | Present results of all sensitivity analyses conducted to assess the robustness of the synthesized results. | Results, sensitivity analyses (Tables S1, S4–S7, S9, S12); IET sensitivity analysis |
| Reporting biases | 21 | Present assessments of risk of bias due to missing results (arising from reporting biases) for each synthesis assessed. | Results, 'Publication Bias'; Table S15 |
| Certainty of evidence | 22 | Present assessments of certainty (or confidence) in the body of evidence for each outcome assessed. | Not reported (see item 15) |
| **DISCUSSION** | | |  |
| Discussion | 23a | Provide a general interpretation of the results in the context of other evidence. | Discussion, 'Main Findings' |
|  | 23b | Discuss any limitations of the evidence included in the review. | Discussion, 'Strengths and Limitations' |
|  | 23c | Discuss any limitations of the review processes used. | Discussion, 'Strengths and Limitations' |
|  | 23d | Discuss implications of the results for practice, policy, and future research. | Discussion, 'Clinical and Public Health Implications'; Conclusions |
| **OTHER INFORMATION** | | |  |
| Registration and protocol | 24a | Provide registration information for the review, including register name and registration number, or state that the review was not registered. | Methods (PROSPERO CRD420251068843) |
|  | 24b | Indicate where the review protocol can be accessed, or state that a protocol was not prepared. | Protocol accessible via PROSPERO record CRD420251068843 |
|  | 24c | Describe and explain any amendments to information provided at registration or in the protocol. | Post hoc decisions (exclusion of pre-2003 studies and one telephone-delivered intervention) described in Methods |
| Support | 25 | Describe sources of financial or non-financial support for the review, and the role of the funders or sponsors in the review. | Funding statement (NIHR Leicester BRC, NIHR203327; funders had no role) |
| Competing interests | 26 | Declare any competing interests of review authors. | Declaration of Competing Interests |
| Availability of data, code and other materials | 27 | Report which of the following are publicly available and where they can be found: template data collection forms; data extracted from included studies; data used for all analyses; analytic code; any other materials used in the review. | Data Availability (analysis dataset and R code via R Shiny application; CAMS ERA5 publicly available) |

*From:*  Page MJ, McKenzie JE, Bossuyt PM, Boutron I, Hoffmann TC, Mulrow CD, et al. The PRISMA 2020 statement: an updated guideline for reporting systematic reviews. BMJ 2021;372:n71. doi: 10.1136/bmj.n71

For more information, visit: <http://www.prisma-statement.org/>
